# A Foundation Model for Intensive Care: Unlocking Generalization across Tasks and Domains at Scale

**DOI:** 10.1101/2025.07.25.25331635

**Authors:** Manuel Burger, Daphné Chopard, Gregor Lichtner, Malte Londschien, Fedor Sergeev, Moritz Fuchs, Hugo Yèche, Rita Kuznetsova, Martin Faltys, Eike Gerdes, Polina Leshetkina, Micha Christ, Moritz Schanz, Nora Göbel, Peter Bühlmann, Elias Grünewald, Felix Balzer, Gunnar Rätsch

**Affiliations:** Department of Computer Science, ETH Zurich, Switzerland; Department of Intensive Care and Neonatology and Children’s Research Center, University Children’s Hospital Zurich, University of Zurich, Switzerland; Department of Anesthesia, Intensive Care, Emergency and Pain Medicine, Universitätsmedizin Greifswald, Germany; Institute of Medical Informatics, Charité – Universitätsmedizin Berlin, Germany; Seminar for Statistics, ETH Zurich, Switzerland; AI Center, ETH Zurich, Switzerland; Helmholtz AI, Germany; Center for Medical Data Integration, Bosch Health Campus, Stuttgart, Germany; Department of Intensive Care Medicine, University Hospital and University of Bern, Switzerland; University Hospital Zurich, Switzerland; Department of Health Science and Medicine, University of Luzern, Switzerland; Department of General Internal Medicine and Nephrology, Robert Bosch Hospital, Stuttgart, Germany; Department of Cardiovascular Surgery, Robert Bosch Hospital, Stuttgart, Germany; Swiss Institute for Bioinformatics, Zurich, Switzerland; Medical Informatics Unit, University Hospital Zurich, Switzerland

## Abstract

**BACKGROUND:** Early recognition of physiological deterioration in critically ill patients enables timely intervention, yet much of the predictive information in routinely collected intensive care data remains underutilized. Existing prediction models are typically developed for a single task at a single institution and often degrade across hospitals with differing patient populations and treatment patterns. Whether a model pretrained on harmonized multi-institutional data can generalize across both hospitals and tasks (dual zero-shot) without task adaptation or local retraining has not been systematically evaluated.

**METHODS:** We harmonized 16 critical care datasets from ICUs and emergency departments across the United States, Europe, and Asia, comprising over 1.1 million patient stays and over one billion data points. We pretrained ICareFM, a transformer-based foundation model, on 130 harmonized clinical concepts to estimate the probability that physiological values will cross clinician-specified thresholds within a given time horizon, enabling flexible event definitions without retraining. We evaluated ICareFM on seven prediction tasks (circulatory, respiratory, kidney, and liver organ failure, hyperglycemia, sepsis, and mortality) across nine external ICU cohorts using a local patient equivalence (LPE) framework that quantifies how many labeled patients a locally trained model requires to match ICareFM performance.

**RESULTS:** Without task- or hospital-specific training (dual zero-shot), ICareFM achieved a median AuROC of 0.837 (95% CI, 0.797–0.858) across held-out datasets and seven prediction tasks, and outperformed task-matched clinical scores used in intensive care by AuROC +0.049 (95% CI, 0.031–0.071). It matched the performance of specialized local models trained on over 1,000 labeled patient stays. With task- and hospital-specific adaptation (staged adaptation), locally trained models required a median of 14,709 patient stays to match ICareFM; in 84% of settings, the available local training data was insufficient to train a model that would outperform staged-adapted ICareFM by a 0.5% AuROC margin. External validation on large cohorts at Robert Bosch Krankenhaus (Germany) and Charité – Universitätsmedizin Berlin (Germany) revealed staged-adapted ICareFM outperformed local models trained on over 60,000 and 100,000 patient stays, respectively. Beyond the ICU, adapted models outperformed locally trained alternatives in 9/10 emergency department and general ward settings. Pairing ICareFM with large language models enabled natural language risk queries that outperformed both native LLM prediction and clinical scoring baselines.

**CONCLUSIONS:** A foundation model pretrained on harmonized, multi-continental critical care data generalized across hospitals and clinical tasks, reducing local data requirements and, with staged adaptation, matching or outperforming locally trained models in the majority of evaluated settings. The data harmonization code, processing pipelines, and model weights are released to support independent validation and further research.

## 1 Introduction

Timely recognition of physiological deterioration is central to improving outcomes in critically ill patients ^1^. Intensive care units (ICUs) and emergency departments (EDs) continuously capture high-frequency physiological data, including vital signs, laboratory measurements, ventilator settings, and medication records, that reflect the rapidly evolving state of critical illness ^2–4^. Yet detecting subtle multivariate patterns in these data remains challenging under the cognitive load and time pressure of acute care ^5^, and much of the predictive information in routinely collected data remains underutilized.

Rule-based clinical scores ^6–8^ and task-specific machine learning models ^1;9–12^ have been developed to predict outcomes such as organ failure and mortality. While effective within their training cohorts, most are designed for a single task and trained on data from a single institution, and their performance degrades under distribution shifts across hospitals with different patient populations, measurement practices, and treatment policies ^12;13^. In our experience and that of others, this lack of robustness forces costly redevelopment at each new institution and has limited the scalability of data-driven decision support in critical care.

Foundation models — neural networks pretrained on diverse datasets that generalize across domains and tasks — have improved prediction in medical imaging ^14^, clinical text ^15;16^, and other modalities ^17–20^. In critical care, similar approaches using physiological time series have shown promise ^21–23^, but remain confined to single institutions or narrowly defined benchmarks, with limited multi-continental data harmonization or systematic cross-hospital evaluation ^1;12;24;25^. Consequently, the prevailing assumption that “all models are local”, i.e., that predictive models rarely generalize beyond their training institution, remains largely unchallenged ^26^.

Existing healthcare foundation models leave this gap largely unaddressed. Models trained on structured electronic health record (EHR) codes ^25;27–29^ require fewer labeled examples but rely on retrospective abstractions that are often unavailable at the bedside in real time. Physiology-first models, as discussed above, have not yet scaled to multi-institutional training or systematic cross-hospital evaluation. General-purpose forecasting and LLM approaches face different limitations: the former targets value prediction rather than clinical event risk ^30–32^ and has not been evaluated in treatment-confounded critical care settings; the latter, although convenient to deploy, struggles with high-volume numerical time series data ^33^. The combination of large-scale multi-institutional data harmonization, flexible event definition without retraining, and systematic dataset-level holdout evaluation for high-frequency critical care physiology has, to our knowledge, not been systematically addressed.

This gap reflects structural barriers to large-scale data harmonization. Differences in variable definitions, measurement frequencies, and treatment encoding complicate cross-institutional data integration. These challenges have practical and equity implications: while small and medium-sized hospitals constitute the majority of healthcare institutions worldwide (in the United States, 62% of hospitals have fewer than 100 beds) ^34;35^, critical care artificial intelligence (AI) research remains concentrated on datasets from large academic centers ^36^. Hence, current approaches risk reinforcing disparities in access to data-driven clinical tools. We argue that developing models which generalize across hospitals without requiring local retraining is essential for equitable clinical decision support, particularly for these institutions.

Deploying a foundation model in a new hospital poses a dual generalization challenge. The model should ideally generalize across *tasks*, performing clinical predictions it was not explicitly trained on, and simultaneously across *institutions*, operating under distribution shifts in patient populations, measurement practices, and treatment policies. In critical care, these shifts are substantial: hospitals differ in which variables are measured, how frequently they are recorded, and how care is delivered, effectively making patient records from different institutions distinguishable by their recording patterns alone. In the machine learning literature, these two axes have been studied independently as task-level ^37;38^ and domain-level ^39;40^ “zero-shot” generalization; each alone is difficult. Their combination, which we term the *dual zero-shot* setting, has not been systematically addressed in critical care.

To address this challenge, we developed **ICareFM**, a foundation model for critical care designed for the dual zero-shot setting, predicting clinical deterioration events across hospitals without task-specific or site-specific retraining. ICareFM is pretrained on a harmonized dataset of vital signs, laboratory values, medication rates, and ventilator settings from ICUs and EDs across three continents. A time-to-event pretraining objective operates directly on continuous physiological values, allowing clinicians to specify clinical threshold-based events at prediction time, including through a natural language interface via LLMs. We evaluate ICareFM using a leave-dataset-out protocol in which entire hospital datasets are held out, alongside a data-efficiency framework that quantifies when a pretrained model surpasses locally developed alternatives, providing guidance for institutions choosing between local development and pretrained deployment.

## 2 Methods

### 2.1 Dataset creation and harmonization

#### Study cohort

We assembled a harmonized critical care time series dataset (**Figure 1 A**) from 16 sources across three continents: North America (MIMIC-III ^41^, MIMIC-IV ^42^, eICU ^43^, NWICU ^44^), Europe (AmsterdamUM-Cdb ^45^, SICdb ^3^, HiRID ^2^, unpublished Personalized Swiss Sepsis Study: PSSS), and Asia (PICdb ^46^, Zigong EHR ^47^, INSPIRE ^48^), supplemented by two U.S. ED datasets (MIMIC-IV-ED ^49^, MC-MED ^4^) and a structured EHR dataset (EHRSHOT ^27^). A large private cohort from Charité – Universitätsmedizin Berlin (Germany) and a private cohort from the Robert Bosch Krankenhaus (RBKICU) Stuttgart (Germany) were used for validation purposes. Dataset statistics are summarized in Tables S1 and S2. All datasets were de-identified and accessed under the data use agreements and institutional review board approvals associated with the original studies.

**Figure 1:**
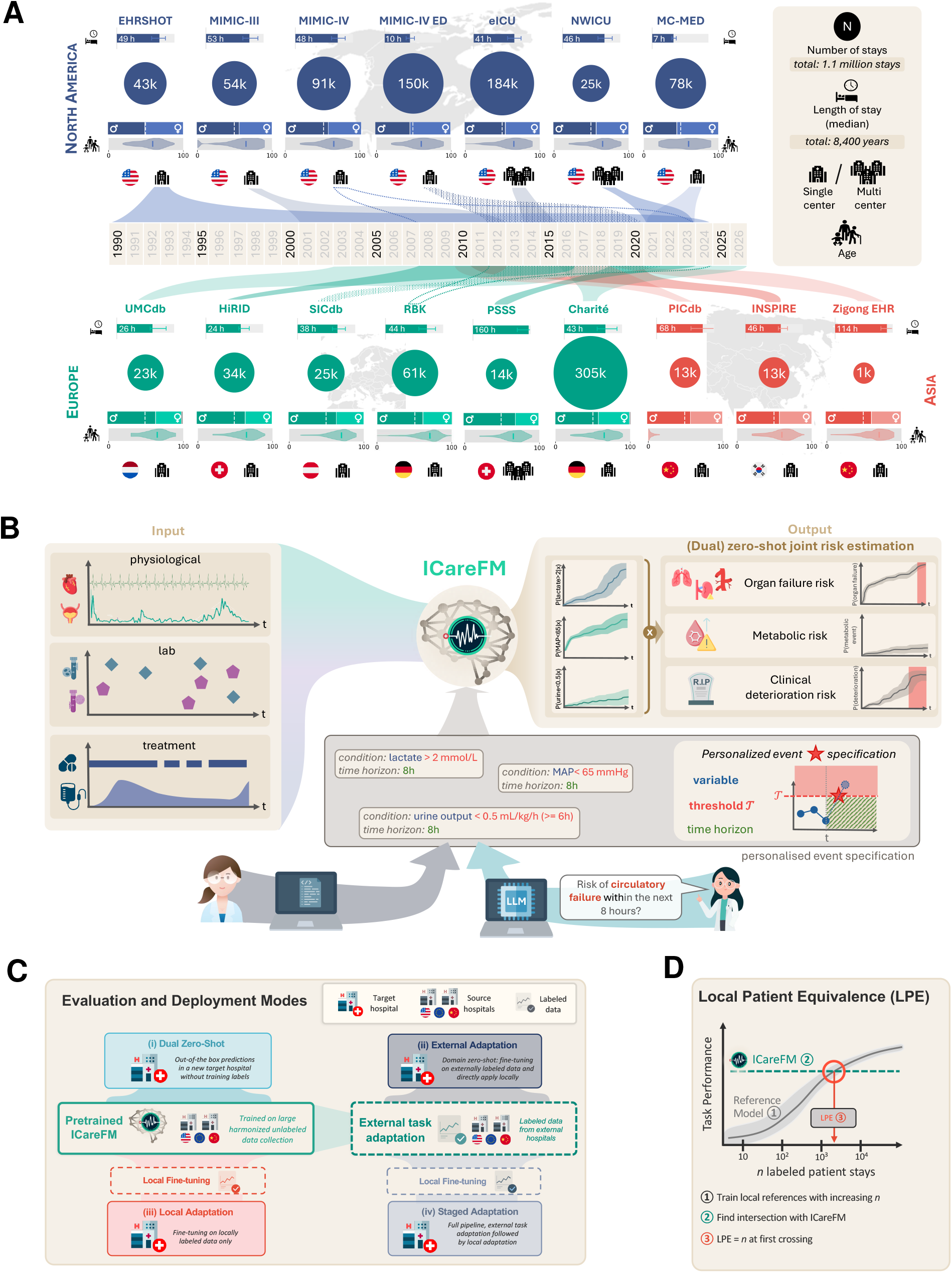
Data, Model, Local Patient Equivalence, and Deployment. **A)** Overview of core datasets. Summary statistics of included cohorts; more details in the Supplements. **B)** Model overview; routine intensive care time series data is processed by ICareFM to produce survival predictions. Queries to ICareFM are expressed as a triplet of target variable, critical threshold, and time horizon. Multiple individual queries are combined to produce estimates for complex outcomes. LLMs can translate clinical questions into complex queries to ICareFM. Further details on zero-shot inference mechanisms are in the Supplements. **C)** Flow of (labeled) source data and (labeled) target data across the different deployment modes. **D)** Illustration of local patient equivalence (LPE) determination.

#### Data harmonization

Building on the ricu^50^ framework, we defined 130 harmonized clinical concepts spanning static demographics, vital signs, laboratory values, and treatments across 16 source datasets, guided by intensivist input (Supplementary A). We extracted administration rates for core intensive care medications (vasopressors, sedatives, anticoagulants, and loop diuretics) together with treatment indicators (e.g., any antibiotic use), yielding approximately 40 treatment concepts. These were mapped from over 16,000 raw descriptors across the sources using LLM-assisted classification ^51^ and verified by physicians (Supplementary A).

#### Inclusion criteria

We included encounters with a length of stay of at least 4 hours, at least four recorded measurements, and a maximum measurement gap of at most 48 hours. After applying these criteria and *k*-anonymization, we extracted a sparse hourly grid for model input. Further details are provided in Supplementary A.

#### Task annotations

We evaluated ICareFM on clinically relevant real-time early event prediction tasks including circulatory failure ^1^, severe respiratory failure ^12^, severe kidney failure ^24^, liver failure (MELD-based) ^52^, hyperglycemia ^53^, mortality (decompensation) ^54^, and Sepsis-3^9;55^. For ED cohorts we additionally considered hypotension and hypoxia ^56^. All tasks were formulated as binary early event prediction with 2− 48 hours prediction horizons. Events were defined as physiological deterioration states derived from routinely collected clinical data (Supplementary A). We additionally varied event-defining thresholds to assess behavior across severity levels. A task is only benchmarked if all required clinical variables are recorded in the target dataset; across nine ICU datasets and seven benchmark tasks, we observe *N* = 50 distinct evaluation settings.

### 2.2 ICareFM development

#### Threshold-conditioned time-to-event pretraining

We developed a self-supervised pretraining objective tailored to continuous physiological monitoring. Conditioned on the patient’s history, the model estimates the time-dependent probability that a core set of target clinical variables (*K*=35, Supplementary B.2.1) will cross a threshold *τ* in a specified direction, while the threshold was randomly varied during training to learn from a diverse set of events. A time-step encoder and causal Transformer ^57^ produce hourly patient state representations (Supplementary B). For each target variable, a dedicated hazard prediction head maps the patient state and queried threshold to a discrete hazard function over a 48-hour horizon via learned threshold and direction embeddings. Treatments were excluded from the target set to reduce learning of hospital-specific treatment patterns. Total model size is approximately 30M parameters with further details in Supplementary B.

#### Zero-shot Inference

At inference, each survival head yields the cumulative failure probability *F*_*k*_(*h* |**H**_*t*_, *τ*_*k*_): the probability that variable *k* will cross a queried threshold *τ*_*k*_ within horizon *h*, given the encoded patient state **H**_*t*_. Composite clinical events are approximated by aggregating univariate failure probabilities through conjunctions and disjunctions under conditional independence (Supplementary B.2.2). For example, 8-hour circulatory failure risk is estimated as *F*_MAP_(8h |**H**_*t*_, < 65 mmHg) ·*F*_Lact._(8h | **H**_*t*_, >2 mmol/l). This allows composing clinical threshold queries at inference time without retraining (**Figure 1 B**). Because the composition follows clinical event definitions rather than learned classifiers, and the representations generalize across hospitals through multi-institutional pretraining, this mechanism can achieve simultaneous task and domain generalization. Sweeping *τ* across discretized bins of each variable’s value range further enables zero-shot approximation of composite clinical scores (Eqs. (7) and (8), Supplementary B.2.2). For some benchmark endpoints, the zero-shot query serves as a clinical proxy rather than an exact label match; for example, sepsis risk is approximated via SIRS-like criteria and mortality via an APACHE score approximation (Supplementary B.2.2).

#### ICareFM inference and deployment modes

We evaluated four deployment scenarios (**Figure 1 C**), all excluding the target dataset (“local hospital”) from pretraining (Supplementary A). For modes involving local data, we systematically varied training set size to assess data efficiency (Supplementary C.1, Supplementary C.4):

*(i) Dual zero-shot*: survival heads are combined according to clinical event definitions without any task-specific or site-specific training; *(ii) External adaptation*: task-specific fine-tuning using only external labeled data, then direct deployment to the target dataset (domain zero-shot, i.e., domain generalization); *(iii) Local adaptation*: fine-tuning using only local labeled data, representing settings with novel endpoints unavailable externally; *(iv) Staged adaptation*: external adaptation followed by additional local fine-tuning.

Mode (i) represents the dual zero-shot setting, requiring neither task-specific training data nor target-site data; modes (ii)–(iv) progressively relax this constraint by introducing task-specific or site-specific labeled data.

### 2.3 Local patient equivalence evaluation

To quantify the practical value of pretraining, we introduced the **local patient equivalence (LPE)** (**Figure 1 D**). For each target hospital, prediction task, and training-set subsample size, we trained reference models using increasing numbers of local patients. Reference models included gradient-boosted trees ^58^ with statistical features ^59^ and deep neural networks ^10;60^. For each training budget, we selected the best-performing algorithm and hyperparameters using a validation set drawn from the same hospital (Supplementary C.1) as part of the same budget. Performance was measured by the area under the receiver operating characteristic curve (AuROC). The LPE was defined as the *first* intersection between ICareFM’s AuROC curve and the best-performing local reference model’s AuROC curve. We extrapolate curves up to at most twice the available data; when no intersection is observed within this range, the LPE is right-censored at the extrapolation limit, yielding a conservative lower bound. This quantity represents the number of locally labeled patients required for a supervised model to match the performance of the pretrained or adapted foundation model (**Figure 1 C**; Supplementary C.4).

## 3 Results

### 3.1 A million-scale cohort supports training a critical care foundation model: ICareFM

We harmonized 16 critical care datasets from ICUs, EDs, and general hospital admissions across three continents into a unified clinical time series representation (Figure 1 A; Tables S1 and S2). After inclusion and anonymization (Supplementary A) of 1.5 million candidate stays, the final cohort comprised over 1.1 million patient stays summing to over 8,400 patient-years with more than one billion data points. After dataset splitting and leave-dataset-out procedures, the median ICareFM pretraining dataset contained 390,000 stays (up to 650,000 for validation at Charité).

We trained ICareFM on different subsets of the harmonized cohort using a leave-dataset-out protocol, enabling systematic evaluation of both task and hospital generalization. ICareFM’s threshold-conditioned time-to-event objective outperformed alternative self-supervised objectives trained on the same dataset and architecture in a linear classifier setting on the fixed patient state representations across nine ICU datasets and seven downstream tasks. For each baseline, we report the mean AuROC difference relative to ICareFM, paired across datasets and tasks. Among objectives trained on the same data and architecture, forecasting-based approaches ^61;62^ (ΔAuROC = +0.022) and time-to-event formulations designed for coded electronic health records ^28;63^ (+0.029) both underperformed ICareFM (Figure S2 and Supplementary B.6). Existing foundation models pretrained on external data performed substantially worse: Chronos-2^31^ (+0.318) and MOTOR ^28^ (+0.163). Adapting Chronos-2 on the harmonized dataset narrowed but did not close this gap (+0.259).

### 3.2 ICareFM predicts clinical deterioration across hospitals without local training

We evaluated ICareFM in the dual zero-shot setting, generalizing across hospitals and tasks, using the local patient equivalence (LPE) framework (Section 2.3, Supplementary C.1), which quantifies how many labeled patients a locally trained model would need to match ICareFM’s performance. We report 95% confidence intervals by bootstrap resampling across datasets and tasks (Supplementary C.4.2). **Figure 2 A and B** show LPEs at thousands of local patient stays, indicating task and hospital generalization.

**Figure 2:**
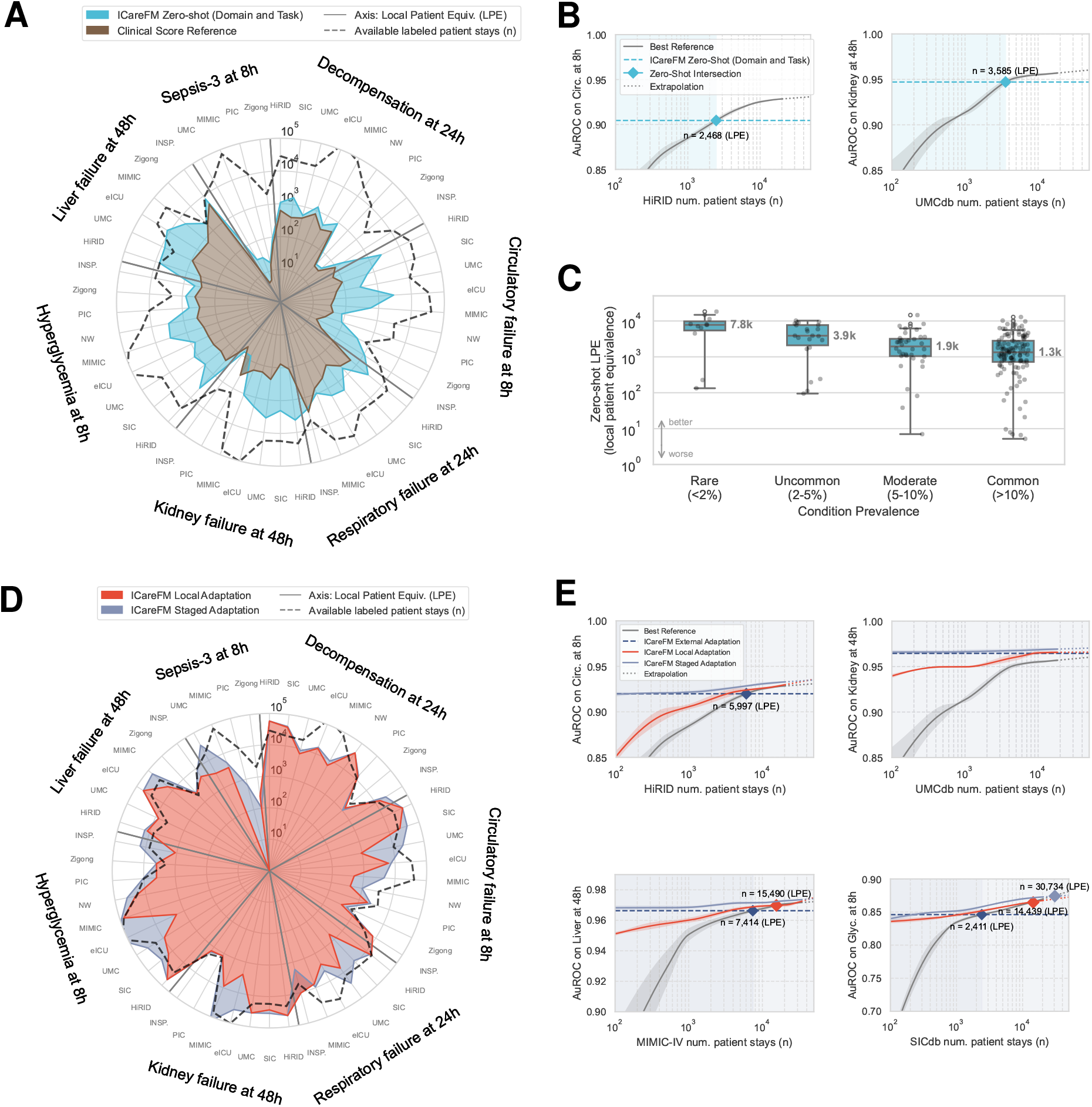
ICareFM generalization measured by local patient equivalence (LPE). Performance across nine ICU datasets and seven benchmark tasks, evaluated using the LPE framework. **A)** Dual zero-shot LPE by task and dataset, shown relative to clinical scores and available local labeled patient stays. **B)** Example scaling curves showing dual zero-shot AuROC as a function of local training set size, with LPE intersection points against locally trained reference models. **C)** Dual zero-shot LPE as a function of event severity across five tasks with varying thresholds. **D)** LPE by task and dataset across external, local, and staged adaptation settings, shown relative to available local labeled patient stays. **E)** Example scaling curves showing adapted ICareFM AuROC as a function of local training set size, with LPE intersection points against locally trained reference models.

Across nine ICU datasets and seven prediction tasks, the median LPE was 1,025 (95% CI, 595–1,796; IQR, 405–2,413). The median AuROC was 0.837 (95% CI, 0.797–0.858). Compared with commonly used task-matched clinical scores, ICareFM achieved stronger discrimination across tasks (median AuROC improvement, +0.049; 95% CI, 0.031–0.071). By modulating severity for five tasks (circulatory, respiratory, kidney, and liver failure, and hyperglycemia), we observed increased LPE for rarer and more severe events (**Figure 2 C**). We observed that performance remained limited in settings with sparse training coverage, large distribution shifts, or events difficult to express as threshold queries. These included pediatric transfer prediction (PICdb; median LPE, 75), low-density cohorts (Zigong; median LPE, 387), and sepsis prediction (median LPE, 29), whose onset depends on delayed blood-culture confirmation, making the label extremely sensitive to recording practices ^55^. These tasks benefited most from task-specific adaptation and local adaptation (Section 3.3).

A natural concern with externally trained models is whether they introduce fairness disparities compared to locally trained alternatives. We assessed fairness across three available demographic properties (sex, age, and ethnicity) and found no aggregate evidence of increased disparity relative to local reference models (Section C.13).

### 3.3 Adaptation progressively improves prediction and reduces data requirements

We also evaluated ICareFM’s adaptation across the three settings described in Section 2.2 (**Figure 2 D/E**).

#### External adaptation

To assess the ability of domain generalization, we used externally labeled data to tune ICareFM to specific tasks, yielding a median LPE of 2,921 (95% CI, 2,057–4,485), corresponding to an absolute median AuROC of 0.869 (95% CI, 0.831–0.917). US and European institutions reached median LPEs of 4,570 and 4,461, respectively. Aggregate fairness metrics across three available demographic properties remained comparable to those of local reference models (Section C.13).

#### Local adaptation

Adapting ICareFM using only local labeled data, relevant when novel biomarkers for labeling are unavailable externally, increased the median LPE to 7,167 patients (95% CI, 3,687–12,073). ICareFM already matches (within 0.5% AuROC) or outperforms local models at full available training data 76% of the time. ICareFM also outperformed a task-specific solution targeting kidney failure ^64^ on a heart surgical subcohort of RBKICU (Section C.15).

#### Staged adaptation

Combining external task adaptation with subsequent local domain adaptation enabled ICareFM to match (within 0.5% AuROC) or outperform locally trained models using the full available labeled training data in 84% (strictly outperforming 66%) of evaluated settings (median AuROC improvement, +0.00373; 95% CI, 0.00073–0.00644). The overall median LPE was 14,709 (95% CI, 8,323–30,218); this estimate is right-censored by available dataset sizes (Section C.4.3), as ICareFM outperformed the local reference at full training scale in the majority of settings. LPE estimates on the public development cohorts are limited by dataset size, data density (Supplementary C.8), and in the case of sepsis, label quality; as shown below, external validation on larger cohorts yields substantially higher LPEs. Staged adaptation LPE remained consistent across AuROC, AuPRC, and event-level recall at low false-positive rates (≤1%) (Supplementary C.4.4, Supplementary C.10).

### 3.4 Scaling a diverse pretraining cohort improves generalization

#### Scaling improves generalization

A power-law fit to pretraining dataset scaling (**Figure 3 A**) suggested a scaling ratio of approximately 5:3, indicating that *fivefold* more external data yields *threefold* gains in local patient equivalence. Dual zero-shot yielded a decreased, approximately square root, transfer efficiency of 4:2 (Section C.5). Extrapolating the fitted power law to the scale of large EHR vendors ^65^ yielded a projected LPE of 135,877 patients (95% CI, 51,721–363,865), assuming continued scaling behavior. Expected calibration error (ECE) likewise decreased with increasing pretraining dataset size (**Figure 3 B**).

**Figure 3:**
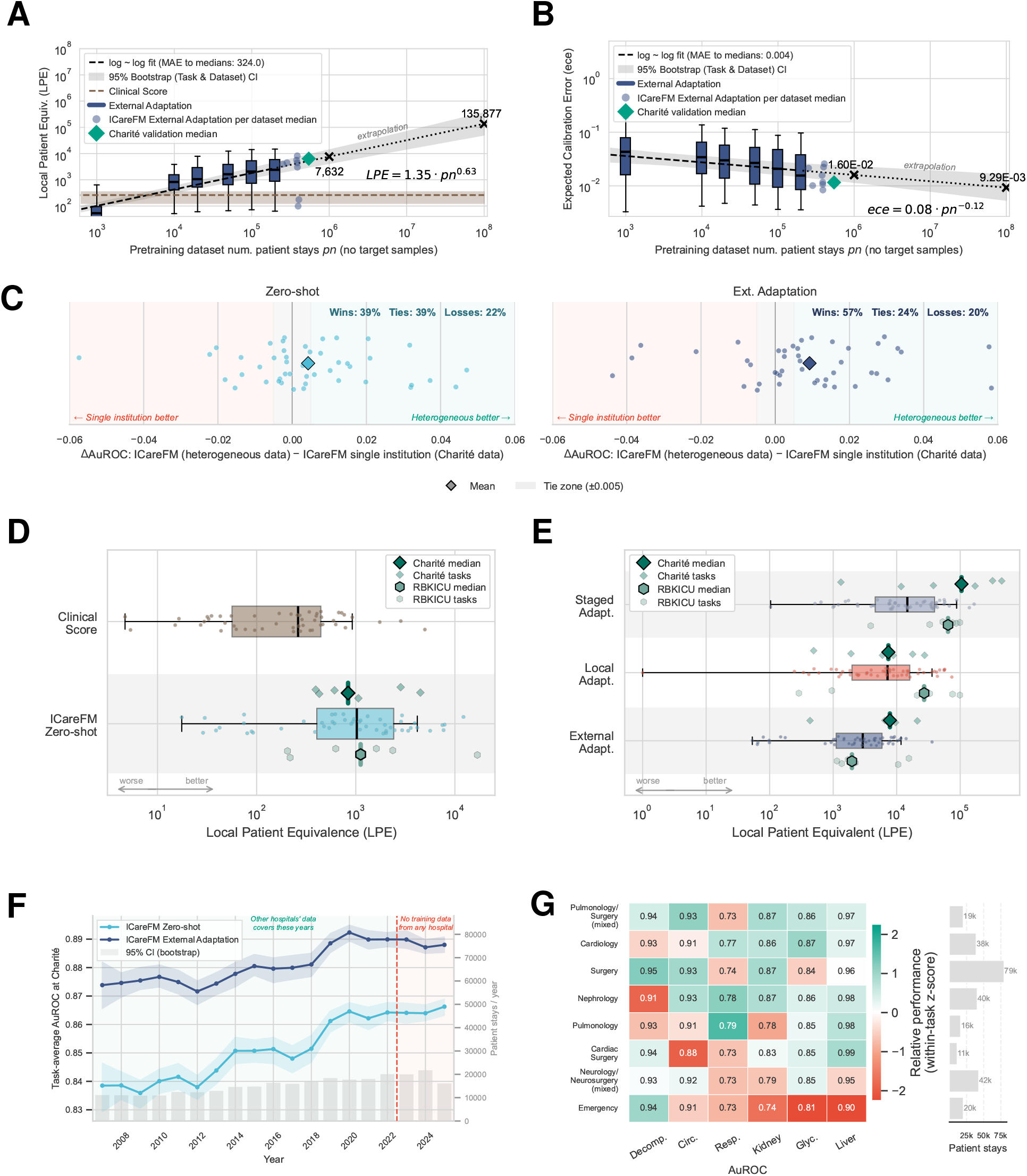
Scaling ICareFM and validating its generalization bounds. **A)** Local patient equivalence (LPE) of externally adapted models as a function of pretraining dataset size, with power-law fit. **B)** Expected calibration error of externally adapted models as a function of pretraining dataset size, with power-law fit. **C)** Comparison of ICareFM trained on a heterogeneous multi-institutional subsample with a sample-size-matched model trained solely on Charité data (250,000 patient stays), evaluated on held-out target datasets. **D)** Dual zero-shot LPE across tasks and datasets, shown relative to clinical scores, with validation on the Charité and RBKICU cohorts. **E)** LPE across tasks and datasets for external, local, and staged adaptation settings, with validation on the Charité and RBKICU cohorts. **F)** AuROC of dual zero-shot and externally adapted ICareFM transferred to the Charité cohort over time. The largest change point coincides with the 2019 migration to a new critical care information system; performance remains stable beyond the 2022 pretraining data cutoff. **G)** AuROC of externally adapted models stratified by ward specialization in the Charité cohort, colored by within-task relative performance.

The Charité cohort was used to extend the pretraining dataset and confirmed the scaling trends established in **Figure 3 A and B** for AuROC-based LPE (22% deviation) and calibration (33% deviation), respectively.

#### Data diversity improves generalization

We further assessed the role of training data diversity by comparing ICareFM with a sample-size-matched model trained solely on Charité data (250,000 patient stays; **Figure 3 C**). We find that a heterogeneously trained ICareFM transfers better to an unseen hospital with 39% wins vs. 22% losses given a 0.5% equivalence margin on AuROC in dual zero-shot mode. The gap widens when labels are used to externally task-adapt the model to 57% wins vs. 20% losses.

### 3.5 External validation supports ICareFM’s generalization claims

After developing ICareFM, collaborating sites independently harmonized two further cohorts. We shipped the pretrained model; all data harmonization, inference, and adaptation were performed onsite. Validation on the independent German cohorts (Charité and RBKICU) yielded a median dual zero-shot LPE of 845 patients (**Figure 3 D**), closely following the predicted LPE of 1,025 labeled patient stays.

When externally adapted, ICareFM achieved an LPE of 7,055 on the independent German cohorts, which exceeded the predicted 2,921 and associated confidence interval (**Figure 3 E**). Local adaptation validated to an LPE of 13,335, exceeding the forecasted 7,167. Staged adaptation validated to a median LPE of 64,368 and 104,754 on the RBKICU and Charité cohorts, respectively, with the Charité LPE observed within the interpolation regime. These results confirm that the previously estimated aggregate median LPE was conservative due to right-censoring attributable to smaller datasets and data quality limitations.

### 3.6 ICareFM’s performance remains stable across time and clinical settings

We assess ICareFM’s performance over time when transferred to the Charité cohort in dual zero-shot and external adaptation mode (**Figure 3 F**). Transfer performance on the Charité cohort remained stable beyond ICareFM’s 2022 training cutoff, and the largest change point was correlated with the migration to a newer critical care data management system in 2019. Stratifying ICareFM’s performance across specialized wards (**Figure 3 G**) does not reveal any concerning performance deviations. However, the model tends to perform worst in settings where patients are most unstable and events more challenging to predict accurately (e.g., emergency wards).

### 3.7 Learned representations capture temporal progression and organ-failure risk

Beyond predictive performance, we examined the internal representations underlying ICareFM’s generalization behavior. Principal component analysis on patient state representations revealed organization around clinically meaningful factors rather than hospital-specific artifacts (Figure S25). **Figure 4 A** shows temporal progression along the first principal component; trajectory flow fields show that patients approaching death deviate from population trends and converge toward a distinct high-risk region. Organ failure events localize to distinct subregions, with circulatory, respiratory, and kidney failure each exhibiting characteristic spatial distributions (**Figure 4 B**). An individual trajectory (**Figure 4 C**) shows the zero-shot risk score rising as creatinine increases, before formal KDIGO ^66^ criteria are met, suggesting that ICareFM captures early signs of deterioration.

**Figure 4:**
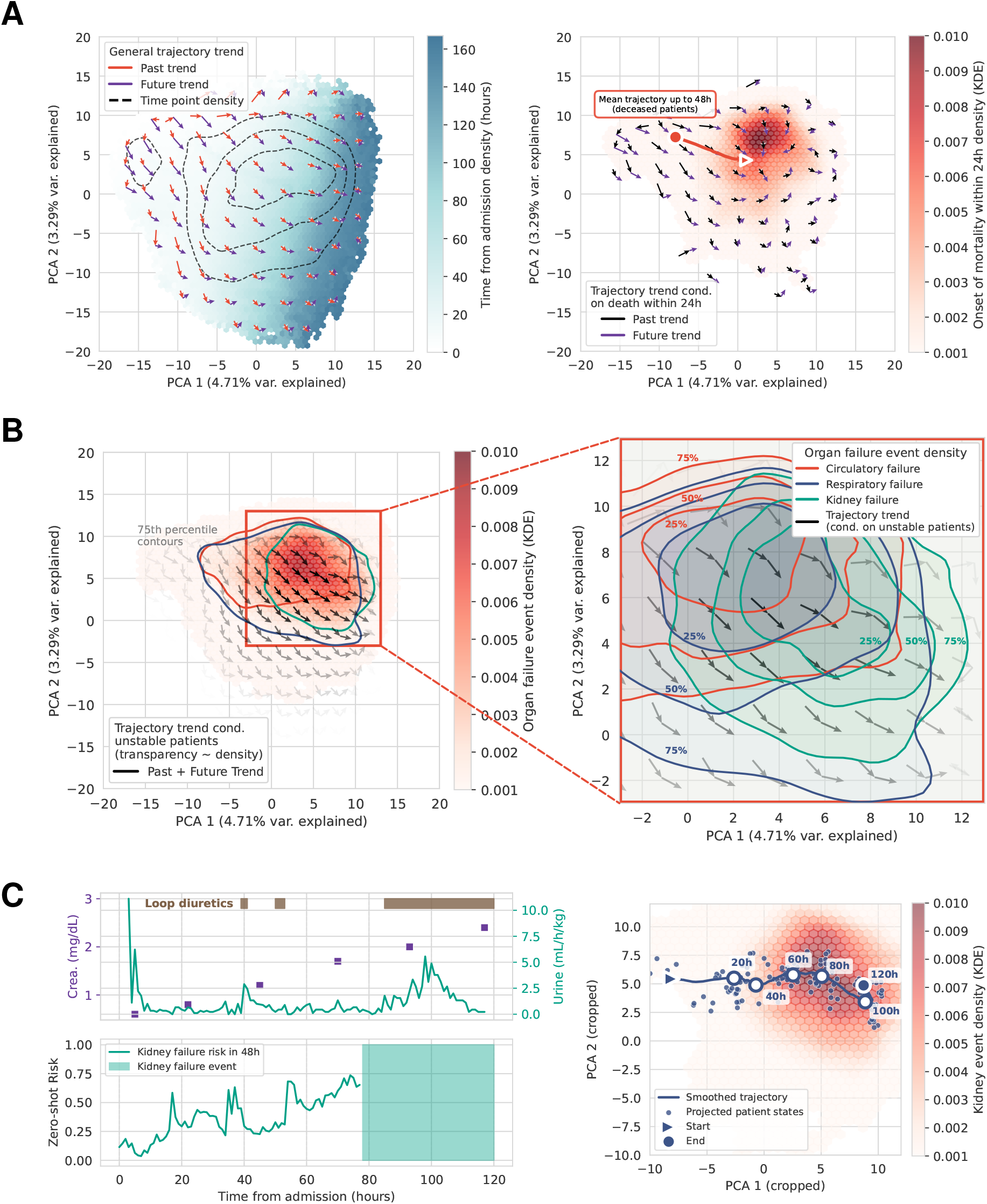
ICareFM learned representations and latent space structure. **A)** PCA projection of patient state embeddings from held-out test patients across seven ICU datasets. *(Left)* Embeddings colored by length of stay. Arrows indicate grid-averaged trajectory directions from preceding (red) and subsequent (purple) time points; dashed contours mark overall density boundaries. *(Right)* Density estimate of embeddings within 24 hours of mortality, with trajectory flow fields of dying patients showing convergence toward a high-risk region. **B)** Organ failure events in the embedding space. *(Left)* Density estimates of organ failure events with trajectory flow among unstable patients (i.e., patients with organ failure throughout stay); arrow transparency scales with sample density. *(Right)* Magnified view of circulatory (red), respiratory (blue), and kidney (green) failure distributions. **C)** Individual patient trajectory through latent space. *(Left)* Kidney function parameters (urine output and creatinine), stage 3 acute kidney injury annotation based on KDIGO guidelines, and ICareFM zero-shot risk score over time. *(Right)* Trajectory of a patient with severe kidney failure, shown against the population density of kidney failure events.

### 3.8 ICareFM generalizes beyond the ICU

We evaluated whether ICareFM generalizes from ICU training data to ED settings characterized by sparser monitoring and more heterogeneous acuity. Transfer to ED cohorts from Boston ^49^ and Stanford ^4^, as well as a general hospital admissions dataset (EHRSHOT), transferred well (**Figure 5 A–C**). Zero-shot and external adaptation achieved median LPEs of 3,641 and 10,879, respectively. Under staged adaptation, ICareFM outperformed locally trained ED and general ward models in 9 of 10 benchmarked settings on the full available local training dataset.

**Figure 5:**
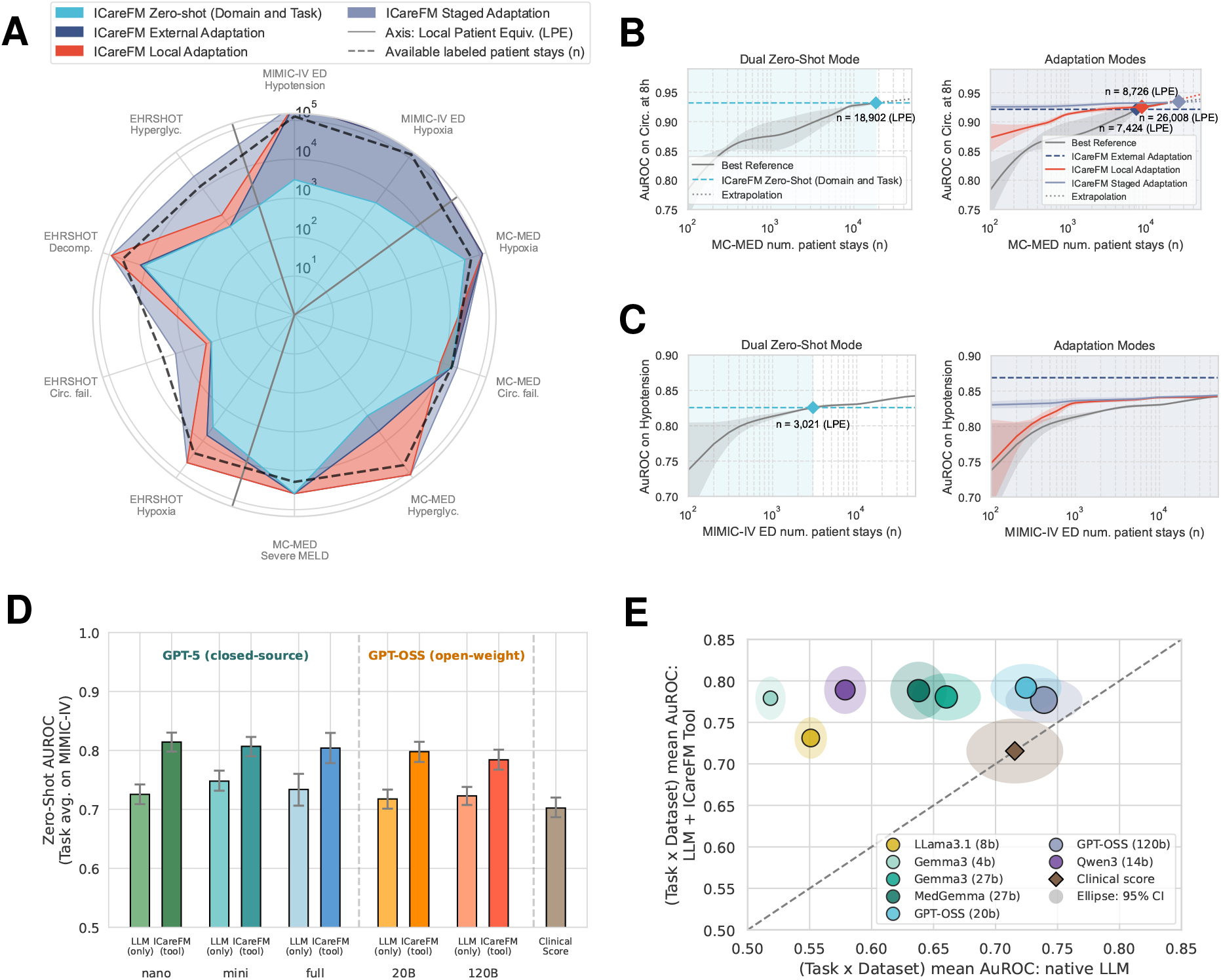
Transfer beyond the ICU and integration with large language models. **A)** LPE across deployment modes for emergency department (MIMIC-IV ED, MC-MED) and general hospital admission (EHRSHOT) cohorts. **B)** Example scaling curves from MC-MED (circulatory failure) under dual zero-shot (left) and adaptation (right) settings. **C)** Example scaling curves from MIMIC-IV ED (hypotension) under dual zero-shot (left) and adaptation (right) settings. **D)** Task-average dual zero-shot AuROC on MIMIC-IV for direct LLM prediction (GPT-5, GPT-OSS), LLM tool-calling ICareFM, and clinical scores. Error bars indicate 95% bootstrap confidence intervals. **E)** Task-average dual zero-shot AuROC across nine ICU datasets and seven tasks for open-weight LLMs of varying size in direct prediction and tool-calling ICareFM mode.

### 3.9 Natural language interfaces unlock ICareFM for clinical use

As a standalone model, ICareFM requires technical expertise to configure threshold queries, limiting direct use by clinicians. General-purpose LLMs offer a more accessible natural language interface and can themselves attempt zero-shot clinical predictions from time series data, making them a natural alternative. By integrating ICareFM as a tool within several LLMs via tool calling (Figure 1 B), we combine the strengths of both: the LLM translates natural language clinical queries into structured threshold and horizon specifications, which ICareFM resolves through probabilistic inference. Compared with direct LLM prediction from compressed time series input, tool-calling ICareFM significantly improved predictive performance (**Figure 5 D**; AuROC, +0.070; 95% CI, 0.048–0.094 on MIMIC-IV for GPT-5). Smaller open-weight models (4B–27B parameters) also achieved effective tool-calling performance across nine ICU datasets (**Figure 5 E**, Supplementary C.6).

## 4 Discussion

In this retrospective multicenter study, we developed and evaluated a critical care foundation model that addresses the dual zero-shot challenge of generalizing across tasks and institutions ^19;20^. Across nine ICU datasets and seven benchmark tasks, ICareFM achieved a median local patient equivalence (LPE) of 1,025 patient stays in the dual zero-shot setting, a median AuROC of 0.837, and a median AuROC advantage of 0.049 over commonly used clinical scores. Adaptation further increased median LPE to 2,921 with external labels, 7,167 with local labels, and 14,709 with staged adaptation; in the staged setting, the model matched or exceeded locally trained reference models in 84% of evaluations. External validation at Charité – Universitätsmedizin Berlin and Robert Bosch Krankenhaus, performed after model development had been completed, confirmed these estimates. With staged adaptation at Charité and Robert Bosch, ICareFM surpassed local models trained on over 100,000 and 60,000 patient stays, respectively, indicating that development-set LPE estimates were conservative due to limited public dataset sizes. These findings relax the assumption that clinical prediction models must be local ^26^ and suggest that harmonized physiological pretraining can reduce the need for repeated model redevelopment.

We designed the LPE framework to provide practical deployment guidance. Institutions whose locally available labeled data falls below the LPE estimates reported here are expected to achieve stronger predictive performance by deploying ICareFM than by training a model on local data alone. This does not replace local validation or governance, but makes the tradeoff between external pretraining and local development explicit. We observed roughly threefold gains in LPE for every fivefold increase in external pretraining data, with parallel improvement in calibration, although this depends on continued scaling beyond the data studied here. Just as importantly, we found that a heterogeneous pretraining corpus transferred better than a sample-size-matched single-center model, indicating that diversity of source hospitals matters in addition to scale. This provides empirical support, in the setting of large-scale foundation model pretraining, for mathematical results predicting that diversity across training environments improves generalization ^67;68^. For health systems and EHR vendors considering foundation model development, this suggests that pooling data from diverse institutions, even at smaller per-site volumes, may yield more transferable models than scaling within a single system. The largest gains were seen for rarer and more severe events, which are also the outcomes for which any single hospital is least likely to have sufficient labeled data. For example, a small community hospital without local training data could deploy ICareFM in dual zero-shot mode and achieve deterioration prediction equivalent to a locally trained model requiring over 1,000 labeled patient stays.

The threshold-conditioned pretraining objective is central to this flexibility. Rather than being limited to a fixed list of endpoints defined during development, ICareFM estimates the probability that a physiological variable or composite state will cross a clinician-specified threshold within a chosen time horizon. This makes the model queryable at inference time without retraining and supports flexible risk assessment, for example by defining kidney injury relative to an individualized baseline creatinine or by using SpO_2_/FiO_2_-based surrogates when arterial blood gases are unavailable ^66;69^. We find that this approach is most reliable for simple or moderately complex physiological events and less robust for endpoints with delayed ascertainment, sparse measurements, or stronger dependence on treatment context.

The learned representations generalize beyond the ICU training domain. ICareFM transferred to emergency department and general ward settings, with median LPEs of 3,641 in zero-shot mode and 10,879 after external adaptation, and staged adaptation outperformed locally trained ED and general ward models in 9 of 10 bench-marked settings. Latent-space analyses suggested that patient states were organized primarily by physiological trajectory and organ-failure risk rather than by hospital-specific artifacts. This interpretation is consistent with the temporal validation at Charité, where performance remained stable beyond the 2022 cutoff of the pretraining data.

At the same time, we found that transfer performance was significantly associated with recording density across nine external datasets, with data density explaining approximately half the variance in zero-shot LPE across sites (Supplementary C.8). Sites with denser vital sign monitoring, more frequent laboratory sampling, and more accurate treatment recording achieved substantially higher patient equivalence. Similarly, the largest temporal change point at Charité coincided with the 2019 migration of the critical care information system, further illustrating that data infrastructure and recording practices are important determinants of transfer utility.

The LLM experiments also clarify how language models may best be used in this setting. Direct LLM prediction from compressed time series inputs only marginally surpassed conventional scores ^33^. By contrast, using an LLM to translate natural-language clinical questions into structured ICareFM queries substantially improved predictive performance. That small open-weight models were sufficient for this routing task suggests a practical path to privacy-preserving local deployment, with the LLM acting as an interface while calibrated risk estimation remains with the specialized physiological model.

This study has several limitations. Pediatric data were represented by a single dataset, and performance was weaker in that population. The model operates on an hourly grid and may miss higher-frequency physiological dynamics. Although treatment variables were included as inputs, they were excluded from pretraining targets to reduce learning of institution-specific treatment patterns, so disentangling treatment effects from physiological deterioration remains an open problem. Sepsis prediction was particularly challenging (median LPE, 29), reflecting not only clinical complexity but also substantial cross-site variation in how sepsis onset is labeled ^55^, underscoring that label harmonization is as important as data harmonization for cross-hospital evaluation. Fairness analyses were reassuring in aggregate across the available demographic attributes, but relied primarily on summary disparity metrics and the harmonized data did not permit a more complete assessment across all clinically relevant subgroups. Formal memorization auditing of the model weights was not performed; privacy considerations for model release are discussed in the Data and Code Availability section. Although external validation was performed after model development on independently harmonized cohorts, all evaluations used retrospective patient data; effects on clinician workflow, decision-making, and patient outcomes remain to be established in prospective studies.

These findings suggest that clinical prediction models do not always need to be developed separately at each hospital. The data harmonization code and processing pipelines, spanning all public sources with thousands of raw variable descriptors mapped to 130 clinical concepts, are released so that the cohort can be reproduced from individually obtained datasets via their respective data access mechanisms. Model weights are shared under a data use agreement to support independent validation and further scaling of multi-site pretraining. A cluster-randomized crossover trial evaluating an early warning system based on a predecessor of ICareFM is registered and recruiting ^70^. If confirmed in prospective studies, this approach could provide a reusable and more equitable foundation for acute care decision support, especially for hospitals that lack the data volume, infrastructure, or regulatory flexibility required to repeatedly build models from scratch.

## Data Availability

All the source datasets are available via PhysioNet (Goldberger et al., 2000) upon completion of CITI's "Data or Specimens Only Research" course (HiRID and SICdb require additional project specific provider approval). Further datasets are accessible directly via the providers (UMCdb and EHRSHOT).

https://physionet.org/

https://github.com/ratschlab/icarefm

## Acknowledgments

We gratefully acknowledge the patients and their families whose willingness to share health-care data makes this work possible. We also thank the clinicians, data engineers, and investigators who collected, curated, de-identified, and released the intensive-care datasets on which this study relies. Their sustained commitment to open, high-quality data has created an extraordinary research community that continues to advance critical-care science. Specifically, this work uses and acknowledges the datasets: MIMIC-III, MIMIC-IV, and MIMIC-IV-ED, eICU Collaborative Research Database 2.0, AmsterdamUMCdb, HIRID (High time-resolution ICU dataset), Northwestern ICU (NWICU) database, Salzburg Intensive Care database (SICdb), Pediatric Intensive Care database (PICdb), Critical care database comprising patients with infection at Zigong Fourth People’s Hospital (Zigong EHR), EHRSHOT, INSPIRE (a publicly available research dataset for perioperative medicine), Multimodal Clinical Monitoring in the Emergency Department (MC-MED), PSSS data from the Swiss Personalized Sepsis Study, data from the Charité – Universitätsmedizin Berlin, and Robert Bosch Krankenhaus Stuttgart.

This work was supported as part of the Swiss AI Initiative (swiss-ai.org) by a grant from the Swiss National Supercomputing Centre (CSCS) under project ID a02 on Alps.

Computational data analysis was performed at Leonhard Med (https://sis.id.ethz.ch/services/sensitiveresearchdata/) secure trusted research environment at ETH Zurich. SNF Funding (213236) to Andre Kahles contributed to compute resources.

The project was supported by grant #2022-278 of the Strategic Focus Area “Personalized Health and Related Technologies (PHRT)” of the ETH Domain (Swiss Federal Institutes of Technology) and ETH Core funding (to Gunnar Rätsch).

Malte Londschien was supported by the ETH Foundations of Data Science and the ETH AI Center. Fedor Sergeev was supported by grant #902 of the Strategic Focus Area “Personalized Health and Related Technologies (PHRT)” of the ETH Domain (Swiss Federal Institutes of Technology). Daphné Chopard received funding from grant #2021-911 of the Strategic Focus Area “Personalized Health and Related Technologies (PHRT)” of the ETH Domain (Swiss Federal Institutes of Technology).

A big thank you to the Swiss AI Research Platform team (https://serving.swissai.cscs.ch), especially Robert Matthew Smith, Xiaozhe Yao, and Imanol Schlag, for serving LLMs to the research community.

We would like to thank Zeynep Babür, Jasmina Bogojeska from ZHAW School of Engineering for their contributions to extracting further harmonized data points from the MIMIC-IV dataset.

We would like to thank the Swiss AI community members and associated students participating in our weekly meetings to push the state-of-the-art in health care time series modeling further for the discussions and their feedback, most notably: Alexander Morgenroth, Fabian Baldenweg, Simon Boutry, and Xiaochen Zheng. Fedor Sergeev thanks Vincent Fortuin for his helpful suggestions to improve the manuscript. We would also like to thank Julia E. Vogt for her support during the duration of this project.

This research was conducted using data obtained from Charité’s Health Data Platform (HDP) / Medical Data Integration Center (MeDIC) (https://hdp.charite.de). Data provision was supported by the Service Unit Data Science (SU-DS) of the Institute of Medical Informatics (IMI) at Charité (https://medinfo.charite.de) and the BIH Core Facility Research IT. The authors acknowledge the Scientific Computing team of the Business Division IT for providing computational resources (HPC). We thank Sophie K. Piper for her support with statistical planning.

We further thank the meDIC team at Robert-Bosch-Krankenhaus (RBK), especially Nico Schmid for his support and enabling this work, by initiating the data extraction that formed the foundation of the RBKICU dataset. We are also grateful to Dr. med. Sebastian Allgäuer for his valuable insights. We acknowledge the Robert Bosch Centrum für Innovationen im Gesundheitswesen for supporting this work as part of the ETH-BHC Joint Fellowship Program.

Figures are created using resources from Flaticon (https://www.flaticon.com; icons by HAJICON, AmethystDesign, Freepik, juicy_fish, iconset.co, and GeekClick) and Bioicons (https://bioicons.com; icons by Marcel Tisch, Simon Dürr, and Servier).

## Ethics

No study protocol was prepared. Due to the retrospective nature of the study, no patient or public involvement was required.

This project received an ethics waiver from the cantonal ethics committee of the canton of Zurich, Switzerland (
`BASEC-Nr.: Req-2024-00528`) to work with the considered collection of retrospective datasets. Furthermore, ethical approval was obtained from the Institutional Review Board (EK) of Charité – Universitätsmedizin Berlin (approval no.: `EA2/291/25`) and the study was registered in the German Clinical Trials Register under no. DRKS00038236. In addition, the study at Robert Bosch Krankenhaus was approved by the Ethics Committee of the Medical Faculty of the University of Tübingen (approval no.: `963/2021BO2`).

## Data and Code Availability

### Harmonized large-scale dataset

If legally and organizationally feasible, we aim to publish the harmonized and processed multi-center dataset (or a subset thereof) on the PhysioNet platform ^71^, making a large collection of critical care time-series data easier to access.

### Source datasets

Most source datasets are available via PhysioNet ^71^ upon completion of CITI’s “Data or Specimens Only Research” certification (HiRID and SICdb require additional project specific provider approval). Further datasets are accessible directly via the providers (UMCdb ^45^ and EHRSHOT ^27^). The small cohort from PSSS (Personalized Swiss Sepsis Study) is not publicly available. The Charité cohort is not publicly available. The RBKICU cohort is not publicly available, but we support any external researchers evaluating alternative methods.

### Data Harmonization Tooling

To harmonize the different source datasets, we built upon the ricu^50^ tool. Upon publication we will release our fork of the package containing code and configuration to harmonize the source datasets and full multi-center dataset. We are additionally working on a python re-implementation of the package making the tooling accessible to Python users.

### Data processing

Code and other required resources to reproduce the full data processing pipeline to obtain the harmonized multi-center dataset (public data subset) will be added or linked in the project’s root GitHub repository https://github.com/ratschlab/icarefm.

### Model development and experiments

Code and other required resources to reproduce model training and evaluation experiments will be added or linked in the project’s root GitHub repository https://github.com/ratschlab/icarefm.

### Model weights

Pretrained model weights will be shared under a data use agreement, aiming to be consistent with the access requirements of the source datasets used during training. Because the model was trained on patient-level physiological data, even though de-identified, the release of model weights requires consideration of memorization and potential disclosure risk. Formal memorization auditing was not performed in this study. We recommend that systematic privacy assessments, including membership inference testing and disclosure-risk evaluation, be conducted before any clinical deployment to complement the de-identification and governance protections already in place for the source datasets.

## Author Contributions

**Conceptualization and study design:** M.B. and G.R. co-conceptualized the study; R.K. contributed to study design and analytical methodology. **Data harmonization and preparation:** M.B., D.C. (including EHRSHOT integration and feedback on additional data sources), M.L. (pipeline design and execution), F.S. (including medication and treatment integration lead), and H.Y. (guidance on processing and harmonization decisions). **Charité cohort integration and validation:** G.L. (harmonization, integration, and validation lead), El.G. (organization, ethical approval, clinical data extraction, and feedback on data integration and experimental design), and F.B. (supervision, data access, and ethical approval). **RBKICU cohort curation and evaluation:** M.Fu. (data curation, preprocessing, and evaluation lead), M.C. (dataset development and percentage-based outcomes evaluation), M.S. (data curation and harmonization feedback), and N.G. (ethical approval and oversight). **Clinical expertise:** M.Fa. (variable selection and harmonization, medical interpretation of results), Ei.G. and P.L. (clinical verification of treatment variable mappings). **Model design and development:** M.B. (model design), H.Y. (architecture, objective function, and training advice), and R.K. (model development feedback). **Experiments, evaluation, and visualization:** M.B. (lead), D.C. (latent space visualization, analysis and interpretation of results, and figure creation), G.L. (Charité validation experiments), M.Fu. (RBKICU evaluation), F.S. (fairness and bias evaluation design and execution), M.L. (baselines and evaluation methodology), and H.Y. (evaluation methodology) **Statistical methodology and experimental design:** P.B. (supervision), M.L. and G.R. (continuous feedback throughout the study). **Funding, data access, and coordination:** G.R. (grant funding, data access approvals, ethics approval, computational resources, and coordination of collaborations). **Manuscript drafting:** M.B., D.C., and F.S. **Manuscript review:** All authors critically reviewed the manuscript; G.R. additionally revised. All authors approved the final version of the manuscript and agree to be accountable for all aspects of the work.

## AI Use Disclosure

Generative AI tools (including large language models such as OpenAI ChatGPT and Anthropic Claude, as well as code-completion assistants such as GitHub Copilot) were used during the preparation of this manuscript to assist with prose editing, literature synthesis, and software development. All AI-generated content was critically reviewed, edited, and verified by the authors, who take full responsibility for the accuracy, integrity, and originality of the final work. No AI tool was used to generate scientific ideas, interpret results, or draw conclusions. In accordance with ICMJE recommendations, no AI tool is listed as an author.

## Notes

### Competing Interest Statement

The authors have declared no competing interest.

### Funding Statement

This work was supported as part of the Swiss AI Initiative (https://swiss-ai.org) by a grant from the Swiss National Supercomputing Centre (CSCS) under project ID a02 on Alps.
Computational data analysis was performed at Leonhard Med (https://sis.id.ethz.ch/services/sensitiveresearchdata/) secure trusted research environment at ETH Zurich. SNF Funding (213236) to Andre Kahles contributed to compute resources.
The project was supported by grant #2022-278 of the Strategic Focus Area "Personalized Health and Related Technologies (PHRT)" of the ETH Domain (Swiss Federal Institutes of Technology) and ETH Core funding (to Gunnar R"atsch).
Malte Londschien was supported by the ETH Foundations of Data Science and the ETH AI Center. Fedor Sergeev was supported by grant #902 of the Strategic Focus Area "Personalized Health and Related Technologies (PHRT)" of the ETH Domain (Swiss Federal Institutes of Technology). Daphne Chopard received funding from grant #2021-911 of the Strategic Focal Area "Personalized Health and Related Technologies (PHRT)" of the ETH Domain (Swiss Federal Institutes of Technology).

### Author Declarations

The Zurich Cantonal Ethics Committee waived ethical approval for this work (REQ-2024-00528).

### Summary of Updates

This revision expands the dataset from 650,000 to over 1.1 million patient stays, adds post-development external validation at two independent German health systems (Charite and Robert Bosch Krankenhaus), and introduces the local patient equivalence (LPE) framework for quantifying when pretrained deployment outperforms local model development. New analyses include dual zero-shot evaluation across both tasks and hospitals, LLM integration experiments, scaling laws for pretraining data size and diversity, temporal stability validation, and fairness assessments. The manuscript text has been substantially revised throughout.

