## Supplementary material for "A Foundation Model for Intensive Care: Unlocking Generalization across Tasks and Domains at Scale"

### Table of Contents


**A Data** **S3**

A.1 Data collection . . . S3
A.2 Data harmonization . . . S3
A.3 Concept reference table . . . S8
A.4 Data processing . . . S10
A.5 Task annotations . . . S11
A.6 K-anonymization . . . S14

**B Model** **S14**

B.1 Architecture . . . S14
B.2 Objective function . . . S15
B.3 Survival prediction head conditioning . . . S17
B.4 Context to related foundation model approaches . . . S18
B.5 Model size - Ablation study . . . S19
B.6 Pretraining objective - Ablation study . . . S20
B.7 Survival horizon - Ablation study . . . S23
B.8 Bivariate events - Ablation study . . . S23
B.9 Training . . . S24

**C Evaluation** **S25**

C.1 Baseline model selection and tuning . . . S25
C.2 Extended comparison . . . S27
C.3 Clinical score references . . . S28
C.4 Determining local patient (performance) equivalence (LPE) . . . S29
C.5 Complementary task zero-shot results with domain adaptation . . . S32
C.6 Language model experiments . . . S35
C.7 Survival head evaluation . . . S39
C.8 Data missingness sensitivity analysis . . . S40
C.9 Zero-shot OOD calibration . . . S41
C.10 Complementary adaptation results . . . S42
C.11 PSSS Study: same-country cross-hospital transfer analysis . . . S42
C.12 Latent space analysis . . . S44
C.13 Fairness analysis . . . S46
C.14 External Validation: Charité – Universitätsmedizin Berlin . . . S47
C.15 External Validation: Robert Bosch Krankenhaus . . . S48

### A Data

#### A.1 Data collection

The overview of datasets is presented in Table S1 and Table S2. Most of these datasets are available freely via PhysioNet[1] or directly from the providers (UMCdb[2] and EHRSHOT[3]). We do not include RICD[4] because it is not freely accessible (a separate contract and payment are required). PSSS, Charité, and RBKICU cohorts are private.

| | MIMIC-III | MIMIC-IV | eICU | NWICU | HiRID | UMCdb | SICdb | PICdb | Zigong EHR |
|---|---|---|---|---|---|---|---|---|---|
| **Demographic information** | | | | | | | | | |
| Country | USA | USA | USA | USA | Switzerland | Netherlands | Austria | China | China |
| Multi-Center | No | No | Yes | Yes | No | No | No | No | No |
| Years | 2001 - 2012 | 2008 - 2022 | 2014 - 2015 | 2020 - 2022 | 2008 - 2016 | 2003 - 2016 | 2013 - 2021 | 2010 - 2018 | 2019 - 2020 |
| Num. Stays | 53,632 | 91,039 | 183,581 | 24,943 | 33,547 | 22,883 | 24,510 | 13,268 | 2,525 |
| Gender (% Female) | 43.6 | 44.0 | 45.9 | 43.1 | 35.7 | 34.0 | 36.6 | 42.6 | 39.2 |
| Gender (% Male) | 56.4 | 56.0 | 54.1 | 56.9 | 64.3 | 63.9 | 63.4 | 57.4 | 60.8 |
| Age (Mean (Std)) | 58.9 (24.6) | 63.2 (16.6) | 63.3 (17.0) | 63.7 (16.2) | 63.5 (15.2) | 62.6 (16.5) | 67.1 (14.6) | 2.2 (3.9) | 66.0 (16.5) |
| Ethnic (% A/B/W/O) | 2.8/9.3/71.4/16.5 | 3.1/10.9/66.9/19.1 | 1.6/10.7/77.3/10.4 | 3.1/13.2/67.7/16.0 | - | - | - | 99.6/-/-/0.4 | - |
| LoS (Median hours (IQR)) | 53 (29, 114) | 48 (27, 94) | 41 (22, 75) | 46 (25, 92) | 24 (20, 52) | 26 (21, 90) | 38 (24, 78) | 68 (23, 214) | 114 (65, 259) |
| **Patient stay prevalence** (num. stays (%)) | | | | | | | | | |
| Mortality | 4,123 (7.7) | 8,244 (9.1) | 9,529 (5.2) | 973 (3.9) | 2,001 (6.0) | 2,242 (9.8) | 1,334 (5.4) | 888 (6.7) | 533 (21.1) |
| Circ. Failure | 7,720 (14.4) | 18,644 (20.5) | 14,356 (7.8) | 1,919 (7.7) | 10,392 (31.0) | 5,657 (24.7) | 7,665 (31.3) | 2,357 (17.8) | 833 (33.0) |
| Sepsis-3 | 17,143 (32.0) | 31,485 (34.6) | - | - | - | 5,357 (23.4) | - | 1,872 (14.1) | 944 (37.4) |
| Hyperglyc. | 15,350 (28.6) | 27,974 (30.7) | 64,257 (35.0) | 9,018 (36.2) | 15,226 (45.4) | 12,833 (56.1) | 7,746 (31.6) | 5,185 (39.1) | 1,365 (54.1) |
| Resp. Failure | 5,594 (10.4) | 13,597 (14.9) | 11,340 (6.2) | - | 3,970 (11.8) | 4,794 (21.0) | 4,616 (18.8) | - | - |
| Kidney Failure | 4,308 (8.0) | 10,771 (11.8) | 19,050 (10.4) | - | 1,375 (4.1) | 1,827 (8.0) | 1,641 (6.7) | 133 (1.0) | - |
| Liver Failure | 2,249 (4.2) | 4,652 (5.1) | 3,224 (1.8) | - | 472 (1.4) | 154 (0.7) | - | - | 99 (3.9) |
| **Time-step label prevalence** (positive %, (negative %, unknown %)) | | | | | | | | | |
| Decomp. 24h | 1.9 (98.1, 0.0) | 2.6 (97.4, 0.0) | 1.7 (98.3, 0.0) | 1.1 (98.9, 0.0) | 2.4 (97.6, 0.0) | 2.8 (97.2, 0.0) | 1.8 (98.2, 0.0) | 1.3 (98.7, 0.0) | 3.4 (96.6, 0.0) |
| Circ. Fail. 8h | 1.4 (35.3, 63.2) | 2.5 (46.6, 50.9) | 0.8 (21.5, 77.7) | 0.8 (9.8, 89.4) | 3.7 (63.3, 33.0) | 2.0 (43.5, 54.5) | 4.2 (68.0, 27.8) | 1.4 (1.9, 96.7) | 3.5 (0.5, 96.0) |
| Sepsis-3 8h | 1.4 (54.5, 44.1) | 1.6 (54.0, 44.4) | - | - | - | 0.6 (49.6, 49.8) | - | 0.5 (84.8, 14.6) | 1.4 (87.2, 11.4) |
| Hyperglyc. 8h | 4.9 (34.9, 60.2) | 7.8 (39.9, 52.3) | 13.6 (49.6, 36.8) | 13.5 (43.9, 42.7) | 14.9 (55.8, 29.4) | 12.1 (71.4, 16.5) | 8.7 (78.3, 13.0) | 4.3 (32.2, 63.6) | 6.9 (15.6, 77.5) |
| Resp. Fail. 24h | 2.9 (20.7, 76.4) | 5.1 (23.1, 71.8) | 2.2 (17.5, 80.3) | - | 5.0 (36.4, 58.6) | 7.5 (47.5, 45.0) | 3.8 (27.7, 68.5) | - | - |
| Kidney Fail. 48h | 2.4 (58.9, 38.7) | 3.9 (78.1, 18.0) | 4.6 (58.8, 36.6) | - | 2.5 (77.9, 19.6) | 2.6 (82.7, 14.7) | 2.6 (77.0, 20.4) | 0.4 (11.0, 88.6) | - |
| Liver Fail. 48h | 3.1 (13.2, 83.6) | 4.3 (22.9, 72.8) | 1.4 (8.7, 89.9) | - | 1.5 (18.0, 80.5) | 0.4 (16.6, 83.0) | - | - | 1.2 (24.6, 74.1) |
| **Data density** (Median % (IQR %)) | | | | | | | | | |
| Vital Density | 26.9 (22.6, 30.9) | 27.2 (23.9, 31.0) | 23.3 (20.3, 28.1) | 16.2 (0.0, 20.1) | 34.0 (27.9, 43.2) | 35.2 (26.9, 43.4) | 26.0 (22.3, 29.6) | 0.7 (0.3, 1.8) | 0.7 (0.4, 0.9) |
| Lab Density | 2.7 (1.9, 3.7) | 3.1 (2.0, 4.6) | 2.0 (1.4, 2.9) | 2.0 (1.4, 2.8) | 5.5 (3.0, 7.6) | 8.8 (5.7, 11.5) | 8.9 (7.2, 10.5) | 5.0 (2.2, 8.1) | 2.2 (1.5, 3.0) |
| Vital Density adj. | 26.9 (22.6, 30.9) | 28.2 (24.8, 32.1) | 23.3 (20.3, 28.1) | 52.2 (0.0, 64.7) | 36.5 (30.0, 46.4) | 37.8 (28.9, 46.7) | 44.4 (38.1, 50.6) | 1.4 (0.6, 3.5) | 1.5 (1.0, 2.0) |
| Lab Density adj. | 2.7 (2.0, 3.8) | 3.3 (2.1, 4.8) | 2.2 (1.4, 3.1) | 2.4 (1.7, 3.5) | 6.4 (3.5, 8.7) | 9.0 (5.9, 11.7) | 10.1 (8.1, 11.9) | 5.4 (2.4, 8.7) | 2.8 (1.9, 3.7) |

**Table S1:** Dataset statistics showing demographic information, patient prevalences for different downstream tasks, time-step prevalence for specific early event prediction labels of each downstream task, data densities for vitals and laboratory tests (regularly and irregularly measured variables, respectively). Adjusted densities (adj.) exclude variables that are not observed for a specific dataset. Statistics for further harmonized data sources are summarized in Table S2. Ethnicity is encoded as A: Asian, B: Black, W: White, O: Other/Unknown.

#### A.2 Data harmonization

We include the following published datasets from the USA (MIMIC-III (v1.4)[5], MIMIC-IV (v3.1)[6], eICU (v2.0)[7], and NWICU (v0.1.0)[8]), Europe (AmsterdamUMCdb (v1.0)[2] as UMCdb, SICdb (v1.0.6)[9], and HiRID (v1.1.1)[10]), and Asia (PICdb (v1.1.0)[11], Zigong EHR (v1.1)[12], INSPIRE (v1.3)[13]). Additionally, we incorporate two emergency department (ED) datasets, MIMIC-IV-ED (v2.2)[14] and MC-MED (v1.0.1)[15]. We further harmonize a structured electronic health records dataset EHRSHOT (v2.1)[3], consisting of coded data but offering a relevant amount of vital measurements, lab test results, and treatment information.

We also harmonize and analyze unpublished data from the Personalized Swiss Sepsis Study (PSSS). Finally, after developing the model, additional private cohorts from the Charité – Universitätsmedizin Berlin (Germany) and Robert Bosch Krankenhaus Stuttgart (Germany) were harmonized and used for additional validation of the established generalization claims and scaling laws.

**MIMIC-III** The "Medical Information Mart for Intensive Care" is a large single-center dataset from the Beth Israel Deaconess Medical Center in Boston, Massachusetts. It contains data from 2001–2014. It has relatively high data density, accurate reporting of treatment rate administration, and ventilation information. The medical center switched their critical care data management system from Philips CareVue Clinical Information System to iMDsoft Metavision ICU in 2008. There are hence already within a single dataset harmonization efforts required to fully model the entire dataset patient population. We extract individual intensive care stays from this dataset.

| | EHRSHOT | MIMIC-IV ED | INSPIRE | PSSS | MC-MED | Charité | RBKICU |
|---|---|---|---|---|---|---|---|
| **Demographic information** | | | | | | | |
| Country | USA | USA | South Korea | Switzerland | USA | Germany | Germany |
| Multi-Center | No | No | No | Yes | No | No | No |
| Years | 1990 - 2023 | 2011 - 2019 | 2011 - 2020 | 2019 - 2022 | 2020 - 2022 | 2006 - 2025 | 2012 - 2024 |
| Num. Stays | 42,562 | 149,546 | 13,250 | 13,565 | 77,824 | 305,458 | 61,554 |
| Gender (% Female) | 51.1 | 53.8 | 42.9 | 31.0 | 55.0 | 41.1 | 38.4 |
| Gender (% Male) | 48.9 | 46.2 | 57.1 | 68.7 | 45.0 | 57.2 | 60.7 |
| Age (Mean (Std)) | 57.0 (15.9) | 61.1 (16.8) | 59.0 (15.4) | 65.2 (13.8) | 56.5 (20.0) | 63.6 (16.6) | 67.6 (15.4) |
| Ethnic (% A/B/W/O) | 16.7/4.3/54.1/24.9 | 1.1/7.5/23.1/68.3 | - | - | 16.5/6.4/42.2/34.9 | - | - |
| LoS (Median hours (IQR)) | 49 (24, 84) | 10 (7, 15) | 25 (20, 46) | 160 (57, 332) | 7 (6, 9) | 43 (20, 122) | 44 (21, 96) |
| **Patient stay prevalence** (num. stays (%)) | | | | | | | |
| Mortality | 233 (0.5) | 48 (0.0) | 134 (1.0) | 3,470 (25.6) | - | 14,501 (4.7) | 3533 (5.7) |
| Circ. Failure | 535 (1.2) | - | 1,207 (9.1) | 5,509 (40.6) | 613 (0.8) | 78,174 (25.6) | 17,099 (27.8) |
| Sepsis-3 | - | - | - | 5,161 (38.0) | - | - | - |
| Hyperglyc. | 4,440 (10.4) | - | 4216 (31.8) | 3,081 (22.7) | 7,351 (9.4) | 123,351 (40.4) | 22068 (35.9) |
| Resp. Failure | 424 (1.0) | - | 515 (3.9) | 3,860 (28.5) | - | 39,308 (12.9) | 4532 (7.4) |
| Kidney Failure | - | - | 740 (5.6) | 1,750 (12.9) | - | 34,998 (11.5) | 5552 (9.0) |
| Liver Failure | - | - | 299 (2.3) | 420 (3.1) | 618 (0.8) | 9914 (3.2) | 1157 (1.9) |
| **Time-step label prevalence** (positive %, (negative %, unknown %)) | | | | | | | |
| Decomp. 24h | 0.2 (99.8, 0.0) | 0.0 (100.0, 0.0) | 0.5 (99.5, 0.0) | 3.5 (96.5, 0.0) | - | 1.1 (98.9, 0.0) | 1.6 (98.4, 0.0) |
| Circ. Fail. 8h | 0.3 (9.5, 90.2) | - | 2.0 (28.9, 69.1) | 2.7 (46.4, 50.9) | 0.3 (20.2, 79.5) | 3.3 (60.7, 35.9) | 3.8 (41.0, 55.2) |
| Sepsis-3 8h | - | - | - | 0.8 (59.0, 40.2) | - | - | - |
| Hyperglyc. 8h | 4.8 (18.1, 77.1) | - | 13.7 (33.2, 53.1) | 4.3 (14.7, 81.0) | 2.4 (20.6, 77.0) | 14.5 (64.3, 21.2) | 13.0 (50.8, 36.2) |
| Resp. Fail. 24h | 0.5 (2.0, 97.5) | - | 2.5 (22.6, 74.9) | 6.5 (25.0, 68.5) | - | 5.5 (40.7, 53.8) | 3.5 (18.0, 78.5) |
| Kidney Fail. 48h | - | - | 2.2 (14.0, 83.8) | 2.7 (46.9, 50.4) | - | 2.7 (57.6, 39.7) | 3.9 (58.3, 37.8) |
| Liver Fail. 48h | - | - | 2.4 (46.7, 50.9) | 1.7 (34.9, 63.4) | 0.1 (8.2, 91.7) | 3.2 (47.1, 49.7) | 1.7 (70.4, 27.9) |
| **Data density** (Median % (IQR %)) | | | | | | | |
| Vital Density | 1.9 (1.0, 3.4) | 10.3 (8.0, 13.3) | 25.6 (12.4, 27.6) | 20.6 (8.7, 30.9) | 13.8 (10.3, 17.2) | 28.1 (22.1, 35.3) | 21.6 (13.9, 27.2) |
| Lab Density | 0.6 (0.2, 1.3) | 0.0 (0.0, 0.0) | 5.0 (3.0, 7.3) | 2.5 (1.1, 4.7) | 4.8 (3.7, 6.1) | 8.6 (4.6, 11.8) | 6.0 (3.2, 8.3) |
| Vital Density adj. | 3.8 (1.8, 6.7) | 42.9 (33.3, 55.1) | 33.7 (16.3, 36.4) | 27.1 (11.5, 40.7) | 57.1 (42.9, 71.4) | 28.1 (22.1, 35.3) | 22.4 (14.4, 28.2) |
| Lab Density adj. | 0.7 (0.2, 1.4) | 0.0 (0.0, 0.0) | 7.2 (4.3, 10.3) | 2.7 (1.1, 4.9) | 6.2 (4.8, 7.9) | 9.1 (4.8, 12.5) | 6.2 (3.3, 8.6) |

**Table S2:** Dataset statistics showing demographic information, patient prevalence for different downstream tasks, time-step prevalence for specific early event prediction labels of each downstream task, data densities for vitals and laboratory tests (regularly and irregularly measured variables, respectively). Adjusted densities (adj.) exclude variables that are not observed for a specific dataset. Statistics for further harmonized data sources are summarized in Table S1. Ethnicity is encoded as A: Asian, B: Black, W: White, O: Other/Unknown.

**MIMIC-IV** The MIMIC-IV dataset represents a major update to MIMIC-III. It contains only data from the period after introducing the iMDsoft Metavision ICU data management system in 2008 and in version v3.1 contains patients up to the year 2022 (including the COVID-19 period). There is a non-trivial overlap of patients between the MIMIC-III and MIMIC-IV datasets. This overlap could be mostly removed by only considering patient data stored in the CareVue system, when using data from MIMIC-III, if this is desirable. We extract individual intensive care stays from this dataset, while the dataset could support extracting entire hospital admissions potentially including multiple subsequent ICU stays.

**eICU** The "eICU Collaborative Research Database" is the largest dataset in the collection and contains patient data from 207 hospitals across the United States. It contains data from 2015 and 2016. The provided data density is comparatively lower, but the data covers a broad range of measurements and facilitates accurate treatment rate extractions, large set of laboratory test results, and ventilation settings amongst other data. We extract individual intensive care stays from this dataset, while the dataset could support extracting entire hospital admissions potentially including multiple subsequent ICU stays.

**NW-ICU** The "Northwestern ICU (NWICU) database" is a multi-center dataset containing data from 2020–2022 spanning the COVID-19 pandemic period. The dataset has lower data densities compared to, e.g., the MIMIC-III/IV datasets and we found it more challenging to retrieve accurate treatment information from the dataset. Surprisingly, in its current version (v0.1.0), we were not able to retrieve comprehensive ventilation information from the dataset, which would be highly relevant to accurately study COVID-19 patient populations. We extract individual intensive care stays from this dataset, while the dataset could support extracting entire hospital admissions potentially including multiple subsequent ICU stays.

**HiRID** The "HiRID, a high time-resolution ICU dataset" is a high-quality intensive care dataset containing data from 2008–2016. The data contains highly dense vital measurements and laboratory test results, accurate reporting of treatment administration and ventilation settings. We extract individual intensive care stays from this dataset. The dataset does not support associating multiple subsequent stays with the same patient.

**UMCdb** The "AmsterdamUMCdb" is a high-quality intensive care dataset containing data from 2003–2016. The data contains highly dense vital measurements and laboratory test results, accurate reporting of treatment administration and ventilation settings. We extract individual intensive care stays from this dataset. The dataset does not support associating multiple subsequent stays with the same patient.

**SICdb** The “Salzburg Intensive Care database (SICdb)” is a high-quality intensive care dataset containing data from 2013–2021. The data contains highly dense vital measurements and laboratory test results, accurate reporting of treatment administration and ventilation settings. The dataset shares the highest frequency for vitals across all datasets. Vitals can be extracted to minute-level updates. We extract individual intensive care stays from this dataset. The dataset does not support associating multiple subsequent stays with the same patient.

**PICdb** The “Pediatric Intensive Care database” is a Chinese pediatric care dataset and hence, except for a few children in the MIMIC-III dataset, the only dataset in the harmonized collection containing children or newborns. It contains data from 2010–2018. The measurement density on this dataset is relatively low. Treatments are mostly extracted only as indicators due to a lack of administration rate information. We extract individual intensive care stays from this dataset, while the dataset could support extracting entire hospital admissions potentially including multiple subsequent ICU stays.

**Zigong EHR** The “Critical care database comprising patients with infection at Zigong Fourth People’s Hospital” is a small dataset focused on infectious diseases and hence a patient population with more severe outcomes compared to the other datasets in the collection. It contains data from 2019 and 2020. We extract individual intensive care stays from the dataset, while the dataset could support extracting entire hospital admissions potentially including multiple subsequent ICU stays. Data recording density is also much lower, which makes this dataset more challenging both to accurately annotate and for the machine learning models to perform well. Treatments are mostly extracted only as indicators due to a lack of administration rate information.

**INSPIRE** The dataset contains a cohort of surgical patients from a South Korean hospital including associated intensive care stays from 2011–2020. We extract the individual stays in intensive care after a surgery (excluding the time spent in surgery) and split longer stays with multiple surgeries, hence each stay is identified by a surgery and its post-surgical ICU stay either until the next surgery, discharge, or death. The cohort is comparatively much healthier considering the prevalences of the various annotated failure events (Table S1 and Table S2).

**EHRSHOT** This dataset contains data from Stanford medicine from 1990–2023. It has not been built with the primary objective to share high frequency data from intensive care such as continuous vital measurements. It contains mostly coded data points recorded for billing and insurance purposes. However, after closer inspection, we found it contains a considerable amount of coded vital, lab, and treatment measurements paired with the actually observed numeric values. We harmonize the data source and develop a heuristic (Supplementary A.2.1) to extract more general hospital stays, which might include periods of intensive care stays, but do not need to. The extracted stays are hence much sparser in available information and represent a comparatively healthier patient population.

**MIMIC-IV ED** This dataset contains emergency department admissions at the Beth Israel Deaconess Medical Center from 2011–2019. The ED cohort is very heterogeneous, with varying disease severities. We extract sparse vital information from the datasets `triage` and `vitalsigns` tables. There is no continuous treatment data available, but we extract treatment indicators based on the information in the `pyxis` table. We additionally merge in missing demographic information from the core MIMIC-IV dataset, where possible.

**MC-MED** The “Multimodal Clinical Monitoring in the Emergency Department” dataset contains a diverse cohort of adult ED visits from Stanford Medical Center spanning the COVID-19 pandemic period (September 2020 to September 2022). With over a hundred thousand monitored ED visits, it is the first ED dataset to include continuously recorded physiological waveforms (electrocardiogram, photoplethysmogram, and respiration) alongside vital signs, in addition to clinical data such as demographics, medical histories, laboratory and imaging results, medication administrations, orders, and visit outcomes. The continuous monitoring and physiological waveforms distinguish MC-MED from existing ED datasets, such as MIMIC-IV-ED which contains only infrequent vital sign measurements. This emergency department dataset hence has higher data density than MIMIC-IV ED. We extract individual emergency department visits from this dataset.

**PSSS** The “Personalized Swiss Sepsis Study” is an unpublished dataset collected from the five Swiss university hospitals located in the cities of Basel (USB, University Hospital of Basel), Bern (Inselspital, University Hospital of Bern), Geneva (HUG, University Hospital of Geneva), Lausanne (CHUV, University Hospital of Lausanne), and Zurich (USZ, University Hospital of Zurich). The data was collected with the primary objective to advance early detection of sepsis, but also provides rich data for general ICU monitoring applications with a broad set of recorded and harmonized vital signs, laboratory test results, as well as treatment and life support device monitoring data. The data from the five hospitals was collected from 2019 until 2022.

**Charité** The Charité dataset is an unpublished dataset consisting of routinely collected intensive care data from Charité – Universitätsmedizin Berlin, Germany. Clinical care at Charité – Universitätsmedizin Berlin is delivered across three campuses (Campus Charité Mitte, Campus Virchow-Klinikum, and Campus Benjamin Franklin) in Berlin. The dataset contains comparatively dense longitudinal information on vital signs, laboratory measurements, medications, and organ support therapies from multiple critical care patient data management systems and hospital information systems (mainly, COPRA5 and COPRA6, SAP IS-H / i.s.h.med), spanning the years 2006–2025 across all sites. Initial inclusion criteria comprise all adult patients ($\geq$18 years) admitted to intensive care units (see below for further selection). We preprocessed the data by harmonizing records across heterogeneous source systems (COPRA5/6, SAP IS-H), aligning measurements temporally, and standardizing units and nomenclature.

**RBKICU** The RBKICU dataset contains intensive care data collected at Robert Bosch Krankenhaus in Stuttgart, Germany, from 2012 to 2024. The data were recorded at a resolution of up to 1 minute and include vital signs, laboratory measurements, medications, and organ support therapies. The dataset was collected primarily to study and predict acute kidney injury.

##### A.2.1 EHRSHOT data extraction details

To extract valid patient stays from the EHRSHOT dataset, a multi-step heuristic was applied to clean and consolidate visit records from the original long-format table provided by the authors. First, rows corresponding to visit information (i.e., those with `omop_table` equal to `visit_occurrence` or `visit_detail`) were extracted, yielding over one million candidate stays. Stays with zero duration (i.e., where the start and end times were identical) were removed. For each unique combination of patient and visit identifier (`visit_id`), an overarching stay was defined using the earliest start and latest end times among all visits with the same identifier. If only one visit was found for a given overarching stay, it was retained unchanged; otherwise, a new consolidated stay was created to span the full time window, with a merged list of codes from all constituent entries. Then, for each patient, visits that were fully contained within another were removed, but only after their codes were merged into the encompassing stay. Events associated with these removed visits were not discarded: they were re-mapped to the appropriate merged stay based on timestamp overlap. Finally, temporally adjacent or overlapping visits were further merged into composite stays, with unified time windows and concatenated codes. This process yielded a final set of 118,898 non-overlapping, chronologically ordered stays across 5,900 patients. These validated stays were then used to map all relevant events, including special handling of death records, producing a clean, gap-free timeline of patient visits suitable for downstream clinical modeling and analysis.

##### A.2.2 Inclusion criteria and data splitting

We consider patient stays that after extraction have

- a valid admission and discharge time,
- a valid length of stay (LoS) of at least 4 hours,
- a maximum gap between measurements of at most 48 hours,
- and at least 4 measurements.

Compared to Van De Water et al.[16], we broaden the inclusion criteria by reducing the LoS requirement from 6 to 4 hours and increasing the allowed maximum gap between measurements from 12 to 48 hours.

We keep inclusion criteria minimal so the pretraining corpus covers a broad demographic; the resulting dataset can be further trimmed for specific downstream studies.

In Figure S1 we visualize the cohort creation flow through the aforementioned inclusion criteria and the anonymization procedure (see Supplementary A.6). Note that a large portion of stays lost when excluding admissions with length of stay below 4 hours come from MIMIC-IV *ED*. Similarly, due to completely missing demographic information for a large portion of admissions in MIMIC-IV *ED* these also got excluded in the anonymization procedure.

Finally, for the purpose of machine learning experiments each dataset was split (by patients if applicable and individual stays otherwise) into a train, validation, and test set of patients. The train and validation datasets were used to train (parametric optimization using gradient descent) and validate (hyperparameter and general model selection) the models and the test sets to report performance.

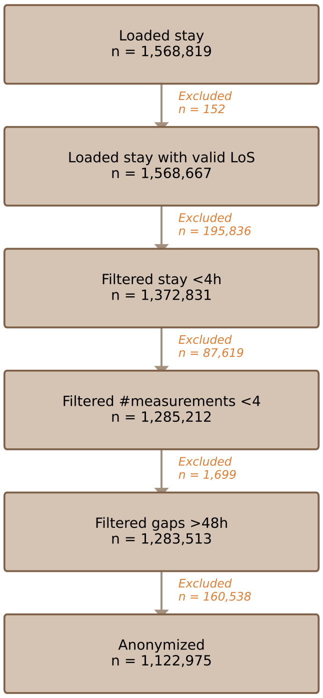


**Figure S1:** Cohort creation flow

###### A.2.3 Data leakage considerations

In our study we perform multiple pretrainings of ICareFM and every time hold out an entire dataset. Most relevant in the case of MIMIC-III and MIMIC-IV we only provide evaluation results on MIMIC-IV. When pretraining to evaluate on MIMIC-IV, we completely remove both MIMIC-III and MIMIC-IV from the pretraining. When pretraining for any other non-MIMIC dataset, we use patient data from both MIMIC-III and MIMIC-IV. For task zero-shot results with domain-adapted pretraining (i.e., the target dataset was included in pretraining but no task-specific labels were used), we excluded MIMIC-III to avoid leakage from MIMIC-III patients into the in-domain MIMIC-IV test set.

For multi-center datasets such as eICU, which comprises multiple hospitals, the entire dataset is held out as one unit; this represents a conservative evaluation, as the held-out data spans a broad range of institutions.

Further we have two datasets containing data from Inselspital Bern in Switzerland. One is the publicly available dataset HiRID[10] (data until 2016) and the second source is in the private PSSS (Personalized Swiss Sepsis Study) collection, where data from Inselspital Bern was collected from 2019 - 2022. We exclude the entire PSSS data for pretraining when evaluating the model on HiRID. This avoids both same hospital but in this case also same country data leakage. Note that for a few selected results (e.g. Figure S23), a model including data from HiRID was evaluated on the PSSS hospitals. Note that in this case the evaluation represents a domain (hospital shift) for the hospitals of Basel, Geneva, Lausanne, and Zurich, but only a temporal shift with a gap of 3 years in between. This is often reflected in the results where ICareFM tends to transfer well if historical data from the same hospital was part of the pretraining corpus.

###### A.2.4 Intensive Care data types

Intensive care data relevant for timely predictions and harmonized in our dataset can be grouped into the following data types:

- Demographic patient information, which is data considered to remain constant during the time period of an intensive care stay such as gender, age, height, etc. Some of these variables can be categorical such as ethnicity (not available in all datasets).
- Numeric physiological vital data measured by various equipment such as heart rate, oxygen saturation or data measured by ventilators such as plateau pressure, peak pressure, etc. This data is observed continuously and also stored with high frequency and density in most of the harmonized datasets.
- Categorical data recorded by equipment or entered manually by nursing staff such as Glasgow Coma Scale score or RASS scores. Human derived scores are available more sparsely and availability also varies across hospitals depending on nursing staff policies.

- Numeric laboratory test results from blood or other body fluid samples. These are available sparsely and the data density varies across hospitals due to different patient population needs, hospital policies on regular monitoring activities, and financial trade-offs.
- Treatments:
  - Life support equipment to support or take over functions of failing organs: Ventilation settings or CRRT (continuous renal replacement therapy) settings.
  - Drugs given via infusions, we extract both accurate treatment administration rates and treatment indicators (binary presence of active infusion).
  - Drugs given via other pathways as bolus form (injections, tablets, etc.). If dosage information is available we harmonize these drug administrations together with the infusions by computing an instantaneous application rate, otherwise a simple administration indicator is extracted.

##### A.2.5 Concepts for treatment variables

To create a set of treatment concepts, we select variables that were identified as most predictive for various labels in published literature (circulatory failure [17], respiratory failure [18], kidney failure [19], sepsis [20]). Then, based on the input from consulting ICU clinicians, some high-priority concepts are made more granular (e.g., vasopressors) while the low priority ones are left out (e.g., laxatives).

In about half of the datasets, treatment variables are stored as plain text, amounting to over 16,000 unique entries. Manual mapping of that number of variables would be slow and error-prone. Instead, we first use LLMs to pre-label the variables according to the provided concept set. This allows grouping of similar treatments, making manual verification much quicker. The manual verification is then performed in three rounds: first by data scientists, and then independently by two physicians, ensuring the mapping is correct.

Previous work by Oliver et al. [21] has proposed a processing pipeline for harmonizing treatments by including drug exposure information as indicators. We expand on this by (1) considering not only indicators, but also administration rates for core medications used in critical care settings, and (2) grouping individual drugs into abstract treatment concepts, thereby increasing the overlap across datasets in concepts while maintaining relevance for downstream applications.

More generally, previous work [20] has suggested that including medication variables harms the accuracy of predictive models when transferring models due to changes in treatment policies [18], but little research has been done on the reasons behind this effect and what can be done to mitigate it. The information about administered medications is an insight into the actions of the clinicians, and could drastically improve the model accuracy and transfer if the model can learn proper causal treatment effects. By including these variables in the harmonization pipeline, we prepared the ground for this future research on learning generalizable treatment effect estimations on critical care time series.

#### A.3 Concept reference table

The full concept (or variable) reference table is shown in Table S3. It includes 130 variables: 6 static demographic, 81 observations, and 43 treatment variables.

| Tag | Name | Type | Organ System | Unit |
|---|---|---|---|---|
| map | Mean Arterial Blood Pressure | observation | circulatory | mmHg |
| lact | Lactate | observation | circulatory | mmol/L |
| age | Age | demographic | None | years |
| weight | Weight | demographic | None | kg |
| sex | Sex | demographic | None | categorical |
| height | Height | demographic | None | cm |
| hr | Heart Rate | observation | circulatory | bpm |
| fio2 | FiO2 | observation | respiratory | % |
| resp | Respiratory Rate | observation | respiratory | insp/min |
| temp | Temperature | observation | infection | C |
| crea | Creatinine | observation | metabolic_renal | mg/dL |
| urine_rate | Urine Rate Per Hour | observation | metabolic_renal | mL/h |
| po2 | Partial Pressure Of Oxygen | observation | respiratory | mmHg |
| ethnic | Ethnic Group | demographic | None | categorical |
| alb | Albumin | observation | gastrointestinal | g/dL |

| | | | | |
|---|---|---|---|---|
| alp | Alkaline Phosphatase | observation | gastrointestinal | IU/L |
| alt | Alanine Aminotransferase | observation | gastrointestinal | IU/L |
| ast | Aspartate Aminotransferase | observation | gastrointestinal | IU/L |
| be | Base Excess | observation | metabolic_renal | mmol/l |
| bicar | Bicarbonate | observation | metabolic_renal | mmol/l |
| bili | Total Bilirubin | observation | gastrointestinal | mg/dL |
| bili_dir | Bilirubin Direct | observation | gastrointestinal | mg/dL |
| bnd | Band Form Neutrophils | observation | infection | % |
| bun | Blood Urea Nitrogen | observation | metabolic_renal | mg/dL |
| ca | Calcium | observation | metabolic_renal | mg/dL |
| cai | Calcium Ionized | observation | metabolic_renal | mmol/L |
| ck | Creatine Kinase | observation | circulatory | IU/L |
| ckmb | Creatine Kinase MB | observation | circulatory | ng/mL |
| cl | Chloride | observation | metabolic_renal | mmol/l |
| crp | C-Reactive Protein | observation | infection | mg/L |
| dbp | Diastolic Blood Pressure | observation | circulatory | mmHg |
| fgn | Fibrinogen | observation | circulatory | mg/dL |
| glu | Glucose | observation | metabolic_renal | mg/dL |
| hgb | Hemoglobin | observation | circulatory | g/dL |
| inr_pt | Prothrombin | observation | circulatory | INR |
| k | Potassium | observation | metabolic_renal | mmol/l |
| lymph | Lymphocytes | observation | infection | % |
| methb | Methemoglobin | observation | circulatory | % |
| mg | Magnesium | observation | metabolic_renal | mg/dL |
| na | Sodium | observation | metabolic_renal | mmol/l |
| neut | Neutrophils | observation | infection | % |
| pco2 | CO2 Partial Pressure | observation | respiratory | mmHg |
| ph | pH Of Blood | observation | metabolic_renal | pH |
| phos | Phosphate | observation | metabolic_renal | mg/dL |
| plt | Platelet Count | observation | circulatory | G/l |
| ptt | Partial Thromboplastin Time | observation | circulatory | sec |
| sbp | Systolic Blood Pressure | observation | circulatory | mmHg |
| tnt | Troponin T | observation | circulatory | ng/mL |
| wbc | White Blood Cell Count | observation | infection | G/l |
| basos | Basophils | observation | infection | % |
| eos | Eosinophils | observation | infection | % |
| mgcs | Glasgow Coma Scale Motor | observation | neuro | categorical |
| tgcs | Glasgow Coma Scale Total | observation | neuro | categorical |
| vgcs | Glasgow Coma Scale Verbal | observation | neuro | categorical |
| egcs | Glasgow Coma Scale Eye | observation | neuro | categorical |
| hct | Hematocrit | observation | circulatory | % |
| rbc | Red Blood Cell Count | observation | circulatory | m/uL |
| tri | Troponin I | observation | circulatory | ng/mL |
| etco2 | Endtidal CO2 | observation | respiratory | mmHg |
| rass | Richmond Agitation Sedation Scale | observation | neuro | categorical |
| hbco | Carboxyhemoglobin | observation | circulatory | % |
| esr | Erythrocyte Sedimentation Rate | observation | infection | mm/hr |
| pt | Prothrombine Time | observation | circulatory | sec |
| adm | Patient Admission Type | demographic | None | categorical |
| hba1c | Hemoglobin A1C | observation | metabolic_renal | % |
| samp | Body Fluid Sampling, Detected Bacterial Growth | observation | infection | categorical |
| spo2 | Pulse Oxymetry Oxygen Saturation | observation | respiratory | % |
| sao2 | Oxygen Saturation In Arterial Blood | observation | respiratory | % |
| icp | Intra Cranial Pressure | observation | neuro | mmHg |
| cout | Cardiac Output | observation | circulatory | l/min |
| mpap | Mean Pulmonal Arterial Pressure | observation | circulatory | mmHg |
| spap | Systolic Pulmonal Arterial Pressure | observation | circulatory | mmHg |
| dpap | Diastolic Pulmonal Arterial Pressure | observation | circulatory | mmHg |
| cvp | Central Venous Pressure | observation | circulatory | mmHg |
| svo2 | Mixed Venous Oxygenation | observation | circulatory | % |
| pcwp | Pulmonary Capillary Wedge Pressure | observation | circulatory | mmHg |
| peep | Positive End Expiratory Pressure - Mechanical Ventilation | observation | respiratory | cmH2O |
| peak | Peak Pressure - Mechanical Ventilation | observation | respiratory | cmH2O |
| plateau | Plateau Pressure - Mechanical Ventilation | observation | respiratory | cmH2O |
| ps | Pressure Support - Mechanical Ventilation | observation | respiratory | cmH2O |
| tv | Tidal Volume | observation | respiratory | ml |
| airway | Type Of Airway Ventilation | observation | respiratory | categorical |
| supp_o2_vent | Supplemental Oxygen From Ventilator | treatment | respiratory | % |
| ygt | Gamma GT | observation | gastrointestinal | U/L |
| amm | Ammonia | observation | gastrointestinal | umol/L |

| | | | | |
|---|---|---|---|---|
| amyl | Amylase | observation | gastrointestinal | U/L |
| lip | Lipase | observation | gastrointestinal | U/L |
| ufilt | Ultrafiltration On Continuous RRT | treatment | metabolic_renal | ml |
| ufilt_ind | Ultrafiltration On Continuous RRT Indicator | treatment | metabolic_renal | indicator |
| dobu | Dobutamine | treatment | circulatory | mcg/min |
| levo | Levosimendan | treatment | circulatory | mcg/min |
| norepi | Norepinephrine | treatment | circulatory | mcg/min |
| epi | Epinephrine | treatment | circulatory | mcg/min |
| milrin | Milrinone | treatment | circulatory | mcg/min |
| teophyllin | Theophylline | treatment | circulatory | mg/min |
| dopa | Dopamine | treatment | circulatory | mcg/min |
| adh | Vasopressin | treatment | circulatory | U/min |
| hep | Heparin | treatment | circulatory | U/h |
| prop | Propofol | treatment | neuro | mcg/min |
| benzdia | Benzodiazepine | treatment | neuro | mg/h |
| sed | Other Sedatives | treatment | neuro | indicator |
| op_pain | Opiate Painkiller | treatment | neuro | indicator |
| nonop_pain | Non-Opioid Analgesic | treatment | neuro | indicator |
| paral | Paralytic | treatment | neuro | indicator |
| abx | Antibiotics | treatment | infection | indicator |
| loop_diur | Loop Diuretic | treatment | metabolic_renal | mg/h |
| ins_ind | Insulin | treatment | None | indicator |
| fluid | Fluid Administration | treatment | None | indicator |
| inf_rbc | Packed Red Blood Cells | treatment | None | indicator |
| ffp | Fresh Frozen Plasma | treatment | None | indicator |
| plat | Platelets | treatment | None | indicator |
| inf_alb | Albumin Infusion | treatment | None | indicator |
| anti_delir | Anti Deliriant | treatment | neuro | indicator |
| oth_diur | Other Diuretics | treatment | metabolic_renal | indicator |
| anti_coag | Other Anticoagulants | treatment | circulatory | indicator |
| vasod | Antihypertensive And Vasodilators | treatment | circulatory | indicator |
| anti_arrhythm | Antiarrhythmic | treatment | circulatory | indicator |
| dobu_ind | Dobutamine Indicator | treatment | circulatory | indicator |
| levo_ind | Levosimendan Indicator | treatment | circulatory | indicator |
| norepi_ind | Norepinephrine Indicator | treatment | circulatory | indicator |
| epi_ind | Epinephrine Indicator | treatment | circulatory | indicator |
| milrin_ind | Milrinone Indicator | treatment | circulatory | indicator |
| teophyllin_ind | Theophylline Indicator | treatment | circulatory | indicator |
| dopa_ind | Dopamine Indicator | treatment | circulatory | indicator |
| adh_ind | Vasopressin Indicator | treatment | circulatory | indicator |
| hep_ind | Heparin Indicator | treatment | circulatory | indicator |
| prop_ind | Propofol Indicator | treatment | neuro | indicator |
| benzdia_ind | Benzodiazepine Indicator | treatment | neuro | indicator |
| loop_diur_ind | Loop Diuretic Indicator | treatment | metabolic_renal | indicator |

**Table S3:** Concept reference

#### A.4 Data processing

##### A.4.1 Harmonized time series extraction

We extract harmonized time series by performing three key steps: 1) concept harmonization 2) outlier removal 3) aggregation into bins across time and different sources into a harmonized sparse hourly grid. The pipeline starts by collecting all data points for a given patient referring to a specific concept. These data points might be sourced from different tables, columns, source data identifiers, formats, units, etc., and require harmonization to standardize them into a unified data type with a fixed unit for the target concept. The collection of data points for the target concept is then filtered for outliers to remove physiologically implausible values. Note that the ranges of allowed values are typically chosen with big margins, to avoid erroneously removing strong signals of severely ill patients. The filtered data is then binned to an hourly sparse time grid, where for numerical data median aggregation is performed, for categoricals the mode is extracted, and for binary information an *any* operation is used. For continuous treatment data (infusion rates) the rate updates are extracted as accurately as possible given the available data w.r.t. time and amount and mean aggregation is used for temporal binning. No imputation is performed at this stage, time bins without any data remain empty.

##### A.4.2 Scaling

Data is scaled depending on its type, scaling parameters are fitted on ground truth data alone:

- continuous observations are standardized (i.e. centered and scaled to unit variance) and if deemed suitable a log transformation is applied before standardization,
- categorical observations are one-hot encoded and each variable has a dedicated class to encode missing information,
- continuous treatments are quantile-transformed and mapped to the $[0, 1]$ range such that a $0$ represents *no medication given*,
- treatment indicators are binary encoded using $\{0, 1\}$ where $0$ represents *no medication given*.

##### A.4.3 Imputation

Gridded time-step data as inputs for model training are forward-filled indefinitely for all observation variables. The remaining missing values are then imputed with $0$ for continuous variables, which corresponds to a population mean imputation after considering standard scaling before the imputation stage. The remaining categorical entries are imputed with a value corresponding to the dedicated class that encodes missing information for each categorical variable.

Any treatment variable is excluded from forward-filling operations and missing data points are strictly filled using $0$, which given the previously introduced scaling and encoding scheme always corresponds to no treatment being applied.

##### A.4.4 Feature extraction

We build on the feature set proposed by Soenksen et al.[22] to process the MIMIC-IV[6] dataset. To improve performance, we then further expand this set of features and select specific features for each variable type. For each time step, each feature is computed over three history sizes of 8, 24, and 72 hours:

- For continuous observations and continuous treatment variables, we compute:
  - mean on raw and imputed data,
  - standard deviation on raw data,
  - slope of a linear fit on the raw data and imputed data,
  - mean absolute change over imputed data,
  - fraction of non-missing data points,
  - quantiles: 0% (Min.), 10%, 50%, 90%, 100% (Max.).
- For categorical variables, we compute the mode, number of missing points, and a binary indicator of whether there are any missing points at all.
- For treatment indicators we compute the number of points with treatment and a binary indicator whether any treatment was applied.

#### A.5 Task annotations

We define sample labels by annotating the time series following the clinical definitions used by prior work for each specific task. As such, these labels constitute proxy labels derived and computed from data given a clinical definition to diagnose a patient state for a certain condition. Most importantly, we annotate a positive and a negative case only if there's enough evidence in favor of either. As such, a label is only computed if the source data, conditioned on a task-specific imputation scheme proposed by prior work, provides all required inputs to compute the score or state annotation of the clinical task definition at a certain time step. Based on these cases, we define early event prediction labels (e.g., respiratory failure) that are then used for online classification of the future state of the patient.

##### A.5.1 Event Annotations

Events are diagnostically annotated based on available data using diagnostic criteria and scoring functions. Given a patient time-series resampled to a 1-hour grid, we initialize the event annotation vector with unknown states.

**Circulatory Failure** We follow Hyland et al.[17] relying on mean arterial pressure (at 65 mmHg) and blood serum lactate (at 2 mmol/l) measurements to annotate circulatory failure events. We apply the lactate imputation scheme as proposed by Hyland et al.[17] to create a denser availability of the patient's possible lactate measurement. The imputation scheme assumes less frequent measurements for stable patients and as such indefinitely forward and backward fills those measurements. Critical measurements above the mentioned threshold are only forward and backward filled up to 6 hours. If two measurements are closer than 6 hours the values in between are linearly interpolated. Mean arterial pressure is assumed to be typically recorded with high frequency on almost all considered datasets, and as such no further imputation scheme is applied, following Hyland et al.[17]. A positive event is annotated if observed mean arterial pressure is below 65 mmHg and imputed lactate is above 2 mmol/l. A negative event is annotated if observed mean arterial pressure is above 65 mmHg (without treatment) and imputed lactate is below 2 mmol/l. Time-steps with any missing information for mean arterial pressure or lactate after imputation or for which neither of the two conditions matches the data, are annotated with an unknown current patient state.

To construct a continuous severity gradient for circulatory failure, we employed lactate as a univariate severity indicator while maintaining the hemodynamic criterion (MAP < 65 mmHg or vasopressive drugs) as a fixed qualifying condition. This approach leverages the established role of lactate as a marker of tissue hypoperfusion, with escalating concentrations reflecting progressive severity of circulatory compromise. We defined five severity strata using lactate thresholds of 1.5, 2.0[17], 3.0, 4.0, and 5.0 mmol/L.

**Respiratory Failure** A patient's respiratory state can be assessed by calculating the ratio of partial pressure of oxygen in blood (PaO2) over the fraction of inspired oxygen (FiO2, 21% in ambient air or higher during ventilation)[18], short the P/F ratio (also known as Horovitz index). Hüser et al.[18] proposed an intricate imputation scheme with a parametric imputation algorithm to obtain a dense approximation of the patients PaO2 measurements. Alternatively, Yèche et al.[23] rely on a simple rule-based algorithm proposed by Ellis et al.[24] to estimate PaO2 from densely available pulse oxymetry oxygen saturation (SpO2) measurements. Due to the heterogeneous nature of large-scale harmonized multi-source datasets, we decided to use minimal assumptions and not use any form of rule-based or parametric imputation algorithm for PaO2. We backward and forward fill both FiO2 and PaO2 measurements each by at most 1 hour (essentially 1 time-step on the hourly grid). We aim to predict severe respiratory failure which is defined as a P/F ratio below 100 mmHg[18]. If after imputation both PaO2 and FiO2 information are available we annotate a positive event for P/F below 100 mmHg and a negative event for P/F over 100 mmHg, time points with missing data are annotated as unknown event state. Note that we do not make any assumption on the patient's FiO2 measurements outside the 1 hour imputation window around ground truth data. We do not impute missing data points with a 21%, which would correspond to ambient air assuming no ventilation. While the harmonization efforts aim to extract ventilation information, we could not reliably extract dense ventilation information from each dataset and as such do not impose any assumptions here, which could falsely impute ambient air inspired oxygen concentration while the patient is actually on a ventilator but with unreliable data recording and export. This decision causes the label to only annotate events for ventilated patients where FiO2 measurements are recorded by the used ventilation equipment. We do believe the task maintains high clinical relevance while relying largely only on ground truth data for annotations.

To evaluate model performance across a spectrum of respiratory compromise, we employed P/F ratio thresholds of 200, 180, 160, 140, 120, and 100 mmHg as severity indicators. These thresholds span the range from mild to severe acute respiratory distress syndrome according to the Berlin definition[25], enabling assessment of model discrimination across clinically distinct severity strata. The 100 mmHg threshold represents severe respiratory failure with high mortality risk, while the 200 mmHg threshold captures moderate respiratory failure.

**Kidney Failure** We annotate most severe form (stage 3) of acute kidney injury as defined by the *Kidney Disease: Improving Global Outcomes* (KDIGO) guidelines[26]. Based on work by Lyu et al.[19], we linearly interpolate missing creatinine measurements between ground truth measurements if they are closer than 48 hours. Borders are backward and forward filled with a maximum horizon of 48 hours. Lyu et al.[19] backward filled missing urine information indefinitely. Due to the heterogeneity in the harmonized multi-center dataset we decided to backward fill urine rates only up to 48 hours at most. After imputation the KDIGO AKI Stage

3 definitions are applied to annotate events. To summarize, a patient is in stage 3 AKI if on renal replacement therapy, creatinine values have increased drastically (either absolutely measured or w.r.t. patient's determined baseline values), or the patient has little to no urine output over prolonged period of time. If any of these conditions can be confirmed from the imputed data, the time step is annotated with a positive event. If both urine and creatinine state can be determined from the aforementioned imputed data and are assessed to be stable, the event annotation is negative. All other time steps remain with an unknown event annotation.

To modulate severity we consider the three levels of severity as proposed by the KDIGO guidelines with AKI stage 1, 2, and 3.

**Liver Failure** To derive an early event prediction task related to liver failure we use the model for end-stage liver disease (MELD)[27]. The score is a linear combination over the logarithms of creatinine, total bilirubin, and the international normalized ratio (INR) all as measured from blood samples. We choose a threshold of 30 for the score to detect rare and severe forms of liver disease. According to Wiesner et al.[28] this threshold corresponds to a 50% expected mortality on their study cohort. We minimally impute the three blood sample values by backward and forward filling for at most 1 hour (implying at most 1 step of imputation in either temporal direction) and then compute the score where all three measurements are available. A positive event is annotated for time steps with available score above 30 and a negative event is annotated for time steps with available scores below 30.

To assess model performance across varying degrees of hepatic dysfunction, we employed MELD score thresholds of 15, 20, 25, 30, and 35. These thresholds encompass a clinically relevant severity spectrum: scores of 10-19 represent moderate liver disease with anticipated 3-month mortality of approximately 6%, scores of 20-29 indicate high-risk patients with 20% 3-month mortality, and scores $\geq$30 represent severe end-stage liver disease with mortality exceeding 50%[28].

**Hyperglycemia** We incorporate an additional simple early event prediction task, which requires minimal assumptions compared to other tasks relying on imputation schemes and heuristics. Based on work by Mehdizavareh et al.[29] we propose to perform the early event prediction of elevated blood glucose measurements. We do not apply any form of imputation. A positive hyperglycemic event is annotated where a ground truth glucose measurement is available and above 180 mg/dL and a negative event is annotated for measurements below said threshold. The event status remains unknown for time-points without ground truth data.

To examine model performance across gradations of glycemic dysregulation, we employed glucose thresholds of 160, 180, 200, 220, 240, and 260 mg/dL.

**Sepsis** We follow Moor et al.[20] to annotate sepsis onsets based on the international consensus sepsis-3 definition[30]. However, unlike Moor et al.[20], we only annotate suspected infection on datasets with available blood culture samples and bacterial culture growth information. We do not apply their antibiotic administration heuristic to annotate suspected infections in datasets without explicit blood culture sampling information. In summary, the Sepsis-3 definition requires two components to be observed. The first is a suspected infection typically confirmed by bacterial growth in blood culture samples. The second is a rapid increase in SOFA[31] score. The sepsis onset is annotated where the SOFA score increase has been observed inside a window around the suspected infection.

**Decompensation** Following Harutyunyan et al.[32] we model severe decompensation as the early event prediction of death. A death event is annotated at the end of a deceased patient's intensive care stay. A negative event is annotated at the end of a patient's stay if no mortality information is recorded. Hence, we impute missing information by assuming discharged alive if no information is available to annotate otherwise.

**Hypoxia** Based on Chen et al.[33] we model hypoxia as oxygen saturation below 90% on the emergency department datasets.

**Hypotension** Based on Chen et al.[33] we model hypotension as mean arterial pressure below 65 mmHg on the emergency department datasets.

##### A.5.2 Early Event Prediction Labels

The early event prediction (EEP) label for a given time step is computed as follows: (1) a detection (positive EEP label) is marked if the patient is currently not in a failure state and any time-point in the future within the horizon indicates the patient is in a failure state (positive event). (2) a negative EEP label (a stable patient without any upcoming failure state) is annotated only if there is no failure state annotation (positive event) within the horizon and there is at least one confirmed stable state (negative event) within the horizon. (3) if the patient is currently in a failure state or there is no data confirmed evidence for either the patient being in failure or being stable within the horizon, no EEP label is assigned and no training and evaluation is performed for that specific time step. We use task-specific and clinically relevant prediction horizons from existing literature (8 hours for circulatory failure[17], hyperglycemic events, and sepsis, 24 hours for decompensation[32] and respiratory failure[18], 48 hours for kidney failure[19] and liver failure, and 2 hours for hypotension and hypoxia[33] on the emergency department datasets).

More specifically given event annotations at time $t$ and $e_t \in \{nan, 0, 1\}$ (where $nan$ corresponds to an unknown diagnostic event state) we compute the early event prediction label $y_t$ for a task specific early event prediction horizon $h$ as follows (Eq. (1)):

$$y_t = \begin{cases} 1 & \exists r \in \{t+1, \ldots, t+h\} : e_r = 1 \wedge \neg(e_t = 1) \\ 0 & \exists r \in \{t+1, \ldots, t+h\} : e_r = 0 \wedge \forall r \in \{t, \ldots, t+h\} : e_r \neq 1 \wedge \neg(e_t = 1) \\ nan & \text{otherwise} \end{cases} \quad (1)$$

**Sepsis** Prior work on detection of sepsis[20] used positive labels both before but also after the annotated sepsis onset. We consider the strict early event prediction of the annotated sepsis onset. We consider only the first annotated sepsis onset for any given patient stay and apply the prior function in Eq. (1). Any time point after the first annotated onset is considered to be in an event and following definition Eq. (1) will not be used for further training and evaluation. This yields a positive early event prediction label up to 8 hours before the annotated onset and negative early event prediction labels more than 8 hours before the onset. Any patient without an annotated onset in a source dataset with available body fluid sampling data is considered a negative case and all time steps are annotated using a negative (stable) early event prediction label. Note that this is a prior towards a stable patient in the absence of data, hence the choice to only model the task on datasets with available body fluid sampling data.

##### A.6 K-anonymization

To align with ethical requirements to obtain approval for this study, we perform k-anonymization w.r.t. to age, gender, height, and weight on the following datasets in the harmonized collection as their published version is only considered de-identified: MIMIC-III[5], MIMIC-IV[6], MIMIC-IV ED[14], eICU[7], Zigong EHR[12], PICdb[11], EHRSHOT[3], NWICU[8], and MC-MED[15]. The PSSS and Charité cohorts also have been k-anonymized.

Available age, weight, and height are binned into intervals of size 5. The procedure then ensures there are at least k patients with the same properties in the dataset. Any patient with a value combination (including missingness of a certain attribute) not present at least k times is removed from the dataset. Missing values are imputed with population means.

#### B Model

##### B.1 Architecture

We consider preprocessed irregularly-sampled, multi-variate time-series data, which has been imputed and transformed to a regular hourly time-grid (for details see Supplementary A). Given a dataset of patients we obtain a tensor $\mathbf{X} \in \mathbb{R}^{N \times T \times C}$ where we have $N$ patients, $T$ time-steps, and $C$ channels of information after preprocessing the raw data. Furthermore, let $\mathbf{M} \in \mathbb{B}^{N \times T \times C}$ be a binary presence mask[34] indicating whether a measurement in $\mathbf{X}$ is imputed or ground truth measured data.

The proposed neural network architecture can be split into three distinct components: a time-step encoder, a sequence encoder, and time-to-event prediction heads. First, a time-step encoder module $f : \mathbb{R}^m \to \mathbb{R}^d$, which has shared learned parameters across each time-step to extract time-independent variable interactions. It projects each time-step vector of dimensionality $C$ into a $d$-dimensional vector. Second, a parametrized sequence model $g : \mathbb{R}^{T \times d} \to \mathbb{R}^{T \times d}$ processing the sequence of encoded time-steps and extracting temporal patterns in a causal manner, while preserving the model dimension $d$.

$$\mathbf{H} = g(f(\mathbf{X} \,||\, \mathbf{M})) \in \mathbb{R}^{N \times T \times d} \quad (2)$$

The time-step encoder $f$ is implemented as a two-layer multi-layer perceptron (MLP) with SiLU activations[35]. Following work by Tomasev et al.[34], the time-step encoder is additionally regularized with an L1 penalty on its weights during training. The sequence encoder $g$ is a vanilla Transformer architecture as introduced by Vaswani et al.[36] but we use the more recent SiLU activations. The feedforward hidden dimension is kept at a constant scaling factor 4 from the model dimension (as proposed by Vaswani et al.[36]). We keep the latent dimension of each attention head at a constant 64 and as such the number of attention heads scale linearly with the model dimension. We use learnable Fourier Positional Encodings[37]. The Fourier Positional Encodings were favorable because they allow time to be represented continuously rather than a fixed discrete set of encodings as in traditional sinusoidal positional encodings. Early stages of the project explored training a multi-resolution model, which made the Fourier Positional Encodings an ideal choice. Even though the current model operates at a fixed hourly grid, they could facilitate adaptation to different resolutions more flexibly.

Finally, a collection of $K$ prediction heads for time-to-event (TTE) estimation $\lambda : \mathbb{R}^d \rightarrow [0,1]^{T_{surv}}$ at a prediction horizon of $T_{surv}$ discrete steps. Each hazard prediction head $\lambda_k$ is implemented as a two-layer MLP with SiLU activations and a sigmoid to map to a probability estimate.

$$\forall_{k=1}^{K} \mathbf{Y}_k = \lambda_k(\mathbf{H}) \text{ where } \mathbf{Y} \in \mathbb{R}^{N \times K \times T \times T_{surv}} \tag{3}$$

Typically, if we retrieve embeddings from the model we consider the output $\mathbf{H}$ of $g$ containing a vector representation of every time-step of a given patient.

#### B.2 Objective function

We propose to use the time-to-event estimation[38–40] (i.e., survival modeling) framework as a large-scale pre-training objective for multi-variate time-series data in critical care settings, where it is more relevant (and more clinically actionable) for a downstream application to simply know whether a trajectory will enter certain critical regions rather than having an exact continuous prediction of how the trajectory will progress into these critical regions.

##### B.2.1 Event definition

To perform time-to-event estimation we require an event definition. Intensive care medicine has historically been data-driven and many clinical scoring systems and diagnostic procedures rely on critical decision boundaries on key measurements[31;41] to stratify patients based on data. We use this intuition of threshold-based stratification and propose threshold-based event targets for a selection of core variables. More specifically, we choose a subset of all variables $\mathcal{S}$ of size $|\mathcal{S}| = K$. We define a threshold $\tau_k$ for each variable $k \in \mathcal{S}$. We define an event indicator for patient $i$ at time step $t$ for variable $k$ as $e_{i,t,k} = \mathbb{1}_{[\mathbf{X}_{i,t,k} > \tau_k]}$ an event being a measurement above the defined threshold and respectively $\mathbb{1}_{[\mathbf{X}_{i,t,k} < \tau_k]}$ for an event being a measurement below the defined threshold. We further encode the direction of the comparator with $\delta \in \{0, 1\}$ for $<$ and $\geq$ respectively (Supplementary B.3).

**Target variable set** The model was trained to predict time-to-event distributions for a subset of all harmonized variables. Variables were selected based on two empirical criteria: 1) usage in common clinical scoring functions such as Apache or SOFA scores[27;31;41] and/or 2) observed in most (if not all) of the harmonized datasets. To avoid training signals biased towards a subset of the harmonized sources, we aimed to exclude variables only observed in a subset of the sources unless deemed clinically critical. Most importantly, treatments are excluded from the target set to avoid training signal related to source specific treatment policies.

The final target set of variables $\mathcal{S}$ is: lactate, mean blood pressure, systolic blood pressure, heart rate, troponin-t, partial pressure of oxygen, partial pressure of CO2, fraction of inspired oxygen, pulse oximetry oxygen saturation, respiratory rate, P/F-ratio, creatinine, blood urea nitrogen, urine output rate, weight normalized urine output rate, total bilirubin, direct bilirubin, aspartate aminotransferase, alanine aminotransferase, platelets, white blood cell count, red blood cell count, hematocrit, international normalized ratio (INR), temperature (anywhere on the body), c-reactive protein, pH level in blood, sodium, potassium, calcium, magnesium, chloride, glucose, creatine kinase, creatine kinase-MB.

##### B.2.2 Time-to-event estimation

We perform discrete dynamic survival analysis and discretize time on the same hourly grid as for the input data (Supplementary A), while additionally truncating the survival prediction horizon at $T_{surv}$ steps into the future. Based on prior work[40;42;43], we parametrize and train the hazard function $\lambda_k$ (if applicable conditioned on an event threshold $\tau_k$) using a survival negative log-likelihood over a set of $N$ patients for a total time-series of length $T$ and a truncated survival prediction horizon $T_{surv}$ as follows (we omit the first patient index $i$ and its corresponding outermost summation to simplify notation):

$$\mathcal{L}_{TTE}(k) = \sum_{t=0}^{T}\sum_{r=1}^{T_{surv}} w_{t,r,k}\Big([y_{t,r,k}\log(\lambda_k(r\,|\,\mathbf{H}_t,\tau_k)) + (1 - y_{t,r,k})\log(1 - \lambda_k(r\,|\,\mathbf{H}_t,\tau_k))]\Big) \quad (4)$$

The final loss function is an average over each survival prediction head $\mathcal{L} = \frac{1}{K}\sum_{k=1}^{K}\mathcal{L}_{TTE}(k)$. Let $y_{t,r,k} = \mathbb{1}_{e_{t+r,k}=1 \wedge \forall j<r: e_{t+j,k}\neq 1}$, i.e., we are looking for the first potential occurrence of an event inside the survival horizon $T_{surv}$ starting from the current position $t$. Let a censored position $t$ (for patient $i$ index omitted) be defined as $c_{t,k} = \mathbb{1}_{\nexists \mathbf{M}_{t:t+T_{surv},k}}$, i.e., there is no ground truth measurement observed within time step $t$ and $t+T_{surv}$. The binary weight $w_{t,r,k} = \mathbb{1}_{\neg c_{t,k}\wedge(\exists y_{t,r:T_{surv},k} \vee \forall \neg e_{t:t+T_{surv},k})}$ ensures we only train on cases, which are not censored (we do observe at least one ground truth measurement of variable $k$ as evidence for a (non-)event in the prediction horizon) and either the prediction step is before the observed event ($\exists y_{t,r:T_{surv},k}$) or all event annotations are negative ($\forall \neg e_{t:t+T_{surv},k}$), essentially right censoring at the truncated survival horizon.

At inference the trained hazard prediction heads can be used to obtain a probabilistic estimate on whether the variable with index $k$ will go above (or below) a desired threshold $\tau_k$ at a fixed distance into the future. However, typically at inference we are interested in the cumulative risk up to a certain horizon $h$ into the future. This can be obtained by performing inference on the cumulative failure function $F_k(h\,|\,\mathbf{H_t},\tau_k)$ defined as follows:

$$F_k(h\,|\,\mathbf{H_t},\tau_k) = 1 - \prod_{r=1}^{h}(1 - \lambda_k(r\,|\,\mathbf{H_t},\tau_k)) \quad (5)$$

We illustrate the dual zero-shot inference procedure with a concrete example: predicting circulatory failure risk at a new hospital that was excluded from pretraining, without using circulatory failure labels from any hospital. Circulatory failure is defined by Hyland et al.[17] as mean arterial pressure (MAP) $<$ 65 mmHg and blood lactate (Lact.) $>$ 2 mmol/l. The procedure is as follows:

1. ICareFM processes the patient's hourly time series (vital signs, laboratory values, treatments) and produces a patient state representation $\mathbf{H}_t$ at each hour.
2. Two univariate threshold queries are issued to the pretrained survival heads: "What is the probability that MAP falls below 65 mmHg within 8 hours?" and "What is the probability that lactate rises above 2 mmol/l within 8 hours?", each resolved via the cumulative failure function (Eq. (5)).
3. The two failure probabilities are multiplied under a conditional independence assumption to yield a composite circulatory failure risk score:

$$F_{\text{Circ. Failure}}(8\text{hr}\,|\,\mathbf{H_t}) \approx F_{\text{MAP}}(8\text{hr}\,|\,\mathbf{H_t},\, <65\,\text{mmHg}) * F_{\text{Lact.}}(8\text{hr}\,|\,\mathbf{H_t},\, >2\,\text{mmol/l}) \quad (6)$$

No task-specific classifier or hospital-specific data is involved at any step: task generalization arises from composing physiological threshold queries according to a clinical event definition, where the foundation model handles the temporal extrapolation of future patient state; hospital generalization arises from pretraining representations on a large, heterogeneous multi-institutional dataset. The same procedure applies to any event that can be expressed as, or approximated by, a threshold condition on one or more of the $K$=35 target variables. The model has been trained across the whole value range of each target variable and at inference time the threshold $\tau$ can be arbitrarily varied. Let $Y_k$ be the random variable of values observed for a specific target $k$ in the upcoming horizon and let $\mathbb{P}(Y = y\,|\,\mathbf{H}_i, h)$ be the probability distribution of $Y_k$ taking value $y$ at $h$ time steps into the future. We can thus approximate the expected value of a specific variable $k$ within the upcoming horizon $h$ hours into the future by discretizing the value space into a set of bins $(\tau_l, \tau_u) \in \mathcal{B}$ with a lower and upper threshold boundary:

$$\mathbb{E}[Y_k\,|\,\mathbf{H}_i, h] \approx \sum_{(\tau_l,\tau_u)\in\mathcal{B}} \big(F_k(h\,|\,\mathbf{H_t},\,\tau_l) - F_k(h\,|\,\mathbf{H_t},\,\tau_u)\big) * \frac{(\tau_u+\tau_l)}{2} \quad (7)$$

This can be used to approximate expected value distributions within an upcoming horizon $h$. We can then approximate clinical scores such as Apache[41] or MELD[27] several hours into the future, giving yet another angle to perform zero-shot predictions without any form of task adaptation on the model.

For example, to estimate MELD scores[27] we compute as follows:

$$\begin{aligned}\mathbb{E}[MELD|\,\mathbf{H}_i, h] \approx\,& 6.43 \\ &+ 3.78 \cdot \ln(\mathbb{E}[Bilirubin|\,\mathbf{H}_i, h]) \\ &+ 11.2 \cdot \ln(\mathbb{E}[INR|\,\mathbf{H}_i, h]) \\ &+ 9.57 \cdot \ln(\mathbb{E}[Creatinine|\,\mathbf{H}_i, h])\end{aligned} \tag{8}$$

To compute zero-shot risk estimates for respiratory failure we use the predicted cumulative failure probability on a specific P/F ratio. To predict kidney failure risks we use the union of the cumulative probability to reach either a high creatinine threshold or a low urine output. For sepsis we approximate the systemic inflammatory response syndrome (SIRS) criteria[44]. For the onset of mortality we approximate the Apache Score[41]. For early event prediction of hyperglycemia we simply provide the cumulative failure probability to reach a critical glucose threshold.

#### B.3 Survival prediction head conditioning

To enable flexible zero-shot inference with arbitrary event thresholds, we condition each hazard prediction head $\lambda_k$ on both the threshold magnitude and direction. For each target variable $k \in S$, we define:

**Variable-threshold embedding** Each variable $k$ has a learnable embedding $\mathbf{u}_k \in \mathbb{R}^{d/2}$, which is scaled by the sampled threshold[45;46]:

$$\mathbf{u}_k^{\tau} = \tau \cdot \mathbf{u}_k \tag{9}$$

**Threshold direction embedding** We sample a direction indicator $\delta \in \{0, 1\}$ that specifies the comparator for event definition:

$$e_{i,t,k} = \begin{cases} \mathbb{1}_{X_{i,t,k} < \tau} & \text{if } \delta = 0 \\ \mathbb{1}_{X_{i,t,k} \geq \tau} & \text{if } \delta = 1 \end{cases} \tag{10}$$

This direction indicator is embedded as $\mathbf{d}_\delta \in \mathbb{R}^{d/2}$, enabling the model to distinguish between "below threshold" ($\delta = 0$) and "above threshold" ($\delta = 1$) events. Explicit direction modeling is necessary because the time-to-event distributions for crossing a threshold from below versus from above are not complementary: if $F_k(h \mid H_t, \tau, \delta = 1) = p$, we cannot assume $F_k(h \mid H_t, \tau, \delta = 0) = 1 - p$, as each direction exhibits distinct hazard patterns influenced differently by patient state and temporal dynamics.

**Conditioning mechanism** The conditioned representation is formed by concatenating the scaled variable embedding with the direction embedding:

$$\tilde{\mathbf{u}}_k^{\tau,\delta} = [\mathbf{u}_k^{\tau} \,||\, \mathbf{d}_\delta] \in \mathbb{R}^d \tag{11}$$

The hazard prediction head then operates on the concatenation of the patient state representation $H_t \in \mathbb{R}^d$ and the conditioned embedding:

$$\lambda_k(r \mid H_t, \tau, \delta) = \mathrm{MLP}_k([H_t \,||\, \tilde{\mathbf{u}}_k^{\tau,\delta}]) \tag{12}$$

where $\mathrm{MLP}_k : \mathbb{R}^{2d} \to [0, 1]^{T_{\text{surv}}}$ is a variable-specific two-layer network with SiLU activations and sigmoid output. This design allows the model to learn direction-aware continuous threshold responses during training while maintaining the ability to query arbitrary thresholds and directions at inference time.

##### B.3.1 Bivariate events

The proposed event definition treats all events from separate variables independently. However, in practice, many of the observed variables are correlated. We extend the training framework by also considering bivariate events. Given two variables $k_1, k_2 \in \mathcal{S}$ we define a joint event as $e_{i,t,(k_1,k_2)} = e_{i,t,k_1} \wedge e_{i,t,k_2}$. Given the joint event $e_{i,t,(k_1,k_2)}$ annotation, the same objective as before can be optimized. To maintain scalability of the approach in practice we sample a fixed number of variable combinations during training at random in each training step, add a single additional hazard prediction head $\lambda_{Bivariate}$ to the network and condition the prediction head on the choice of the two variables by providing an appropriate embedding vector representing each of the $k_1$ and $k_2$ as an input to $\lambda_{Bivariate}$.

To condition the bivariate hazard prediction head on the variable pair, we re-use the threshold-conditioned embeddings from the univariate case. For a bivariate event defined over variables $k_1, k_2 \in \mathcal{S}$, each with independently sampled thresholds $\tau_1, \tau_2$ and directions $\delta_1, \delta_2$, the conditioning is formed by concatenating the patient state with both conditioned variable embeddings:

$$\lambda_{\text{Bivariate}}(r \mid H_t, \tau_1, \delta_1, \tau_2, \delta_2) = \text{MLP}_{\text{Bivariate}}([H_t \,||\, \tilde{\mathbf{u}}_{k_1}^{\tau_1,\delta_1} \,||\, \tilde{\mathbf{u}}_{k_2}^{\tau_2,\delta_2}]) \tag{13}$$

where $\text{MLP}_{\text{Bivariate}} : \mathbb{R}^{3d} \to [0,1]^{T_{\text{surv}}}$ is a single shared prediction head for all bivariate combinations, enabling the model to learn joint event patterns across arbitrary variable pairs.

#### B.4 Context to related foundation model approaches

Several families of foundation models are relevant to positioning ICareFM. We review each below and note the gaps that motivated our approach.

##### B.4.1 Code-first foundation models

A growing number of foundation models operate on structured electronic health record (EHR) codes. Delphi[47] is a generative transformer trained on millions of coded health records. MOTOR[38] uses a time-to-event objective but takes predominantly coded EHR data as input (see Supplementary B.6 for an empirical comparison). CLIMBR[48] learns EHR representations from coded sequences using a language modeling objective and was evaluated on the EHRSHOT benchmark[3]. ETHOS[49] demonstrates zero-shot health trajectory prediction via autoregressive token generation; while it incorporates discretized lab values and vitals alongside diagnostic and procedure codes, it was trained on a single dataset (MIMIC-IV) with continuous measurements coarsened into quantile bins, limiting its scale and resolution relative to a multi-site foundation model. Its evaluation focuses on per-admission outcomes (mortality, readmission) and first-day SOFA score estimation at ICU admission, rather than continuous, online prediction of organ-failure events at each time step over variable future horizons. Guo et al.[50;51] have studied the adaptability and robustness of coded EHR foundation models across institutions, reporting substantial performance variation under distribution shift.

While these models demonstrate label efficiency, they rely on retrospective abstractions that are often unavailable at the bedside in real time, such as administrative and diagnostic codes. These codes are typically assigned after discharge, provide coarse temporal summaries of patient state, and are constrained by coding schemas that vary across institutions and countries. As a result, code-first models do not capture the continuous physiological dynamics (e.g., heart rate trajectories, blood pressure trends, or evolving laboratory values) on which real-time clinical decisions in critical care depend. The empirical evaluation in Supplementary B.6 further illustrates this modality gap: despite encoding health-relevant temporal patterns, MOTOR's transfer to high-frequency ICU time series was substantially affected by the shift from coded to continuous data.

##### B.4.2 General-purpose time series foundation models

General-purpose time series foundation models have achieved strong performance on standard forecasting benchmarks. Chronos[52] and Chronos-2[53] are among the most established, with Chronos-2 supporting multivariate input (see Supplementary B.6 for an empirical evaluation on ICU data). Other recent models include Toto[54], Moment[55], TimesFM[56], and Moirai[57]. MIRA[58] bridges toward the medical domain but is still a univariate model.

These models optimize forecasting objectives rather than clinical event risk, and remain largely unevaluated in treatment-confounded critical care settings where therapeutic interventions actively alter physiological trajectories. Furthermore, most of these models are univariate and do not support multivariate input. Their pretraining corpora and evaluation benchmarks contain minimal health-related data and do not reflect the multivariate, sparse, and irregularly sampled nature of ICU recordings. As shown in Supplementary B.6, even after substantial domain adaptation (100k fine-tuning steps), Chronos-2 achieved poor clinical event discrimination on ICU prediction tasks.

###### B.4.3 Time-to-event foundation models

In this work we propose core modifications to existing approaches for time-to-event pretraining (such as done in MOTOR by Steinberg et al.[38]), which are essential for obtaining strong performance on intensive care data streams (heterogeneous multi-variate numerical time-series). We highlight these modifications and how they differ from existing approaches in the following paragraphs.

**Continuous Event Targets** While MOTOR by Steinberg et al.[38] does encode numerical data by binning and assigning a discrete code (token) to each bin at its input, the target and output space of MOTOR[38] (or also e.g. STRAFE[59]) only considers the presence of a certain event and omits a potentially associated value. This is sensible for coded electronic health records (paired with billing codes for diagnosis, prescriptions, etc.). However, in real time intensive care data streams, these data (diagnostic codes, procedure codes, etc.) have not been created yet, while the patient is being monitored in the ICU in real time (or other critical care settings such as the emergency department). ICareFM is trained with survival prediction heads across channels of physiological measurements learning a continuous response function from thresholds in the data space to survival curves conditioned on the patient state. This goes beyond the simple presence of a code, incorporating the observed values into the survival objective and preserving the continuous structure of the data space.

**Random Sampling of Event Thresholds during Training** ICareFM is trained by sampling thresholds to define events across the value space at random during training. This allows flexibly choosing arbitrary thresholds at inference time, while encouraging a stable mapping from thresholds to survival curves without overfitting to specific fixed event thresholds.

**Conditioning on Event Thresholds** MOTOR[38] trains separate survival heads for each specific event in the target code set. In ICareFM, we train a separate prediction head for each variable (or a single head for a bivariate combination thereof (Supplementary B.3.1)) but then condition the prediction head on the target threshold for the event. This models a continuous response, shares parameters across the value space, and allows for flexible threshold choice at inference time.

**Summary** Continuous event encoding lets the model generalize across thresholds unseen during training. Random sampling of event thresholds acts as a regularizer, preventing overfitting to fixed cutoffs. Conditioning prediction heads on the threshold shares parameters across the value space, reducing the number of required heads from $O(|\text{thresholds}|)$ to one per variable.

###### B.4.4 Contrastive learning approaches

Contrastive learning has shown promise for representation learning from clinical time series and biosignals, including neighborhood contrastive learning for online patient monitoring[60], contrastive learning of cardiac signals[61], multi-modal contrastive learning for clinical time series[62], and contrastive approaches for unsupervised domain adaptation of time series[63]. We explored contrastive pretraining during the development of ICareFM but did not pursue it further due to two fundamental challenges in the multi-site critical care setting.

First, effective contrastive learning requires augmentation strategies that can a priori disentangle domain shifts, such as hospital-specific artifacts in the form of differing measurement frequencies, sensor calibration, and recording practices, from genuine physiological signal variation. In a multi-continental setting with substantial cross-hospital heterogeneity, designing such augmentations is extremely difficult. A contrastive objective risks either collapsing clinically meaningful variation or remaining sensitive to spurious recording-specific patterns. Second, contrastive pretraining yields an embedding model without explicit zero-shot event prediction capabilities. The resulting representations require downstream task-specific adaptation (e.g., linear probing or fine-tuning) for each clinical application, making the approach less suited for a directly deployable foundation model where the goal is to provide clinical decision support at inference time without retraining. The time-to-event pretraining objective sidesteps these challenges by directly learning clinically interpretable survival distributions conditioned on user-specified event thresholds.

##### B.5 Model size - Ablation study

To find an optimal configuration in terms of learnable parameters we scale the model dimension $d$ from 128 to 1024 and model depth from 1 to 4 transformer blocks. The survival prediction horizon $T_{surv}$ is fixed at 24 hours and we perform dynamic event thresholding where an event threshold is sampled at random during training time from the lower and upper 40% of a standard normal distribution. We train on the complete dataset and report performances in-distribution of the pretraining data.

We report joint test set TTE loss performance across all variables and datasets for each model. We then perform linear probing for 6 downstream tasks: decompensation at 24h, circulatory failure at 8h, severe respiratory failure at 24h, severe kidney failure at 48h, severe liver failure at 48h, and hyperglycemia at 8h. We report task-averaged AuROC and AuPRC for each model across all datasets, where for both training and evaluation all predictions and labels are simply aggregated into a single collection irrespective of their source hospital.

**Ablation conclusion** Based on the results in Figure S5 A) we consider the validation loss for the pretraining task and notice overfitting w.r.t. the pretraining objective after approx. 30 million parameters. We take a model from this parameter region (20-30 million parameters) and consider the best performing model on the downstream objectives in this region, which yields an architecture with depth 3 and 512 hidden dimensions. While results on the downstream tasks suggest we could choose a much larger model, we cautiously select a model that does not overfit to the pretraining for two reasons: 1) the goal is to test the model's out-of-distribution performance and as such overfitting to the training distribution is not advisable 2) the goal is to directly leverage the model's survival prediction heads for zero-shot early event predictions without any linear probe training.

#### B.6 Pretraining objective - Ablation study

We perform an ablation w.r.t. the type and configuration of our objective function. We compare 5 different pretraining objectives by training the same backbone architecture from scratch on our large harmonized dataset:

- **Forecasting** here the survival prediction heads $\lambda_k$ are replaced with a Transformer decoder[64] regressing the exact values of all ground truth measurements (we mask out imputed data from the loss) within the prediction horizon and optimizing Huber Loss[65] (to stabilize training for the noisy data).

- **Indicator** This is a time-to-event objective with a simplified event definition, similar to the MOTOR target event definition by Steinberg et al.[38] for coded electronic health records. The event definition for patient $i$ at time $t$ for variable $k$ reduces to $e_{i,t,k} = \mathbb{1}_{[\mathbf{M}_{i,t,k}]}$, i.e., whether a ground truth measurement has been obtained. In the context of intensive care measurements, this can only be applied for irregularly-sampled variables such as lab tests (and potentially treatments) as they are an indication of a clinician's state belief of the patient. We exclude regularly measured vitals in this objective as their regular acquisition is trivially predicted.

- **Fixed Tau** This is a time-to-event objective with an event definition as introduced in Supplementary B.2.1. For each of the $K$ target variables a fixed $\tau_k$ is chosen. Given centered data the chosen threshold is symmetrically applied below (as $-\tau_k$) and above the mean of the distribution.

- **Mixed** This is an intermediate step between the *Indicator* and the *Fixed* case, where for all irregularly measured variables the *Indicator* objective is applied and for all regularly measured variables the *Fixed* objective is applied.

- **Dynamic Tau** This is the proposed time-to-event objective with an event definition as introduced in Supplementary B.2.1. For each variable and time-step an event threshold $\tau_k$ is sampled from a uniform distribution up to a large extreme value e.g. 4 standard units of the scaled data distribution. Sampling thresholds at random during training allows fixing a specific threshold of interest at inference for which a probability to cross that threshold should be estimated by the model. In this setting the hazard prediction heads $\lambda_k$ are conditioned on the threshold (see Supplementary B.3) to learn a continuous response function from threshold to survival.

We further evaluate ICareFM against two state-of-the-art foundation models pretrained on out-of-domain data to assess the importance of domain-specific pretraining versus general-purpose or adjacent-domain approaches:

- **Chronos-2** by Ansari et al.[53] represents one of the strongest general-purpose time series foundation models currently available. It handles sparsity through its encoder architecture and supports multivariate time series data. However, its multivariate capabilities were primarily developed using synthetic data. While demonstrating excellent performance across large benchmark collections, its training corpus and evaluation benchmarks contain minimal health-related data, creating a substantial domain gap for clinical applications.

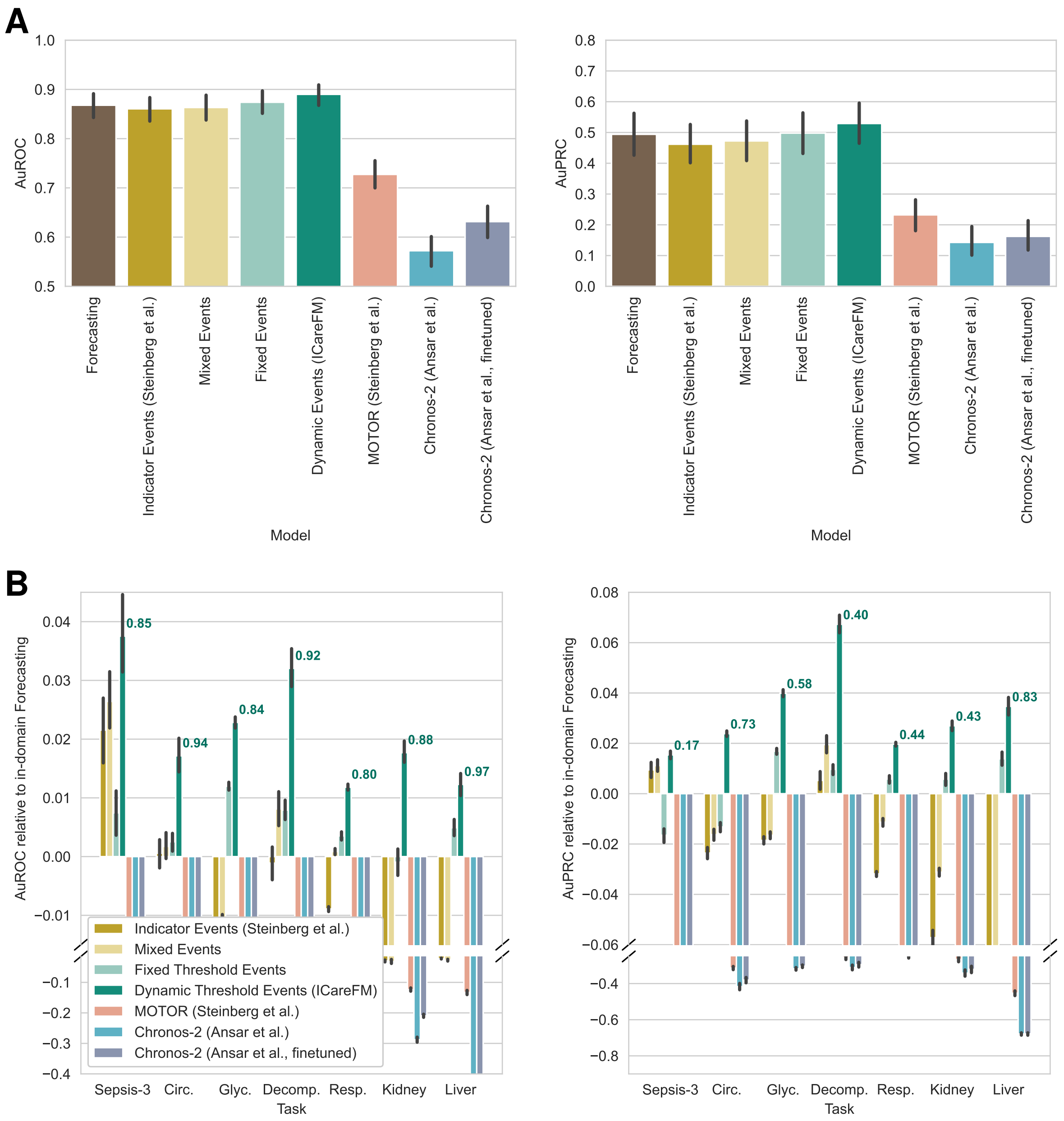


**Figure S2: Comparison of foundation model approaches for ICU time series prediction. A)** Absolute performance comparison showing raw AuROC (left) and AuPRC (right) distributions across all task-dataset combinations. Bar plots show mean performance with 95% confidence intervals. ICareFM Dynamic Threshold Events achieve mean AuROC of 0.89 and mean AuPRC of 0.53, substantially exceeding alternative pretraining objectives with the same architecture trained from scratch (Forecasting, Indicator Events as proposed by Steinberg et al.[38] for coded EHR data, and further event definitions considering numeric values of observed measurements in the event definitions). General-purpose time series models (Chronos-2[53]) exhibit poor discrimination (median AuROC $< 0.65$) for clinical event prediction, even after fine-tuning the network with the original pretraining objective for $100{,}000$ steps on the ICareFM training set. MOTOR pulls ahead of Chronos-2, indicating that training on health data is beneficial, but the data modality shift from predominantly coded EHR data collected for billing purposes to critical care time series data dampens its utility for relevant applications in intensive care. **B)** Relative performance compared to in-domain forecasting baseline across six clinical prediction tasks. Bars show mean difference in AuROC (left) and AuPRC (right) between each foundation model approach and the in-domain forecasting baseline, aggregated across all available datasets per task. Positive values (above zero line) indicate superior performance to in-domain forecasting. Numbers above ICareFM Dynamic Threshold bars indicate absolute in-domain forecasting performance (AuROC/AuPRC) for reference. Error bars represent 95% confidence intervals across datasets. Y-axis breaks accommodate large negative performance gaps for general-purpose time series models and models trained on coded EHR data (Chronos-2[53], MOTOR[38]). ICareFM's dynamic threshold survival prediction (dark teal) consistently outperforms, while general-purpose forecasting models show substantial performance degradation.

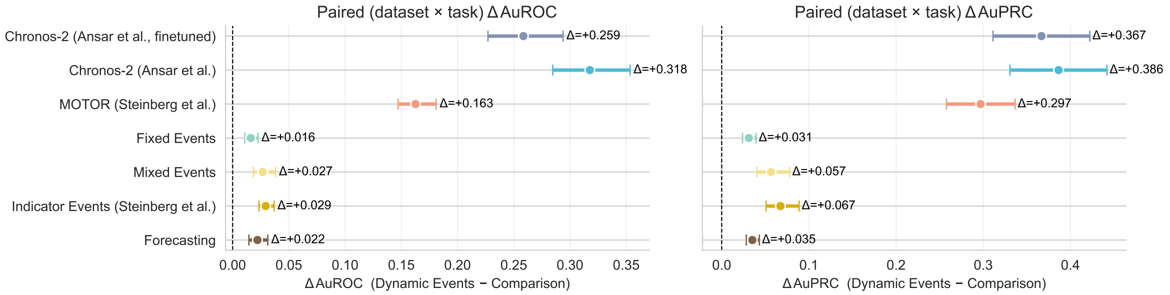


**Figure S3: Model comparison to ICareFM on pairwise (dataset and task) differences A)** Each point shows the mean difference in test-set AuROC (left) and AuPRC (right) computed on paired dataset $\times$ task combinations across 8 ICU databases (MIMIC-IV, eICU, NWICU, HiRID, UMCdb, SICdb, PICdb, Zigong EHR) and 7 clinical prediction tasks. Horizontal bars denote 95% confidence intervals from bootstrapping. Positive values indicate that Dynamic Tau Events outperforms the comparison model; intervals not crossing zero indicate a statistically significant difference at the 5% level (significance persists when applying Bonferroni correction for testing multiple models to p-values computed from the bootstraps).

  – **Source Model** We perform embedding inference for each time step on the Chronos-2 model as made available on the HuggingFace platform[1].
  – **Domain Adapted** We adapt the Chronos-2 model by fine-tuning on our intensive care datasets with the Chronos-2 quantile regression objective. We obtain a checkpoint after $10k$, $50k$, and $100k$ fine-tuning steps. For each of these checkpoints we perform inference, obtain a time-step representation vector by mean pooling across channels, and train logistic regression probes. $100k$ steps marginally comes out with best results (selected per validation data).

- **MOTOR**[38] We obtained pretrained model weights from the HuggingFace platform[2], originally trained on a large coded EHR dataset from Stanford Medicine. Applying MOTOR to our data required mapping harmonized ICareFM concepts to medical codes compatible with MOTOR's tokenizer. That process proved highly non-trivial. MOTOR's tokenizer spans multiple code vocabularies (SNOMED, LOINC, etc.), each offering numerous candidates for a single harmonized concept and sometimes also no good candidate match for one of our concepts could be found due to MOTOR's limited vocabulary size. For instance, a fundamental measurement like mean arterial blood pressure maps to multiple SNOMED and LOINC codes. This mapping ambiguity exposes a core challenge in coded EHR systems: despite their promise of standardization, variation in healthcare system practices causes substantial heterogeneity in coding system usage. Following a best-effort mapping, we transformed our data and performed embedding inference using the pretrained model.

We trained on nine intensive care datasets (MIMIC-IV, eICU, NWICU, UMCdb, HiRID, SICdb, PICdb, Zigong EHR, and INSPIRE) and evaluated in-distribution performance via linear probe comparisons across pretraining objectives. For *Mixed* and *Fixed Tau* configurations, we set $\tau_k$ to the 5th- or 10th-percentile of a standard normal distribution, selecting final thresholds based on validation performance. In the *Dynamic Tau* setting, thresholds were sampled uniformly across the scaled data space (emphasizing distributional tails given prior standardization). Results appear in Figure S2: panel A presents aggregate performance by model/objective, while panel B decomposes task-specific performance relative to the forecasting baseline.

**Ablation conclusion** Supplementary B.6 shows paired differences across datasets and tasks for different models. The forecasting baseline and all time-to-event variants (Indicator, Mixed, Fixed, and Dynamic Events) were pretrained from scratch using identical backbone architectures, isolating the contribution of the pretraining objective to downstream early event prediction. Performance improves consistently as event definitions progress from simple indicator events (as proposed by Steinberg et al. for coded data) to progressively more fine-grained representations of numerical time series structure. This advantage likely stems from two factors: (1) improved alignment between pretraining objective and downstream applications, and (2) time-to-event formulations employ carefully bounded binary cross-entropy loss, which exhibits greater resilience to outliers and measurement noise common in ICU data.

[1] `https://huggingface.co/amazon/chronos-2`
[2] `https://huggingface.co/StanfordShahLab/motor-t-base`

Despite severe domain shift from its pretraining corpus, Chronos-2 achieved above-random performance on clinical prediction tasks. However, substantial computational investment in fine-tuning (up to 100k steps) yielded only marginal improvements. These results suggest an important direction for future work: incorporating multivariate, sparse, irregularly-sampled health time series directly into general-purpose foundation model pretraining to enhance clinical utility without requiring extensive domain adaptation.

Embeddings from MOTOR substantially outperformed Chronos-2, confirming that MOTOR successfully encodes health-related temporal patterns despite its coded EHR training data. Nevertheless, the modality shift from low-frequency coded records to high-frequency critical care time series impeded direct transfer performance. Given the non-trivial mapping between harmonized concepts and MOTOR's tokenizer, we consider the *Indicator Event* baseline more informative: training our architecture from scratch on harmonized ICU data using MOTOR's objective produced strong performance, which we further improved by incorporating fine-grained information from the high-frequency numerical structure inherent to critical care monitoring. Preliminary results from concurrent work exploring self-supervised pretraining for critical care time series on single-institution datasets[66–68] further support the value of domain-specific pretraining for ICU data. These findings also ruled out the relevance of any other code-first foundation model such as Delphi[47] to meaningfully transfer to the intensive care setting (see Supplementary B.4 for a broader discussion).

#### B.7 Survival horizon - Ablation study

Conceptually, a survival model should be trained up to the largest observed time in the cohort. In practice, many algorithms need to strike a balance between theoretical accuracy and technical feasibility, in this case often bounded by available memory resources on e.g. GPU hardware. To accommodate efficient training the maximum survival horizon considered in training can be truncated. Yèche et al.[40] report that truncation can even be beneficial due to low signal-to-noise ratio at large prediction horizons and truncation can act as a form of regularization. We investigate the impact of the truncated survival horizon $T_{surv}$ during pretraining for different downstream tasks.

The results in Figure S5 B/C) indicate that adapting the survival horizon yields some performance improvements w.r.t. linear probe and zero-shot performances on different downstream tasks, but for some tasks performance can also slightly decrease again for very large horizons. Events at larger distances suffer from increased confounding actions[69], which are common in intensive care, and as such the additional events considered during pretraining at the larger distances provide limited actual predictable and learnable signal, under increased noise.

**Ablation conclusion** Due to varying performance implications depending on the downstream task, observed in the performed ablation, we fix $T_{surv} = 48$ for practical reasons to facilitate zero-shot inference and trajectory visualizations on any relevant downstream application up to the full relevant prediction horizon. We note that due to strong trajectory changes in intensive care patients and the ultimate goal to discharge the patient from the station, performing predictions far beyond 48 hours (2 days) does not yield any practical advantage in an intensive care setting.

#### B.8 Bivariate events - Ablation study

The core pretraining objective relies on events defined on a single variable (univariate) as introduced in Supplementary B.2.1. However, the objective function itself (Supplementary B.2.2) is agnostic to the way an event has been derived from the data. Modeling only univariate events does not encourage the model to learn correlations across the covariates and assumes independent events. Ultimately, one could model events defined on the full set of variables ($k$-variate events). However, this would lead to extremely sparse events and potentially slow training. We investigate adding bivariate events to complement the univariate events in the pretraining.

In Figure S4 we compare zero-shot (OOD) performance and probe (domain and task adaptation) performance, when pretraining the foundation model with and without bivariate events. We find a statistically significant improvement in zero-shot performance, but no noticeable performance implications when target data is used for domain and task adaptation with a probe prediction head.

**Ablation conclusion** Due to the observed zero-shot performance improvements we train the foundation model including bivariate events in the pretraining. Future work should address the scalability issues to consider

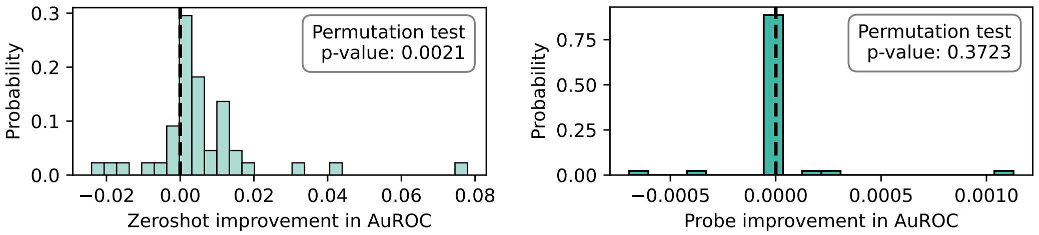


**Figure S4:** Comparing out-of-distribution (held-out hospital) zero-shot and linear probe (domain and task adaptation) performance when training with and without bivariate events (Supplementary B.3.1) in the pretraining objective. We observe a statistically significant (permutation test on the paired difference in AuROC for each dataset and task pair) improvement for the zeroshot transfer performance. The additional bivariate consistency in pretraining however does not noticeably benefit downstream task adaptation probe performance.

events defined on even larger joint variable sets to further improve the model's consistency and better distill the correlations across the variables.

#### B.9 Training

##### B.9.1 Implementation

Training is implemented in PyTorch[70] and loss functions are optimized using AdamW[71]. Alongside the chosen self-supervised objective for time-series modeling, we also add an $L_1$ regularization term on the time-step encoder[72]. We do not apply any form of explicit reweighting or regularization on the objective to deal with any type of imbalances in the data during pretraining or downstream training. We do not apply any type of post-hoc recalibration.

For pretraining we consider all the intensive care datasets in the harmonized collection (see Tables S1 and S2), that is MIMIC-III, MIMIC-IV, eICU, NWICU, HiRID, UMCdb, SICdb, Inspire, PICdb, Zigong EHR, and PSSS. For generalization experiments the target is typically excluded. Additionally, due to data leakage concerns (Supplementary A.2.2), when transferring to MIMIC-IV we exclude MIMIC-III, and when transferring to HiRID we exclude PSSS, unless explicitly stated otherwise for a specific experiment.

The adaptation settings described in Section 3.3 and Section 2.2 are implemented as multi-stage training. First ICareFM is pretrained using the self-supervised objective until early stopped on the validation loss. Then each adaptation setting proceeds as follows:

- **(ii)** *External adaptation (Domain generalization)* External labels are used to finetune the pretrained network. We investigate different fine-tuning approaches, fine-tuning only a randomly initialized linear prediction head, fine-tuning the entire network with a randomly initialized linear prediction head, fine-tuning only an appropriate survival prediction head (e.g. the mean arterial pressure head for circulatory failure), and fine-tuning the entire network and considering the output of an appropriate survival prediction head. The best approach is selected on the validation set of the external data.

- **(iii)** *Local adaptation* We initialize from the pretrained (only self-supervised pretraining) ICareFM network and again compare either a randomly initialized linear head or a survival head from the pretrained network as final output prediction head and compare training only that head or the entire network, we also considered LoRA[73] to adapt the backbone time-series encoder. The best configuration is selected on the validation set of the target dataset. Note that in the scaling study the validation set is part of the sampled training budget and hence grows in size as we scale the available data.

- **(iv)** *Staged adaptation* We initialize from the best network determined in the *External Adaptation* stage and fine-tune either only the prediction head or the entire network and then select the best configuration based on the validation set. Note that also here for the sample size scaling studies the validation set is part of the sampled training budget and hence grows in size as we scale the available data.

###### B.9.2 Infrastructure

The ICareFM foundation model was trained on the Alps Research Infrastructure[74] at the CSCS (Swiss National Supercomputing Centre). A training run of ICareFM uses a single node of Nvidia's Quad GH200, which tightly couples four Grace Hopper Superchip (GH200) GPUs (and Arm processors) on a single compute node[75]. Each GPU has 96GB of HBM3 memory for a total of 384 GB of GPU memory paired with a total of 480GB LPDDR5 ECC memory attached to the Arm processors. Each Arm Neoverse V2 CPU has 72 processing cores for a total of 288 cores on a single compute node.

Computational data analysis was performed at Leonhard Med (`https://sis.id.ethz.ch/services/sensitiveresearchdata/`) secure trusted research environment at ETH Zurich on a heterogeneous collection of CPU and GPU resources.

Validation experiments involving the Charité dataset were executed exclusively on the internal high-performance computing (HPC) infrastructure of Charité – Universitätsmedizin Berlin. At no point were clinical data or trained models including patient data from the Charité transferred to or accessed by external partners.

Validation experiments involving the RBKICU dataset were executed exclusively on the internal infrastructure of the Robert Bosch Krankenhaus Stuttgart. At no point were clinical data or trained models including patient data from the Robert Bosch Krankenhaus transferred to or accessed by external partners.

### C Evaluation

#### C.1 Baseline model selection and tuning

We tuned strong baseline models separately for each downstream task, dataset, and sample size, as described below.

##### C.1.1 Gradient boosted trees

We employ LightGBM[77] (gradient-boosted decision tree) as our primary classical machine learning baseline. The model utilizes an extended feature engineering pipeline that builds upon the approach of[22], incorporating statistical features computed over multiple temporal horizons (detailed in Supplementary Section A.4.4). This choice is motivated by extensive prior benchmarking from Burger et al.[78] and Yèche et al.[23] demonstrating that gradient boosted trees consistently achieve dominant task-specific performance in the intensive care time series domain. For each combination of downstream task, target dataset, and training set size, we perform hyperparameter optimization to identify the optimal LightGBM configuration. For each downstream task, target dataset, and set of fixed hyperparameters we perform three distinct training runs with different random seeds (which also affects the subsample used for training).

##### C.1.2 Deep sequence models

We employ Gated Recurrent Unit (GRU) networks as our deep learning baseline architecture. This selection is based on benchmarking by Burger et al.[78] comparing multiple modern sequence architectures including Transformers, GRU, Mamba (state-space models), and xLSTM (a modern alternative to the established long-short-term-memory recurrent neural network architecture), where GRU emerged as the strongest overall performer for intensive care time series modeling. Our GRU implementation incorporates established architectural enhancements from prior work[72]: we provide both imputed data and ground truth presence masks as dual inputs to the network, and apply L1 regularization to the time-step embedding layers to prevent overfitting. As with gradient boosted trees, we perform hyperparameter optimization for each combination of downstream task, target dataset, and training set size. For each downstream task, target dataset, and set of fixed hyperparameters we perform three distinct training runs with different random seeds (which also affects the subsample used for training).

##### C.1.3 Best baseline selection

For each specific combination of downstream task, target dataset, and training set size, we select the best-performing model between the optimized LightGBM and optimized GRU architectures based on validation set performance. This selection process creates state-of-the-art scaling curves that represent a strong task-specific performance for each dataset, downstream task, and sample size combination using locally available data from the target hospital in a supervised machine learning setting.

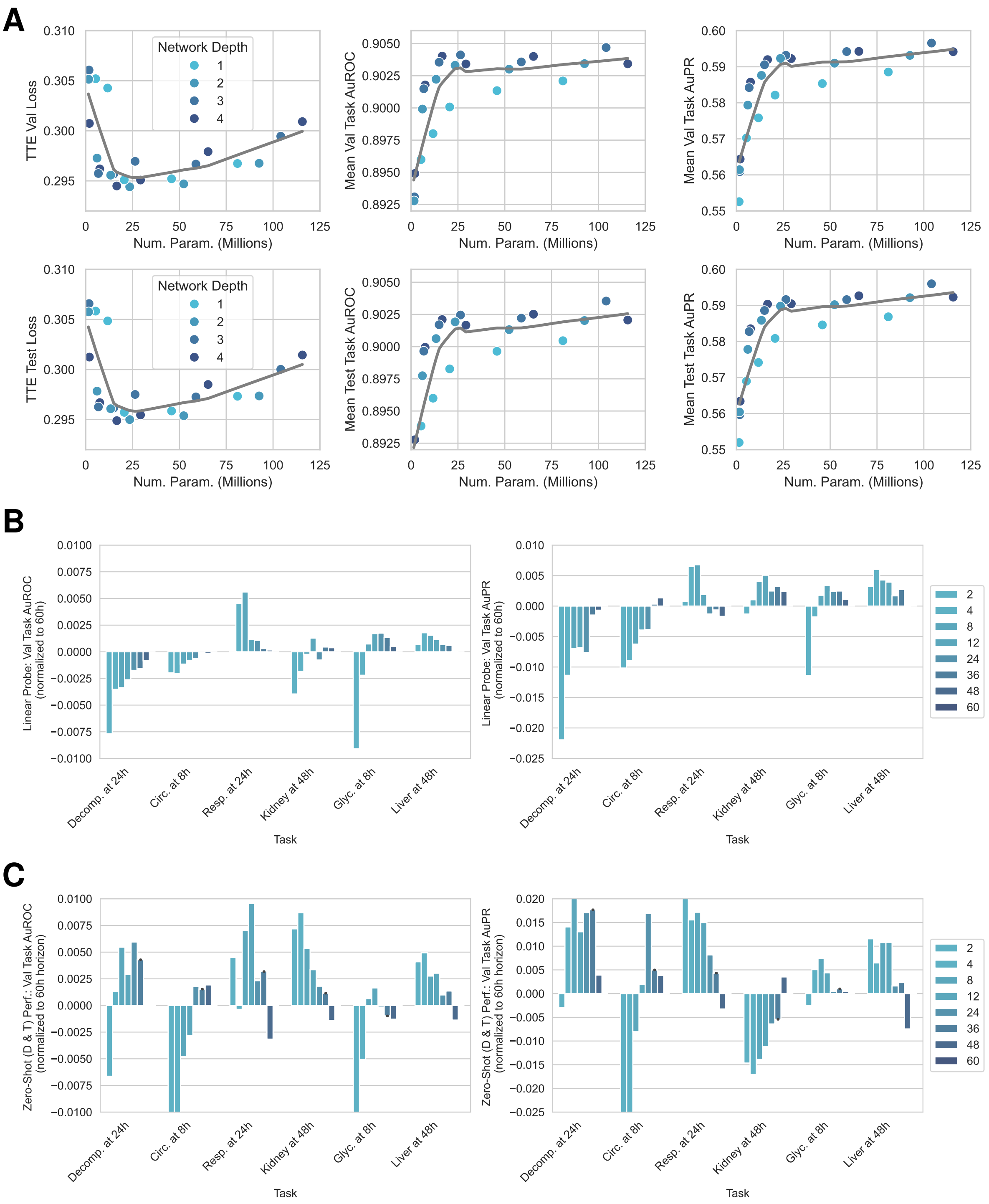


**Figure S5: Model Ablations A)** Model size scaling of the Transformer architecture by hidden dimension and Transformer depth (colored by depth). Left column shows the time-to-event pretraining objective loss, middle column average AuROC, and right column average AuPRC across 6 downstream task linear probes. Grey is showing a Loess[76] regression line. **B/C)** Different truncated survival horizons $T_{surv}$ introduced in Supplementary B.7. Row **B** shows linear probe performance and Row **C** shows zero-shot performance (no task adaptation).

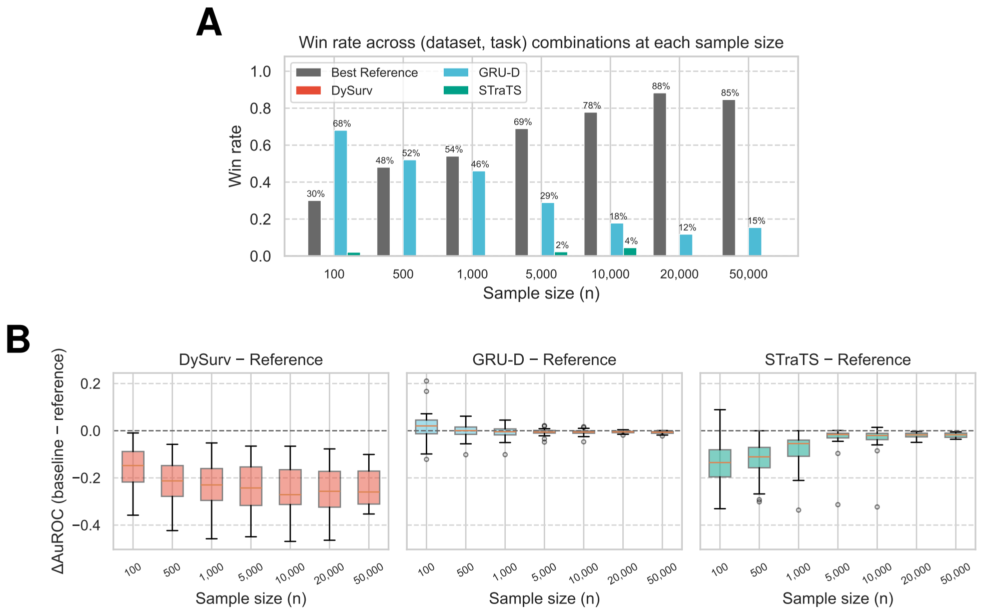


**Figure S6: Extended neural network reference performance comparison. A)** Win rate of each alternative architecture (DySurv, GRU-D, STraTS) against the best reference baseline (per-configuration optimum of tuned LightGBM and GRU) across all dataset–task combinations at each training set size. Percentages indicate the proportion of configurations in which each model achieves the highest AuROC. The best reference baseline dominates increasingly with growing sample size, exceeding 75% win rate beyond 10,000 patients. **B)** Distribution of AuROC differences (alternative architecture minus best reference baseline) across dataset–task combinations at each sample size. The dashed horizontal line at zero indicates parity with the reference. DySurv consistently underperforms with median deficits of approximately 0.20 AuROC. GRU-D performs near parity with slight advantages at small sample sizes. STraTS shows moderate deficits that diminish with increasing data availability.

#### C.2 Extended comparison

Our LightGBM baseline covers a selection of strong prior works across early event prediction for circulatory [17], respiratory [18], and kidney failure [66] all relying on well-tuned implementations of LightGBM. Based on prior benchmarking work [23;78] we choose GRU enriched with strong domain-specific adjustments [72] successfully applied in kidney failure prediction as the deep learning representative architecture.

There is a large body of work in the deep learning literature, which proposes to address the challenges of irregular and sparse measurements encountered in critical care data to further improve performance. However, none of those have typically performed such a large-scale benchmarking effort across many downstream tasks and datasets. We compare against three further task-specific modeling architectures, each representing a distinctively different approach to address the challenge of early event prediction over critical care data using deep neural networks.

**DySurv** as proposed by Mesinovic et al. [79] is an LSTM based sequence model. The distinctive feature of this model is that its training objective is a time-to-event loss, which can then be set to a fixed horizon at inference time for a specific cumulative failure early event prediction tasks at fixed horizons. Additionally DySurv proposes a joint optimization with a VAE encoder bottleneck. Overall, we found this approach to drastically underperform. The joint optimization problem with time-to-event optimization and a VAE bottleneck seemed unfeasible to comprehensively tune for every single combination of sample size, target dataset, and downstream task, complemented by the prior observation that deep survival models are notoriously challenging to train on heavily imbalanced prediction tasks [40] as is commonly the case here.

**GRU-D** by Che et al. [80] is a variant of the gated recurrent neural network architecture tailored for sparse and irregular sequence data, which explicitly learns exponential decay functions with a regression back to the mean in the absence of information—strong inductive biases for the domain of critical care data.

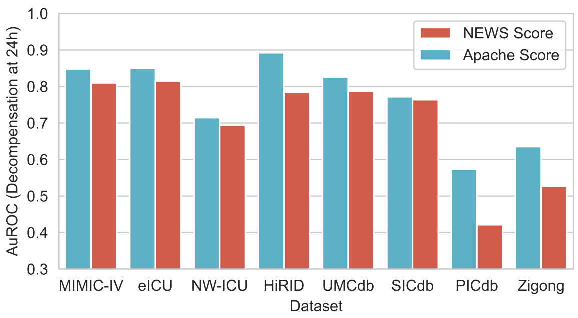


**Figure S7: Clinical Score Comparison for Mortality.** We compare the usage of either Apache-II[41] or NEWS[82] for predicting the onset of mortality at a 24-hour prediction horizon.

**StraTS** by Tipirneni et al.[81] uses a modern take on representing sparse multi-variate data in critical care and transforms the sparse multi-variate time-series into a one-dimensional sequence of measurements (i.e., "tokens") akin to modern language models. The work itself also proposes a forecasting based pretraining objective as a preliminary stage to pretrain the transformer before fine-tuning for a specific downstream task. Supplementary B.6 provides ample evidence for our choice of pretraining objective and the superior performance of time-to-event based pretraining for critical care data as opposed to optimizing a forecasting regression loss. However, we consider the StraTS architecture for its different approach to representing the sparse data and train it in a supervised fashion.

**Results and Conclusion** Figure S6 summarizes the comparative performance of DySurv, GRU-D, and STraTS against our best reference baseline (the per-configuration optimum of LightGBM and GRU) across all dataset–task–sample size combinations. The win rate analysis (Figure S6A) confirms that our best reference baseline dominates at moderate to large sample sizes, achieving win rates of 69% at 5,000 patients and exceeding 85% beyond 20,000 patients. At smaller sample sizes ($n \leq 500$), GRU-D and STraTS occasionally match or exceed the reference, though without consistent advantage.

The AuROC difference distributions (Figure S6B) reveal distinct performance profiles across the three architectures. DySurv exhibits substantial and consistent underperformance across all sample sizes, with median AuROC deficits of approximately 0.20, corroborating the joint optimization challenges noted above for imbalanced prediction tasks. GRU-D performs closest to the reference baseline, with median differences near zero and marginal advantages at small sample sizes ($n = 100$–$500$) that diminish as training data increases beyond 1,000 patients. STraTS shows intermediate behavior, with moderate deficits at small sample sizes that gradually converge toward the reference as more supervised data becomes available.

These findings validate our choice of optimally tuned LightGBM and GRU as strong reference baselines: none of the alternative architectures—despite incorporating domain-specific design choices for sparse and irregular time series—consistently outperform the carefully tuned reference models across the full range of experimental configurations. The best reference baseline thus provides a rigorous and conservative comparator for evaluating ICareFM's foundation model performance throughout this study, especially given that most LPE (local patient equivalence, Supplementary C.4) intersection points observed in the study occur beyond the 500 training (or fine-tuning) sample size.

#### C.3 Clinical score references

For each downstream task, we compute a reference performance using an established clinical score or relevant proxy:

- Decompensation (mortality): Apache-II[41] without the chronic health components (Figure S7 compares also against NEWS[82])
- Sepsis: full SOFA[31] score
- Circulatory failure: SOFA[31] cardiovascular component
- Respiratory failure: SOFA[31] respiratory component

- Kidney failure: SOFA[31] renal component
- Hyperglycemia: latest glucose measurement
- Liver failure: MELD score[27]

The scores are computed on the latest available forward-filled data at each time point.

#### C.4 Determining local patient (performance) equivalence (LPE)

Intensive care data shows strong distribution shifts across hospitals[18], which makes it challenging to outperform a locally trained model using only external data. However, foundation models are typically pretrained using external data and aim to show strong performance when used out-of-the-box or with minor adaptation to reduce the effective local model development and training time. Typically, when the unseen target hospital has large amounts of patient data available for developing its local model, state-of-the-art machine learning algorithms can achieve strong predictive performance. However, for smaller hospitals or hospitals in countries with stricter regulations on patient data usage for AI model training, using external data to achieve optimal performance can be key. The goal of the employed evaluation scheme is to identify the regimes in terms of available data in the target (local) hospital of interest.

Given the model tuning procedure established in Supplementary C.1 we obtain a performance curve showing how, for each dataset and downstream task, performance scales with increasing amounts of local (in-distribution) patient data.

We then perform zero-shot evaluations (no target data used) and adaptations of the foundation model (pretrained on external data) using different fine-tuning approaches with an increasing budget of target patients for adaptation. We compute the intersection of the mean performance curves of the foundation model and the prior computed best reference performances of models trained solely on data from the target hospital and trained using best practices for state-of-the-art machine learning model development. This yields a performance assessment expressed as *number of local patient performance equivalent* (short **LPE**). Unless stated otherwise, LPE is computed using AuROC as the performance metric. Conservatively, we report the first observed intersection between the two mean performance curves. Given an unseen target hospital and downstream task, if the available number of patients eligible for adaptation is lower than the reported number, then it is advisable to use the foundation model in its suggested mode at that sample size. If larger amounts of local training data are available, it might be advisable to develop a local task-specific solution if pure statistically observed performance is the main concern. However, development time, flexibility during deployment, and model robustness might still favor the use of the foundation model in one of its proposed deployment modes. When no intersection is observed within the available data, we extrapolate performance curves using a model logarithmic in sample size up to at most twice the maximum available training set size (see Supplementary C.4.1 for details). If the curves still do not intersect, the LPE is right-censored at the extrapolation limit, yielding a conservative lower bound. The extent to which this censoring affects aggregate LPE estimates is quantified in Supplementary C.4.3.

##### C.4.1 Performance curve refinement

The baseline tuning approach described in Supplementary C.1 yields for each downstream task and dataset a performance curve scaling with increasing training set size. Each training set size has three associated data points from three different random training runs.

However, due to the stochasticity in the training and model selection process these curves are not guaranteed to be monotonically increasing, even if they often are. To robustify the process of computing intersections of these scaling curves we perform the following procedure:

1. **Monotonisation.** For each seed we fit an isotonic regression through the data points of that specific seed. This yields three monotonically increasing curves (training set size vs. test set performance). Monotonicity ensures robust intersection matching between the reference and the foundation model scaling curves.
2. Then we report standard deviations and averages across the three seeds.
3. **Interpolation.** Each per-seed monotonised curve is interpolated onto a dense grid of 1,000 points using Piecewise Cubic Hermite Interpolating Polynomials (PCHIP)[83], which preserve monotonicity. Interpolation is performed in $\log_{10}(n)$ space (sample sizes are log-transformed before interpolation and back-transformed afterward), ensuring uniform resolution across orders of magnitude of training set size.

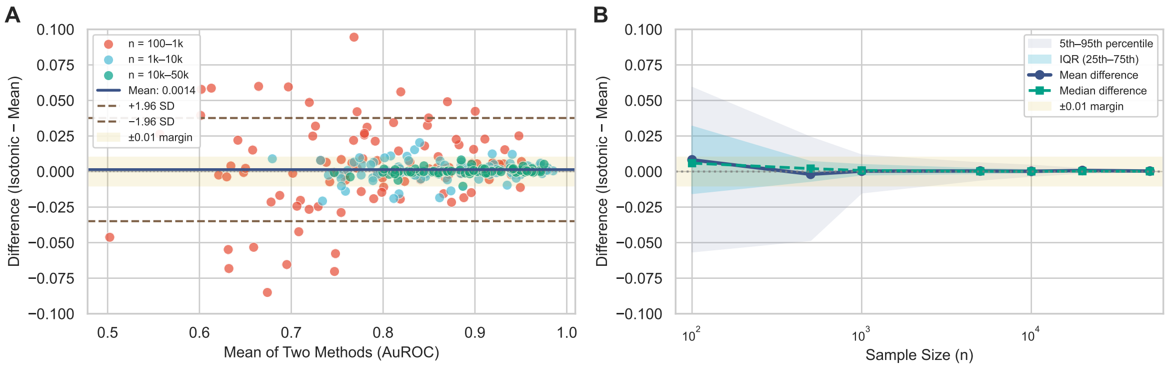


**Figure S8: Sensitivity analysis of isotonic regression.** **A)** Bland–Altman plot comparing monotonicity-enforced and raw seed-averaged AuROC values across 277 dataset–task–sample size combinations. Points are colored by sample size range. Horizontal lines indicate mean difference (blue) and 95% limits of agreement (brown dashed). The yellow shaded region indicates the $\pm 0.01$ equivalence margin. **B)** Mean difference (blue) and median difference (green dashed) across sample size, aggregated over 50 dataset–task combinations. Shaded bands show the interquartile range (dark) and 5th–95th percentile (light). Variability is largest at small sample sizes but converges to near-zero at $n > 10{,}000$, confirming that isotonic regression robustifies sample efficiency curves for reliable intersection computation without biased performance distortion.

4. **Extrapolation.** When the observed range does not reach the extrapolation bound (set to twice the number of labeled patients at each site and task), both the reference and ICareFM curves are extrapolated using a model logarithmic in sample size ($\hat{y}_{\text{perf}} = a \cdot \log_{10} n_{\text{train}} + b$), fitted by ordinary least squares on the last $100$ of the $1{,}000$ interpolated points. Logarithmic growth provides a conservative continuation of each scaling curve.

To assess whether isotonic regression introduces systematic bias, we compared the monotonicity-enforced estimates to raw seed-averaged values across 277 dataset–task–sample size combinations for AuROC test set performances (see Figure S8). A Bland–Altman analysis revealed negligible mean difference ($\Delta = 0.001$, SD $= 0.019$), with Lin's concordance correlation coefficient of $0.98$ indicating excellent agreement (Figure S8A). As expected, differences were larger at small sample sizes ($n < 1{,}000$: 42% within $\pm 0.01$ AuROC) where curve variability is highest, but virtually eliminated at larger sample sizes ($n > 10{,}000$: 99% within $\pm 0.01$ AuROC). Curve-level analysis across 50 dataset–task combinations confirmed this pattern: the mean absolute deviation between methods was $0.011$ AuROC (median: $0.009$), with both mean and median differences converging to zero as sample size increased (Figure S8B). This confirms that isotonic regression serves its intended purpose of robustifying curves for reliable intersection computation without biasing performance estimates across all the performed experiments considerably in any direction.

##### C.4.2 Statistical analysis

Performance metrics reported include the area under the receiver-operating-characteristic curve (AuROC), area under the precision–recall curve (AuPRC), and expected calibration error (ECE). Scaling analyses shown in the main text report means and one-standard-deviation bands across three random-seed runs. We computed 95% confidence intervals using bootstrap resampling: sampling across datasets and tasks for multi-dataset analyses and across patients for single-dataset analyses. Data from the Charité and RBKICU cohorts were used exclusively for validation and were not included in confidence interval estimates for cross-hospital generalization trends.

##### C.4.3 Right-censoring of LPE estimates

LPE estimates are right-censored when ICareFM outperforms the locally trained reference model at the maximum available training set size, because the true intersection of the two performance curves lies beyond the observed data range. To quantify this effect, we examined per-dataset censoring rates and their relationship to dataset size (Figure S9 A). The largest cohorts (eICU, MIMIC-IV) exhibit low censoring rates because sufficient local training data is available to observe the performance intersection. The highest censoring rates occur in mid-sized datasets where ICareFM transfers well (HiRID, UMCdb, NW-ICU, SICdb; censoring rates 60–85%): the model consistently outperforms the local reference at maximum available training size, but the datasets are not large

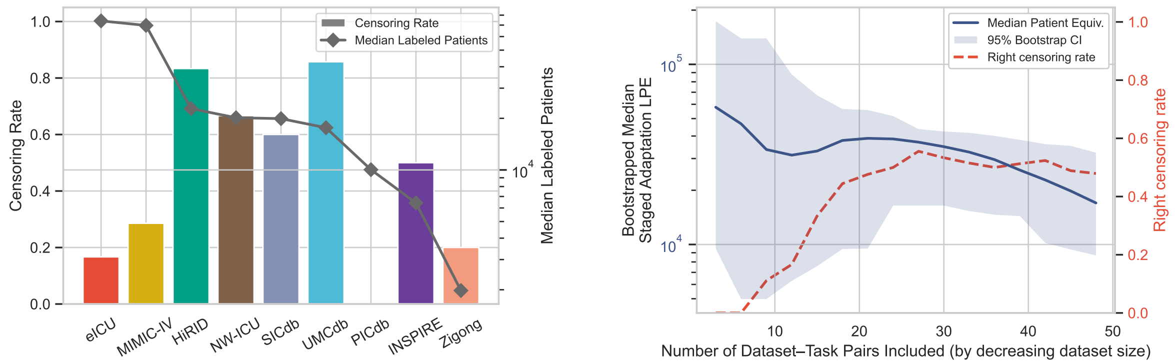


**Figure S9: Right-censoring of staged adaptation LPE estimates. A)** Per-dataset censoring rate (bars) and median number of labeled patients (diamonds, log scale), ordered by decreasing dataset size. The highest censoring rates occur in mid-sized datasets where ICareFM transfers well (HiRID, UMCdb, NW-ICU, SICdb) but available data is insufficient to observe the true performance intersection. Datasets with weaker transfer (e.g., Zigong) show low censoring despite small size. **B)** Bootstrapped median staged adaptation LPE (blue, left axis) and right-censoring rate (red dashed, right axis) as a function of the number of dataset–task pairs included in decreasing order of dataset size. The median LPE decreases and censoring increases as well-transferring but size-limited cohorts enter the aggregate, confirming that reported LPE statistics are conservative.

enough to reveal the true crossing point. In contrast, datasets where transfer is less effective (e.g., Zigong) show low censoring despite their small size, as the intersection is observed within the available data range.

When progressively including dataset–task pairs in decreasing order of dataset size (Figure S9 B), the bootstrapped median LPE decreases while the right-censoring rate increases, reflecting the entry of these well-transferring but size-limited cohorts into the aggregate. This confirms that the LPE statistics reported in the main text are conservative lower bounds of ICareFM's true patient equivalence.

##### C.4.4 LPE sensitivity to choice of evaluation metric

Throughout the main text, LPE is computed using AuROC as the default performance metric. To assess whether this choice materially affects the conclusions, we repeat the LPE analysis using two alternative families of evaluation metrics: the area under the precision–recall curve (AuPRC) and event-level recall at fixed false positive rates (EventEvaluation@FPR/recall).

**AuROC vs. AuPRC.** Figure S10 compares LPE distributions across all six deployment modes when computed under AuROC and AuPRC. For zero-shot modes and clinical scores, AuROC-based LPEs are moderately higher than their AuPRC counterparts. This is consistent with AuROC being rank-based and therefore invariant to calibration, whereas AuPRC is sensitive to the alignment between predicted probabilities and local prevalence, which is a property that penalizes zero-shot models that have not been calibrated on local data. Conversely, for adaptation-based modes (external, local, and staged adaptation), AuPRC-based LPEs exceed their AuROC counterparts, indicating that adaptation not only improves discrimination but also calibrates the model to specific outcomes and local data, yielding precision–recall gains that shift the local scaling curve intersection further and produce even larger LPEs than AuROC alone would suggest. Importantly, however, LPE values remain comparable in order of magnitude across both metrics. The relative ordering of deployment modes is fully preserved. This consistency indicates that LPE provides a stable quantification of data efficiency that is not an artifact of the AuROC metric.

**Event-level recall at fixed FPR.** To further probe metric sensitivity under clinically relevant operating conditions, we compute LPE using event-level recall evaluated at seven fixed false positive rate thresholds ranging from $10^{-4}$ to $10^{-1}$ (Figure S11), deployment settings with low false alarm rates optimized for an autonomous alarm system. This family of metrics directly measures how well a model detects clinical events at a specific alarm rate.

For the dual zero-shot mode, median LPE increases monotonically as the FPR budget grows from $10^{-4}$ to $10^{-1}$, rising from approximately 100 to 300 patient equivalents. This pattern is expected: at very strict operating points (low FPR), the achievable recall is inherently limited for both models, leaving less room for the foundation model to differentiate itself from a local reference trained on modest amounts of data. At more permissive alarm

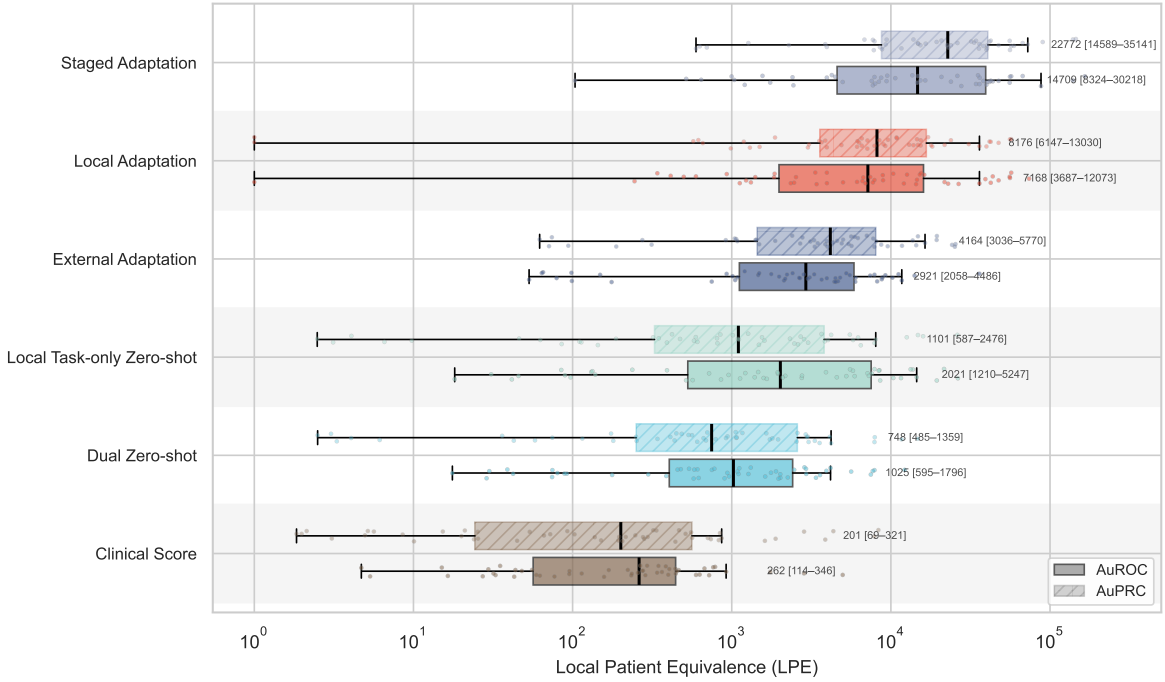


**Figure S10: Local patient equivalence (LPE) based on AuROC or AuPRC** For each deployment mode, LPE is computed using either AuROC (solid fill) or AuPR (hatched fill) as the underlying performance metric. Boxplots summarize the distribution across all dataset–task combinations ($n = 50$), with individual points overlaid. Annotations show the median and bootstrapped 95% confidence interval. LPE values of zero (cases where the method does not exceed the local reference curve) are clamped to 1 for display on the log scale. LPE values remain comparable in scale across both metrics, indicating that the measure provides a consistent quantification of data efficiency regardless of the underlying evaluation criterion.

rates (higher FPR), the foundation model's broad pretraining allows it to sustain higher recall relative to the local baseline, pushing the performance intersection further along the local scaling curve and thus increasing the LPE. Clinical scores are largely unable to operate at such low false positive rates altogether, as their coarse discrete outputs lack the precision required to maintain meaningful recall at strict alarm budgets.

For the adaptation-based modes, LPE remains largely stable across FPR thresholds. External and staged adaptation, which both use labeled data from external hospitals, maintain median LPEs near the AuROC and AuPRC reference lines across all operating points. Local adaptation, which relies solely on local labels, falls consistently below both reference lines, reflecting greater sensitivity to the evaluation metric when no external supervision is available. This likely reflects that local adaptation and the local reference are trained on the same labeled data distribution, yielding scaling curves that lie closer together and whose intersection is therefore more sensitive to the choice of evaluation metric. External and staged adaptation, by incorporating external labeled data, establish a performance margin that is preserved across metrics and operating points. Overall, the scale of the LPE value conferred by adaptation is stable across clinically relevant operating points.

**Summary.** These analyses confirm that the LPE framework yields consistent and interpretable results across a broad range of evaluation metrics. The relative ranking of deployment modes, the order of magnitude of patient equivalents, and the qualitative conclusions drawn in the main text are not sensitive to whether discrimination is measured by AuROC, AuPRC, or even event-level recall at fixed false positive rates. We retain AuROC as the default metric throughout the main text because, unlike AuPRC and recall at fixed FPR, it is invariant to event prevalence, which is an important property given the substantial variation in label rates across tasks, datasets, and training set sizes in our evaluation protocol. This ensures that the resulting LPE estimates reflect the model's transferable discriminative performance.

#### C.5 Complementary task zero-shot results with domain adaptation

**Domain Adaptation during Pretraining** In Figure S12 we show additional zero-shot results complementary to the results presented in Figure 2. The Figure S12 A) and B) results show task zero-shot results when the

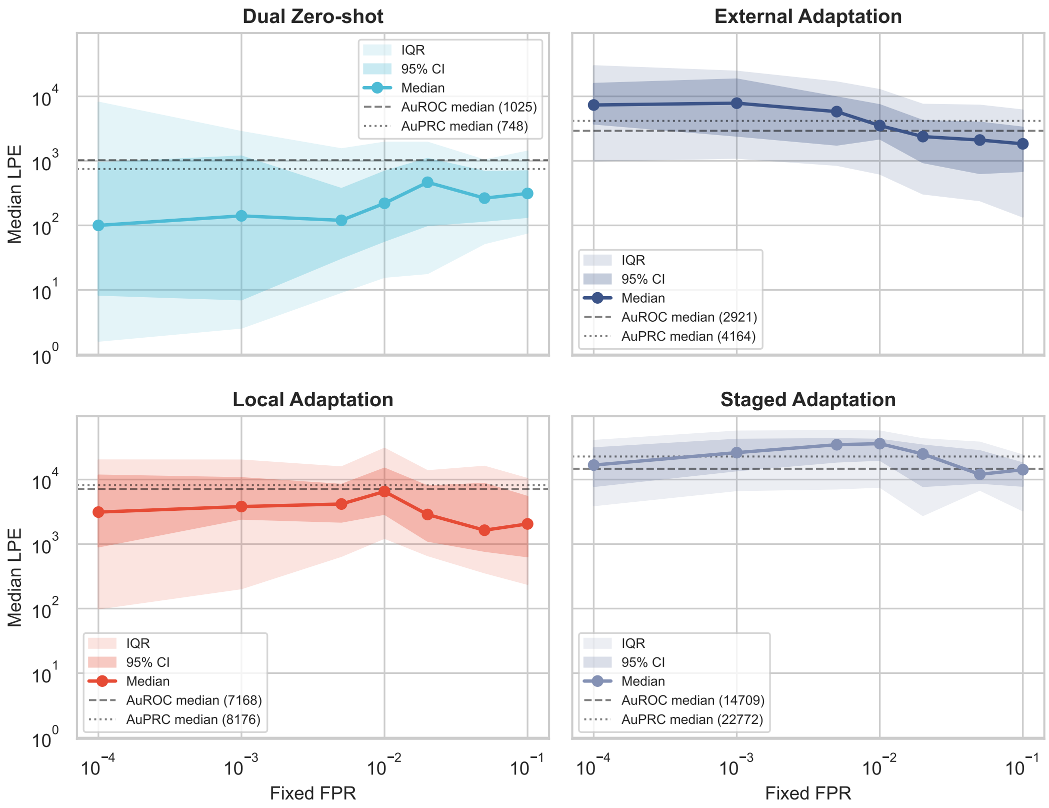


**Figure S11: Local patient equivalence (LPE) based on event-level recall at fixed false positive rates.** For each deployment mode, LPE is computed using event-level recall at a fixed FPR threshold as the underlying metric, with the FPR varied from $10^{-4}$ to $10^{-1}$ along the $x$-axis. Shaded bands show the interquartile range (light) and bootstrapped 95% confidence interval (dark) across all dataset–task combinations. Dashed and dotted horizontal lines indicate the median LPE obtained when using AuROC and AuPR as the evaluation metric, respectively.

model's pretraining was continued on the local target's training dataset (domain adaptation during pretraining).

**Scaling properties of ICareFM** In **Figure S12 C), D)** we analyse different scaling properties of ICareFM. Figure S12 C) provides empirical evidence that scaling the pretraining dataset of ICareFM can increase its zero-shot performance in new hospitals. We observe an approximate square root relationship, where quadrupling the external pretraining data doubles the number of patients required in the target to match ICareFM's zero-shot performance. Generously extrapolating by two orders of magnitude (an amount of data available to market leading electronic health record system vendors[84]), we predict that ICareFM could in expectation outperform local models trained with over $11{,}984\,[5{,}143–28{,}143]$ patient stays. This extrapolation suggests substantial gains from additional pretraining data, particularly given the flexibility of zero-shot out-of-domain prediction. Figure S12 D) suggests that ICareFM's performance implications slightly increase as we aim to predict events further ahead of time.

**Ablating the impact of rare events** In Figure 2 C) we analyse the relative performance implications of ICareFM with respect to the prevalence (rarity) of the target events. We modulate clinical event definitions according to the definitions described in Supplementary A and hence obtain for each dataset and task multiple severities of the same target organ failure with different prevalences and perform the LPE analysis. We observe increased relevance of ICareFM for rarer events benefiting from large scale data collection to observe larger numbers of rare events.

**Comparison with clinical scores** In dual zero-shot mode, ICareFM outperformed commonly used ICU clinical scores, achieving a median AuROC improvement of $+0.049$ ($95\%$ CI, $0.031$–$0.071$). Using the LPE framework, clinical scores achieve an LPE of approximately $261$ locally labeled patients.

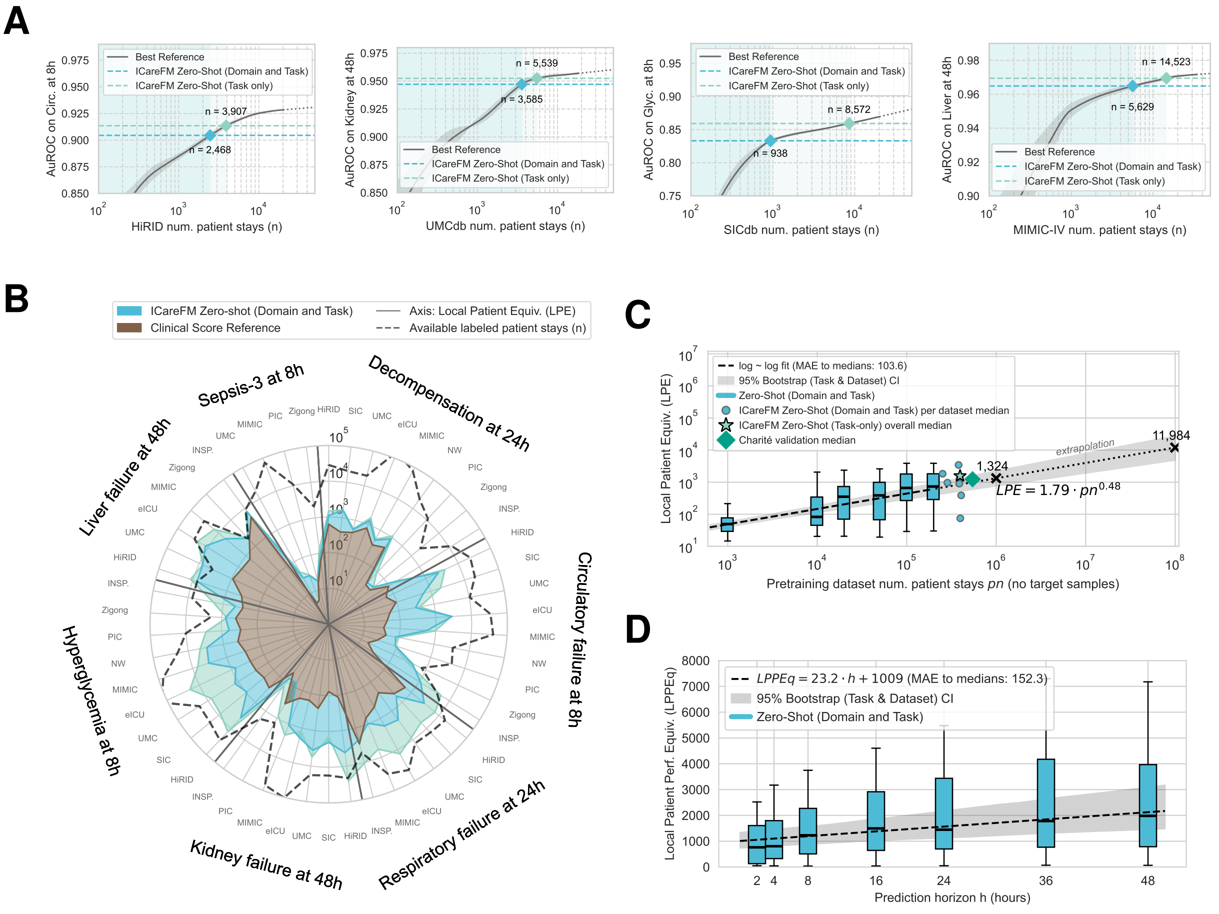


**Figure S12: Zero-shot Capabilities A)** Sample scaling studies with zero-shot ICareFM performances including performance of a model where pretraining was continued on the local dataset (domain adaptation during pretraining). **B)** Scaling study summary over tasks and datasets comparing against clinical scores including performance of a model where pretraining was continued on the local dataset (domain adaptation during pretraining). **C)** Scaling law: Growing the pretraining dataset of ICareFM increases zero-shot performance in new hospitals. **D)** Performance scaling behavior with increasing prediction horizon.

#### C.6 Language model experiments

In the following we provide details on the LLM integration experiments both for the tool-calling approach to integrate ICareFM and the direct zero-shot prediction of LLMs on critical care time series for early event predictions. Note that at no point any LLM was trained or adapted in any way on any data from the harmonized critical care cohorts. These results have been obtained with fixed API or local open-weight models without adaptation.

##### C.6.1 ICareFM tool calling LLM integration

Beyond direct prediction, we evaluated an alternative approach where large language models configure and invoke ICareFM's zero-shot prediction capabilities. This tool-calling approach uses LLMs' natural language understanding to translate clinical queries into structured prediction requests, while delegating the actual risk estimation and temporal extrapolation to the specialized time-series foundation model.

**Motivation.** Clinicians benefit from expressing prediction requests in familiar terminology rather than specifying variables, thresholds, and horizons directly. However, general-purpose LLMs lack the specialized training for accurate clinical risk estimation from time-series data. Tool calling addresses this by separating the two roles: the LLM interprets clinical intent and configures the prediction, while ICareFM executes risk estimation using its pretrained survival heads.

**System Architecture.** The tool-calling interface positions the LLM as an abstraction layer between the clinician and ICareFM. Given a natural language query describing a clinical outcome of interest (e.g., "risk of circulatory failure in the next 8 hours"), the LLM generates a structured configuration that specifies which ICareFM prediction heads to invoke and how to combine their outputs. For each task we design three different queries.

**Prompt Design.** The LLM receives a system prompt establishing its role as an interface to a time-series foundation model trained for survival prediction. The prompt provides:

1. A list of available clinical variables with their abbreviations and measurement units (e.g., blood lactate in mmol/L as `lact`, mean arterial pressure in mmHg as `map`)
2. Instructions for constructing boolean expressions in disjunctive normal form (DNF) to specify complex clinical endpoints
3. Examples demonstrating how to translate clinical concepts into prediction configurations

The prompt instructs the LLM to reason through the clinical problem systematically: identifying relevant variables, determining appropriate thresholds based on clinical guidelines and published literature, selecting prediction horizons, and constructing the boolean expression that captures the desired clinical outcome. Note that in this setting the LLM never actually sees patient data, this is purely about translating a query to a probabilistic formula that invokes ICareFM's prediction heads.

**Probability Aggregation.** Given the boolean expression from the LLM, ICareFM computes cumulative failure probabilities $F_k(h \mid \mathbf{H}_t, \tau, \delta)$ for each atomic condition and aggregates them according to the logical structure:

- **Conjunction (AND):** For conditions within a clause, probabilities are multiplied assuming conditional independence:
$$P(\text{clause}) = \prod_{i \in \text{clause}} F_{k_i}(h_i \mid \mathbf{H}_t, \tau_i, \delta_i) \tag{14}$$
- **Disjunction (OR):** Across clauses, the union probability is computed as:
$$P(\text{event}) = 1 - \prod_{j \in \text{clauses}} (1 - P(\text{clause}_j)) \tag{15}$$

The current tool-calling implementation focuses exclusively on threshold-based probability combinations using these aggregation rules. ICareFM additionally supports expected value forecasting for clinical scores (e.g., APACHE, MELD) by integrating predicted probability distributions across discretized value bins (Supplementary B.2). Future extensions of the tool-calling interface could incorporate these forecasting capabilities, enabling LLM-configured predictions of continuous clinical scores rather than solely binary threshold-crossing events.

**Threshold Transformation.** Thresholds specified by the LLM in clinical units are transformed to the standardized space used during ICareFM pretraining using the same scalers applied during data preprocessing. Values are clipped to the range $[-4, 4]$ in standardized space to remain within the model's trained distribution. This transformation ensures that clinically meaningful thresholds (e.g., lactate $> 2$ mmol/L) are correctly mapped to the model's internal representation.

**Evaluation Protocol.** We evaluated the tool-calling approach on the same subsampled test sets used for direct LLM prediction (Supplementary C.6.2). For each clinical prediction task, we provided the LLM with a natural language description of the target endpoint and evaluated whether the generated configuration produced accurate risk estimates when executed by ICareFM.

This controlled comparison isolates the effect of the prediction approach: we perform direct LLM prediction versus LLM-orchestrated tool calling, while holding constant all other factors including the chosen LLM, test data, and evaluation metrics. Results are presented in Figure 5 and Figure S14.

**Practical advantages.** Once the LLM generates a configuration (a single API call per query type), ICareFM executes predictions for all patients and time points at orders-of-magnitude lower latency and cost than per-prediction LLM inference. The explicit configuration also provides transparency into which variables, thresholds, and logical relationships are used, and clinicians can review or modify it before execution.

**Limitations.** The tool-calling approach depends on the LLM's ability to correctly translate clinical intent into appropriate variable selections, thresholds, and logical structures. Errors in configuration, such as selecting inappropriate thresholds or omitting relevant variables, propagate to prediction quality. Additionally, the current implementation is constrained to endpoints expressible as boolean combinations of threshold-crossing events on the variables available in ICareFM's prediction head set. Complex endpoints requiring reasoning about temporal patterns, rate-of-change dynamics, or variables not included in the model's training cannot be directly addressed through this interface without further extension of the tool capabilities.

##### C.6.2 LLM zero-shot reference baselines

To contextualize ICareFM's zero-shot prediction capabilities against contemporary large language models (LLMs), we evaluated direct clinical event prediction using state-of-the-art LLMs in a purely zero-shot setting without task-specific fine-tuning or in-context learning examples.

**Motivation and Scope.** This evaluation serves two purposes: (1) establishing whether general-purpose LLMs can perform competitive zero-shot clinical risk prediction when provided with time-series patient data (or structured features thereof), and (2) providing a reference point for the tool-calling integration approach where LLMs orchestrate tool calls to ICareFM rather than performing predictions directly themselves. We evaluated all task-dataset combinations for which ground-truth labels were available, spanning the clinical endpoints described in Supplementary A.5.

**Models Evaluated.** We evaluated a diverse set of large language models spanning both proprietary and open-source architectures:

- **Proprietary models:** We tested GPT-5 variants in the full base model and its more compact `mini` and `nano` variants. `gpt-5-2025-08-07` to be explicit, being the latest model when these experiments were conducted. Accessed via a dedicated Azure hosted model deployment.
- **Open-source models:** We run inference on a collection of open-source models either locally using Nvidia RTX 4090 or Nvidia H200 GPUs or through the Swiss AI Model Serving Infrastructure[3]. We considered: Gemma-3 variants[85] (4B and 27B parameters), Llama 3.1 8B[86], and GPT-OSS variants[87] (20B and 120B).

[3] `https://serving.swissai.cscs.ch`

**Computational Considerations and Subsampling.** Running LLM inference for clinical event prediction at every hourly time point across complete ICU test sets proved computationally prohibitive both in terms of computational resources (and time) and financially (due to closed-source model API costs). For a single task-dataset combination, evaluating all eligible time points would require hundreds of thousands of API calls, rendering full evaluation infeasible given cost and time constraints.

We therefore subsampled each test set to 500 patients per task-dataset combination: up to 150 patients with at least one positive event label within the first 72 hours (or as many as available), and 350 patients without any positive labels in this window (or more if less than 150 patients were sampled for the positive cases). For the experiment on the full GPT-5 model we further subsampled to 50 patients with events and 100 without for a total of 150 per task-dataset combination. This stratified sampling ensures adequate representation of both outcome classes while reducing computational burden. For each sampled patient, we evaluated predictions at all available time points up to hour 72, still resulting in several thousand predictions per task and dataset.

Beyond cost considerations, this experiment highlights a fundamental limitation of LLM-based approaches for real-time ICU monitoring: generating predictions at regular intervals (e.g., hourly) for all patients in an intensive care unit would require substantial computational infrastructure, electricity consumption, and ongoing API costs. While technically feasible, deploying large language models as real-time clinical predictors remains impractical for routine clinical use compared to specialized foundation models that can generate predictions with orders-of-magnitude lower latency and resource requirements.

**Prompt Design.** Each prediction request consisted of two components:

*System prompt.* We established the model as an expert ICU physician with extensive experience in clinical risk assessment, familiar with standard severity scoring systems (APACHE, SOFA, SAPS, MELD). The system prompt instructed the model to analyze provided clinical data, identify concerning patterns, and return a JSON object containing a single probability estimate (0–100%) for the specified clinical event.

*User prompt.* Task-specific templates specified the clinical event definition and prediction horizon. For example, circulatory failure predictions used the prompt: "Given the following patient vital signs and clinical data: {data}. Predict the probability (0–100%) that this patient will experience circulatory failure within the next 8 hours."

**Clinical Data Encoding.** We transformed the high-dimensional feature vectors (identical to those used for gradient boosted tree baselines; see Supplementary A.4.4) into human-readable clinical summaries optimized for language model comprehension.

For each clinical variable, we computed summary statistics over the preceding 24-hour window and presented them as an 8-point tuple: (last known value, minimum, 10th percentile, median, 90th percentile, maximum, mean, linear trend slope). The trend slope was computed in standardized space to provide scale-invariant trend information. Variables with entirely missing observations within the 24-hour window were excluded from the prompt to reduce context length and avoid misleading the model with placeholder values.

Crucially, all continuous feature values were inverse-transformed from standardized space back to original clinical units using the same preprocessing scalers applied during data normalization. This ensures the model receives physiologically interpretable values (e.g., heart rate in beats per minute, creatinine in mg/dL) rather than z-scored representations. Categorical variables (e.g., admission type, care unit) were decoded from integer indices to their original clinical labels. Units were provided as part of the prompt for each variable.

**Inference Configuration.** For the GPT-5 model family, we employed reduced reasoning effort settings to manage costs: GPT-5-nano and GPT-5-mini used "low" reasoning effort, while GPT-5 (full model) used "minimal" reasoning effort. We acknowledge that these settings may slightly reduce model capabilities compared to default configurations, but evaluation at full reasoning capacity was cost-prohibitive.

API calls were executed asynchronously with rate limiting, automatic retry with exponential backoff for transient failures, and incremental checkpointing to enable resumption of interrupted evaluation runs. Open-source models were deployed locally on hardware ranging from consumer-grade NVIDIA RTX 4090 GPUs to datacenter-class NVIDIA H200 accelerators, depending on model size requirements. Proprietary GPT-5 variants were accessed through secure Azure-hosted endpoints.

**Response Parsing.** Model outputs were parsed to extract probability estimates using a multi-stage approach: (1) attempting JSON parsing for structured responses, (2) regex extraction of numeric values for unstructured responses. Probabilities expressed as percentages (0–100) were normalized to the $[0, 1]$ interval for evaluation.

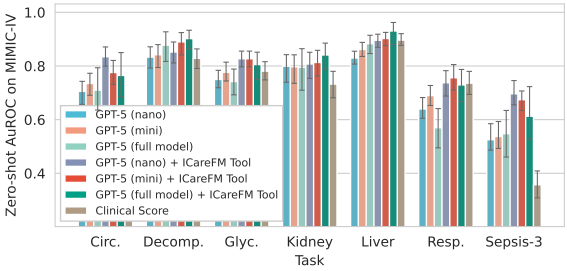


**Figure S13: GPT-5 native LLM and ICareFM tool call experiments.** Comparing early event prediction performance of GPT-5 variants. We compare the LLM's native prediction capabilities by encoding the clinical data into the prompt with orchestrating a query to ICareFM to process the time series data to assist in answering the query. Error bars show 95% confidence intervals across patient stay bootstraps.

**Evaluation Protocol.** LLM predictions were evaluated predominantly using AuROC due to being less sensitive to prevalence shifts in the data efficient subsample used for the LLM experiments. Performance is compared against ICareFM zero-shot predictions where the zero-shot configuration prompt for ICareFM itself was performed by the same LLM (Supplementary C.6.1). Results are presented in Figure 5 and Figure S13.

**Limitations.** This evaluation represents a purely zero-shot approach. The reduced reasoning effort settings for GPT-5 variants and the subsampled evaluation sets may underestimate optimal LLM capabilities. However, given the small performance differences even between the full GPT-5 model and, for example, the mini variant, we do not anticipate improvements that would affect our study's conclusions. Additionally, our structured tabular data representation may not optimally exploit language models' strengths compared to narrative clinical notes or purpose-built multimodal architectures. However, they represent the reality of this large amount of available information, which needs to be processed to perform accurate and timely predictions.

These limitations do not compromise the validity of our comparative analysis. The primary comparison of interest is between LLM direct prediction (evaluated here) and LLM-orchestrated tool calls to ICareFM (Supplementary C.6.1), where the language model serves as an interface layer that interprets clinical context and invokes the foundation model for prediction. This controlled comparison isolates the question of whether LLMs are more effective as direct clinical predictors or as intelligent orchestrators of specialized foundation models. This is independent of absolute LLM capability levels that might be achieved with unlimited computational budgets.

These results should therefore be interpreted as establishing the relative merits of direct LLM prediction versus tool-augmented prediction within practical deployment constraints, rather than as a definitive assessment of maximal LLM capabilities for clinical risk assessment.

##### C.6.3 Complementary LLM results

Considering results on open-weight models (Figure S14 ICareFM significantly improves early event prediction performance (AuROC $+0.1428\,[95\%\,CI\,:\,0.1311-0.1546]$ across models, tasks, and datasets comparing native LLM prediction with ICareFM tool calls). We do observe a trend that larger models achieve better performance on direct zero-shot time-series predictions up to model sizes of 20B to 30B parameters. However, when comparing the achieved performance to clinical scores, the native LLM approach often falls short or only marginally improves performance. Especially noteworthy is the effective tool calling capabilities of even smaller models such as the 4 billion parameter version of Gemma-3 (Tool calling AuROC: $+0.0621\,[95\%\,CI\,:\,0.0259-0.1009]$ over clinical scores $P<0.001$) or also the 27 billion parameter Gemma-3 (Tool calling AuROC: $+0.0637\,[95\%\,CI\,:0.0313-0.0974]$ over clinical scores) and 20 billion parameter GPT-OSS (Tool calling AuROC: $+0.0763\,[95\%\,CI\,:0.0495-0.1060]$ over clinical scores). These models can be run locally inside a hospital without the large computational, cost, or privacy concerns of using a closed-source frontier model. The LLM provides an intuitive natural language interface and ICareFM ensures a much more robust prediction and efficient processing of the time series data. Most interestingly MedGemma 27B is not significantly better at tool calling ICareFM (AuROC difference $0.0064\,[95\%\,CI\,:-0.0176-0.0284]$, $P=0.3001$). While MedGemma has been tuned for medical tasks, the adaptation most likely focused less on time-series data rather than clinical notes as well as imaging modalities, potentially explaining the small drop in native LLM time-series

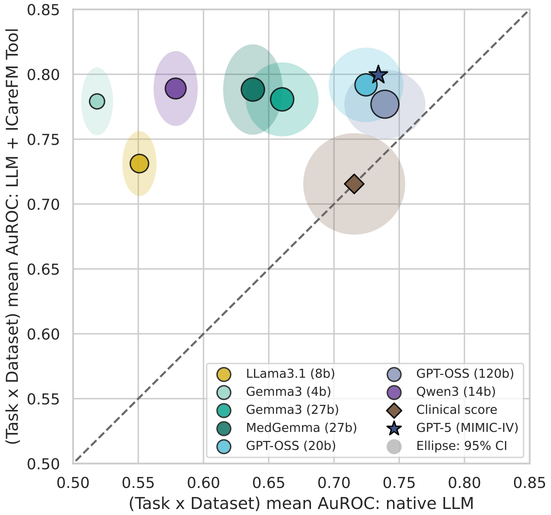


**Figure S14: LLM Native vs. ICareFM tool calls** Comparing native LLM prediction performance vs. using ICareFM as an agentic tool for LLMs. Uncertainty regions show 95% confidence intervals on the mean by bootstrapping across dataset and task pairs.

processing capabilities (dropping by $0.0223\,[95\%\,CI : 0.0107 - 0.0340]$ AuROC).

#### C.7 Survival head evaluation

##### C.7.1 Univariate survival head evaluation

We evaluated the *survival head* of ICareFM as a flexible time-to-event predictor for clinically meaningful threshold-crossing events on routinely measured variables. Given a target variable (e.g., lactate) and an event definition parameterized by a z-score threshold $\tau$, we define an *event* at time $t$ if the variable crosses the threshold at that time point. At each time point $t$, the model outputs the hazard predictions and the cumulative failure probability for each target variable conditioned on a chosen threshold. To ensure the event labels reflect observable outcomes, we censor time points for which no *raw* measurement of the target variable is available anywhere in the prediction horizon.

We report discrimination and calibration-aware performance using *time-dependent* Dynamic AUC and the *integrated Brier score* (IBS). Dynamic AUC is computed over evaluation times $\Delta \in \{1, \dots, H\}$ using the predicted cumulative failure probabilities as risk scores, and IBS integrates the Brier score across the same times (lower is better). We compare against a Cox proportional hazards baseline (elastic-net regularized) fit separately for each variable and threshold $\tau$ using the same feature representation at time $t$. Unless otherwise stated, results aggregate over all dataset–variable–threshold–scenario combinations, with uncertainty estimated by pooling across these units. The Cox model baseline should help to understand trivially achieved predictive performance and highlights ICareFM's strong performance even without using any data from the target hospital across a large range of survival predictions.

Across all comparisons (Fig. S15), ICareFM substantially outperforms Cox in both discrimination (higher Dynamic AUC) and accuracy (lower IBS). We evaluate two ICareFM settings: a *zero-shot* model pretrained on external data (from different hospitals) and applied directly to a target dataset (out-of-distribution, OOD), and an *adapted* model that continues pretraining on in-distribution (ID) data after the large external pretraining stage. Note this model is not tuned for the specific variable and threshold target but merely continues the self-supervised survival objective on data from the same hospital. This ablates the performance gap created by the data distribution shift of the target hospital compared to the external pretraining dataset. Paired comparisons show consistent gains of ICareFM over Cox, and additional (typically smaller) improvements from adaptation over zero-shot.

We next characterize robustness to the event definition and forecasting horizon (Fig. S16A). ICareFM maintains an advantage over Cox across clinically relevant prediction horizons and over a wide range of threshold severities, indicating that benefits are not driven by a particular operating point. Finally, forest-plot summaries show improvements are broadly distributed across datasets (Fig. S16B) and clinical variables (Fig. S17–S18), with particularly strong gains for several high-impact laboratory and vital-sign targets. Overall, these results suggest that the ICareFM survival heads show strong predictive performance pretrained from external data easily outperforming a baseline model using large amounts of local data. These evaluations underpin the presented zero-shot early event prediction results on more complex organ failure endpoints in Figure 2 and support the broad applicability of ICareFM across arbitrary clinical endpoints.

##### C.7.2 Bivariate vs. independence inference comparison

While Section B.8 demonstrates that including bivariate events during *pretraining* improves zero-shot transfer, this section evaluates the *inference-time* choice between two approaches for predicting joint threshold-crossing events:

- **Independence (Product of Marginals):** $F_{\text{Joint}}^{\text{Indep.}}(h) = F_{k_1}(h \mid \mathbf{H}_t, \tau_1) \cdot F_{k_2}(h \mid \mathbf{H}_t, \tau_2)$
- **Joint (Bivariate Head):** Direct prediction via the shared interaction head $\lambda_{\text{Bivariate}}$

We evaluate on three composite events: circulatory failure (MAP $< 65$ mmHg $\wedge$ lactate $> 2.0$ mmol/L), respiratory failure ($SpO_2$ $< 90\% \wedge$ $FiO_2$ $> 60\%$), and kidney dysfunction (creatinine $> 2.0$ mg/dL $\wedge$ urine rate $< 0.5$ mL/kg/hr). Performance is measured using mean time-dependent AuROC and integrated Brier score across nine ICU datasets in both zero-shot and adapted settings (pretraining continued on the target dataset).

**Results and Analysis** Counterintuitively, the product of marginals consistently outperforms the bivariate head in more than $75\%$ of evaluation scenarios (Figure S19). We attribute this to several factors:

- **Signal Sparsity:** Joint events are inherently sparser than univariate constituents (when two variables must simultaneously exceed thresholds), event rates decrease multiplicatively, substantially reducing training signal for the bivariate head.
- **Shared Head Bottleneck:** To maintain scalability, we employ a single shared bivariate head for all $\binom{K}{2}$ variable combinations rather than dedicated heads. This architectural constraint prevents optimal specialization for any particular variable pairing, whereas each univariate head fully specializes to its target.
- **Implicit Correlation Capture:** The sequence encoder produces state representations $\mathbf{H}_t$ that already capture cross-variable correlations. Univariate heads conditioning on this shared representation implicitly account for correlations, and their product may approximate joint distributions more accurately than a capacity-constrained bivariate head.
- **Limited Practical Scope:** Most clinically-relevant composite events cannot be expressed using bivariate thresholds alone. The full circulatory failure definition additionally considers vasopressor administration[17] or KDIGO criteria for acute kidney injury involve complex multi-window logic on creatinine trajectories and cumulative urine output[26]. Given that practical deployment requires the independence approach for these higher-order events regardless, the marginal performance gap for the bivariate-expressible subset has limited practical consequence.

**Recommendation** For composite event prediction, practitioners should use the product of marginals approach. While bivariate pretraining remains beneficial for representation learning (Section B.8), the well-trained univariate heads provide superior inference-time performance and naturally extend to arbitrarily complex event definitions beyond the bivariate case. Future work should investigate scalable architectures for joint prediction heads that can achieve sufficient specialization without requiring $\mathcal{O}(K^2)$ dedicated parameters.

#### C.8 Data missingness sensitivity analysis

Clinical datasets exhibit substantial heterogeneity in recording practices and data completeness across institutions. To assess whether data density—the proportion of observed versus missing values—influences foundation model performance, we conducted a post-hoc analysis examining the relationship between dataset completeness and zero-shot patient equivalence performance.

##### C.8.1 Methodology

We quantified data density for each dataset by calculating the proportion of non-missing values. For each dataset, data density was computed as:

$$\text{Data Density} = 1 - \frac{\sum_{i,j} \mathbb{1}[\text{missing}(x_{ij})]}{N \times P} \tag{16}$$

where $N$ denotes the number of hourly observations, $P$ represents the number of included variables, and $\mathbb{1}[\text{missing}(x_{ij})]$ is an indicator function for missing values.

We then computed the LPE performance estimates (see Supplementary C.4) across all clinical tasks benchmarked for each dataset under two evaluation paradigms: out-of-distribution (OOD), where the test hospital dataset was excluded from pretraining, and in-distribution (ID), where training data from the target hospital was used to continue pretraining. Linear regression with ordinary least squares estimation was performed to quantify the relationship between data density and patient equivalence, with 95% confidence intervals for the mean prediction calculated using the $t$-distribution.

Further results on local adaptation and staged adaptation revealed similar but weaker trends, suggesting that even if the model is adapted with local labeled data, adaptation is more successful if local data is dense and of high quality.

##### C.8.2 Results

Data density varied substantially across datasets, ranging from 1.7% (Zigong) to 15.8% (UMCdb) on the selected variable subset, reflecting institutional differences in monitoring intensity and data recording practices also highlighted in Tables S1 and S2. Linear regression revealed a positive correlation between data density and zero-shot performance all evaluation settings with significant p values for the slope of the linear fit in both zero-shot settings and an almost significant result in the staged adaptation setting (see Figure S20).

For out-of-distribution evaluation, the relationship was evaluated with $R^2 = 0.5$, $p = 0.032$), indicating a moderate and statistically significant association.

For in-distribution evaluation, a slightly stronger correlation emerged: $R^2 = 0.53$, $p = 0.026$), suggesting that data density has a more pronounced effect when the test distribution is represented during pretraining since we observe a much steeper slope of approx. $47{,}000$ compared to $13{,}239$ in the dual zero-shot (OOD) setting.

Similar but weaker trends are observed when local data is used to finetune the foundation model in Figure S20 B).

##### C.8.3 Interpretation

These findings suggest that data density moderately influences foundation model performance, with stronger effects observed for in-distribution zero-shot evaluation than for cross-site generalization and weaker results for adapted models.

The steeper relationship in the ID setting likely reflects two complementary mechanisms: (1) denser datasets provide richer training signals during pretraining for adaptation to the target, and (2) distributional alignment between pretraining and evaluation reduces the domain shift that must be overcome during zero-shot inference if large parts of the pretraining corpus exhibit denser measurement patterns.

The relatively modest $R^2$ values indicate that data density explains only a fraction of performance variability across datasets, with other factors such as patient case mix, clinical protocols, temporal resolution, and dataset size, likely playing important roles.

#### C.9 Zero-shot OOD calibration

Beyond discriminative performance, calibrated probability estimates are essential for clinical decision-making. We evaluated the calibration of zero-shot predictions across clinical tasks and external datasets without any task-specific or site-specific training or calibration adjustments.

**Classification Tasks.** Figure S21 A) shows the Expected Calibration Error (ECE) and reliability diagrams for seven early event prediction tasks. Calibration performance varied considerably across tasks: circulatory failure, respiratory failure, and hyperglycemia predictions exhibited the lowest ECE, indicating quite well-calibrated probability estimates. In contrast, decompensation and sepsis-3 predictions showed higher calibration errors, likely reflecting greater heterogeneity in outcome definitions and prevalence across centers. Despite the observed deviations, some tasks show reasonable calibration without requiring post-hoc recalibration, suggesting that the foundation model's pretrained representations transfer well to unseen clinical contexts. This result is encouraging as our study also suggests an improvement in calibration if the model is further scaled and trained on an ever growing dataset (Figure 3).

**Regression Task.** MELD scores are computed as $\text{MELD} = 9.57 \cdot \ln(\text{Creatinine}) + 11.2 \cdot \ln(\text{INR}) + 3.78 \cdot \ln(\text{Bilirubin}) + 6.43$, where individual values are clipped below at $1.0$ to avoid negative contributing factors after applying the logarithm. When we use MELD score forecasts scaled to a probability range in $[0, 1]$ to perform zero-shot liver dysfunction predictions, the clipping is responsible for the overconfidences seen in Supplementary C.9 A).

To assess the actual continuous zero-shot prediction capabilities and avoid overconfidence issues caused by clipping of observations in score computation, we evaluated direct MELD score estimation at 48 hours across six datasets with available ground truth labels (Figure S21 B). The model achieved a mean $R^2 = 0.41 \pm 0.30$ and $\text{MAE} = 4.9$ MELD points across datasets. Performance varied by center, with MIMIC ($R^2 = 0.73$) and HiRID ($R^2 = 0.64$) showing strong agreement, while INSPIRE ($R^2 < 0$) exhibited poor fit, potentially due to the differences in patient populations (surgical patients in an Asian dataset) or laboratory measurement practices. The regression fit closely follows the identity line for MELD scores below 40, with increasing deviation at higher scores where fewer training examples exist. These results demonstrate that the foundation model can provide clinically meaningful continuous risk estimates, though site-specific calibration may benefit centers with divergent patient characteristics.

#### C.10 Complementary adaptation results

**High precision deployments** In **Figure S22 A)** we focus on a high precision regime for a targeted deployment of the model with low false positive rate (low false alarm rate in a deployed early warning system) and observe that in these high precision regions ICareFM's performance compares well to locally trained reference models. The adapted models become inferior at $34{,}013$ local patients or even only beyond $50{,}000$ patient stays in the case of stage-adapted models (conditioned on the subset where intersections are observed within the extrapolation bounds of at most twice the available labeled patient stays).

**Data efficiency** We quantify ICareFM's data efficiency by measuring the factor by which fine-tuning the foundation model reduces the training data required to match a given performance level compared to training local models (**Figure S22 B)**). For each task-dataset combination, we sample performance levels within the range achievable by both the fine-tuned and reference models and compute the ratio of training samples each requires to reach that level. We restrict this comparison to the overlapping performance range to avoid extrapolation artifacts where the fine-tuned model exceeds the reference model's performance (observed $52\%$ and $72\%$ for local and staged adaptation respectively). At low sample sizes (100–300 stays), ICareFM provides a median efficiency gain of roughly $4\times$, meaning the reference model requires 4 times more data to reach the same AuROC. This advantage decreases with increasing local data availability, with both adaptation strategies converging toward the $100\%$ baseline beyond ten thousand patients. The staged adaptation strategy consistently shows higher efficiency than local adaptation across all sample size bins above the baseline, reflecting the additional benefit of external task-agnostic supervision until break even (note again that in many cases the available local data was not enough for the reference model to reach the same performance and was hence excluded from this analysis). These results indicate that ICareFM's adaptation yields relevant data efficiency gains for small cohorts and often allows outperforming local models even for large cohorts. Beyond predictive performance, understanding *how* ICareFM represents patient states can provide insight into what the model has learned and whether its internal organization reflects clinically meaningful structure.

#### C.11 PSSS Study: same-country cross-hospital transfer analysis

Foundation models derive their value from the ability to generalize across distribution shifts even without requiring local data for adaptation. However, in healthcare settings where regulatory and practical considerations may favor regional data sharing, a relevant question emerges: does training supervised models on same-country data from different hospitals provide superior sample efficiency compared to dual zero-shot transfer from a globally trained foundation model?

To investigate this question, we designed an experiment comparing ICareFM's transfer performance to Swiss hospitals against supervised baselines trained on data from other Swiss hospitals. Specifically, we trained reference models (LightGBM and GRU architectures as described in Supplementary C.1) using varying amounts of data sampled from Swiss hospitals excluding the target hospital, then evaluated performance on the held-out target Swiss hospital. This differs from our primary analysis where reference models were trained exclusively on in-distribution data from the target hospital itself.

In Figure S24 we compare ICareFM trained on data from US, Asia, Europe, and including HiRID data from Inselspital Bern and compare its performance to supervised models trained on increasing amounts of external data from other Swiss hospitals. Note that in the case of the Bern cohort in the PSSS study this represents only a temporal shift from 2016 (as the latest collection point in HiRID) to 2019 (start of the PSSS data collection in Bern). Compared to the results presented in Figure 2 we observe that ICareFM's estimated value increases when the comparator changes from a model trained on the target hospital to a model trained on the target country (but excluding the target hospital). Median zero-shot patient performance equivalence is estimated at $1,024$ patients, whereas the median zero-shot patient performance equivalence in Figure S24 comparing to same country external patients is $1,585$ representing an over $50\%$ improvement. The externally task-adapted trend (domain generalization) is even stronger going from median $2,959$ to $32,031$ but the estimate here is inflated because transfer performance of the same country supervised models plateaued and often the externally task-adapted ICareFM strictly transferred better.

This finding has important implications for deployment strategies: while regional data sharing initiatives may seem advantageous for building locally adapted models, globally trained foundation models can achieve comparable or superior performance without requiring local training infrastructure or cross-institutional data sharing agreements. The sample efficiency advantage of foundation models persists even when comparing against models trained on data from the same country, suggesting that the distributional knowledge captured during pretraining on diverse international ICU datasets generalizes effectively to new hospitals regardless of geographic proximity.

###### C.11.1 Comprehensive out-of-distribution transfer performance

To provide a broader view of cross-hospital transfer capabilities, we evaluated ICareFM against a diverse set of potential data sources for transfer to Swiss PSSS cohorts. Beyond same-country pooled data, we assessed transfer performance from individual hospitals across different continents (Asia, US, Europe, Switzerland), Swiss pooled PSSS data combining multiple Swiss centers but excluding the target, ICareFM zero-shot transfer with varying geographic training data compositions (US+Asia, US+Asia+Europe, US+Asia+Europe+Switzerland), and ICareFM domain generalization (externally task-adapted ICareFM) with the same geographic training compositions. Note that for the last stage when including Swiss data from HiRID in the pretraining corpus for the single PSSS target of Bern this no longer represents a domain shift but only a temporal shift with a time gap of 3 years (2016 to 2019) in between the HiRID and Bern PSSS cohorts.

The heatmap visualizations in Figure S23 present task-averaged performance metrics for all target-source combinations across four Swiss PSSS cohorts (Basel, Bern, Lausanne, Zurich). Each cell represents the mean performance across all prediction tasks available for that target cohort. The inclusion of median and minimum statistics across targets provides aggregate measures of transfer robustness.

Several patterns emerge from this evaluation. First, transfer performance from single-source hospitals varies substantially by geographic origin, with US hospitals (MIMIC-IV, eICU) generally providing stronger transfer than Asian sources (INSPIRE), suggesting that data characteristics and clinical protocols in US academic medical centers may share greater similarity with Swiss institutions (note that we did not control for training dataset size to avoid underestimating performance). Second, Swiss pooled data combining multiple PSSS centers achieves competitive performance (median AuROC: 0.83, median AuPRC: 0.41), demonstrating that regional data sharing can be effective when feasible. However, ICareFM's external adaptation transfer matches or exceeds this performance (median AuROC for US+Asia+Europe configuration: 0.83, median AuPRC: 0.43), achieving similar discrimination and superior precision without requiring cross-institutional data sharing agreements or local model training.

Third, the progressive inclusion of geographically diverse training data in ICareFM consistently improves transfer performance even if the additional training data from European institutions does not substantially grow the pretraining dataset size. Zero-shot models trained on US+Asia alone achieve median AuROC of 0.78 and AuPRC of 0.35, while adding European data increases these to 0.79 and 0.37, and further including Swiss historical data (HiRID, pre-2016 Bern) reaches 0.79 and 0.36 for zero-shot, with external adaptation achieving

0.83 and 0.44 respectively. This gradient suggests that exposure to diverse ICU environments during pretraining enhances the model's ability to capture generalizable clinical patterns beyond region-specific practices.

The minimum performance statistics reveal robustness patterns: ICareFM external adaptation maintains minimum AuROC of 0.80 and AuPRC of 0.38 across all Swiss targets even without training on Swiss data, while single-source transfers can degrade to AuROC of 0.73 and AuPRC of 0.21 (INSPIRE to Basel). This worst-case stability matters for clinical deployment, where consistent performance across diverse patient populations and institutional contexts is essential.

These findings collectively support the hypothesis that foundation models trained on globally diverse ICU data can achieve transfer performance competitive with or superior to regional data pooling approaches. While same-country data sharing may seem intuitively advantageous due to shared clinical protocols and patient demographics, the distributional knowledge captured by large-scale pretraining on heterogeneous international datasets appears to provide comparable or greater generalization capabilities. This has practical implications for healthcare systems where cross-institutional data sharing faces regulatory, technical, or organizational barriers: deploying globally pretrained foundation models may be a more feasible path to achieving predictive performance than establishing regional data sharing infrastructure.

These results support ICareFM as a practical default option, with effective adaptability given even small samples of local data (Figure 2).

#### C.12 Latent space analysis

To characterize the structure of ICareFM's learned representations and their relationship to clinical trajectories and outcomes, we performed latent space analysis using dimensionality reduction, trajectory flow field estimation, and outcome density mapping. This analysis reveals how the foundation model organizes clinical states and temporal dynamics within its internal representation space.

##### C.12.1 Dimensionality reduction

We performed stratified sampling of patient trajectories from datasets for visualization and analysis. For general trajectory analysis and visualization, we sampled 3,000 patients per dataset with up to 168 hourly (1 week) observations per patient starting from admission.

We found that already linear PCA successfully extracted clinically relevant structure over dataset-specific artifacts. Despite PCA's theoretical limitations for capturing non-linear manifold structure, we highlight strong separation of core clinical trajectories suggesting that the foundation model's pretraining induced approximately linear separability of key clinical patterns.

We obtain ICareFM representations using a model trained on all datasets jointly to obtain a single shared latent representation space. We fit PCA with 2 components on validation set embeddings and applied the learned projection to test set embeddings to avoid overfitting visualization choices to the analyzed data. The first two principal components captured substantial variance in the embedding space, with cumulative explained variance reported in all visualizations. All subsequent trajectory and density analyses operated on these PCA-projected coordinates.

##### C.12.2 Trajectory flow field estimation

**Direction Vector Computation.** For each timepoint in the embedding space, we estimated a local trajectory direction by examining temporal neighborhoods within each patient stay. We extract two trajectory estimates:

- **Past-mode direction** (default): Computed from the most extreme past point (within $n_{\text{prev}}$ timesteps) to the current point, indicating "where the trajectory came from." The extreme point was selected as the past observation (within 2 hours) with minimum projection along the estimated trajectory direction vector.
- **Future-mode direction**: Computed from the current point to the most extreme future point (within $n_{\text{prev}}$ timesteps ahead), indicating "where the trajectory is going." The extreme point was selected as the future observation (within 2 hours) with maximum projection along the trajectory direction vector.

The extreme-point aggregation strategy was chosen over mean-based aggregation to emphasize trajectory momentum and avoid dampening directional signals from noise or physiological oscillations. Direction vectors were represented as angles in radians $\theta \in [-\pi, \pi]$ via $\theta = \arctan 2(\Delta y, \Delta x)$. Velocity magnitude (Euclidean distance) was computed as $v = \sqrt{(\Delta x)^2 + (\Delta y)^2}$.

**Grid-Based Spatial Aggregation.** To create interpretable flow field visualizations, we discretized the 2D embedding space into regular grids (typically $10 \times 10$ to $15 \times 15$ cells with 0.5–1.0 unit padding beyond data extent). For each grid cell with sufficient observations (minimum 10 timepoints), we computed:

1. **Circular mean direction**: Aggregated angles using circular statistics to properly handle $\pm\pi$ wraparound:
$$\bar{\theta} = \arctan 2\left(\frac{1}{n}\sum_{i=1}^{n}\sin(\theta_i), \frac{1}{n}\sum_{i=1}^{n}\cos(\theta_i)\right) \tag{17}$$
2. **Mean resultant length** $R \in [0, 1]$: Measures directional consistency, where $R = 1$ indicates perfect alignment and $R = 0$ indicates uniform angular distribution:
$$R = \sqrt{\left(\frac{1}{n}\sum_{i=1}^{n}\cos(\theta_i)\right)^2 + \left(\frac{1}{n}\sum_{i=1}^{n}\sin(\theta_i)\right)^2} \tag{18}$$
3. **Mean velocity**: Average Euclidean velocity magnitude across timepoints in the cell.
4. **Cell centroid**: Mean position of all timepoints assigned to the cell, used for arrow placement in visualizations.

**Arrow Visualization.** Grid-aggregated direction vectors were visualized as arrows with length proportional to normalized mean velocity (scaled to 0.5–1.0 of base arrow length to maintain visibility) and transparency encoding data density within the cell. Arrows were positioned at cell centroids and colored to distinguish past-trend and future-trend directions.

##### C.12.3 Outcome density estimation

**Kernel density estimation.** To visualize spatial clustering of clinical outcomes in the latent space, we estimated probability density functions for specific events (circulatory failure, respiratory failure, kidney failure, mortality) using Gaussian kernel density estimation. For computational efficiency, we subsampled to 100,000 timepoints when necessary before KDE fitting.

Density was estimated on a $100 \times 100$ regular grid spanning the embedding space extent. We used Scott's rule for automatic bandwidth selection, which typically yielded effective smoothing of $\sigma \approx 1$ PCA unit.

**Stratified Trajectory Analysis.** To isolate outcome-specific trajectory patterns, we computed separate flow fields conditioned on clinical states:

- **Event-free patients / stable**: Patients with no organ failure events (circulatory, respiratory, kidney, liver) or mortality throughout their stay
- **Unstable patients**: Patients experiencing at least one organ failure event or death
- **Outcome-specific trajectories**: Flow fields computed using only timepoints preceding specific events (e.g., circulatory failure within 8 hours)

This stratification revealed distinct trajectory dynamics: event-free patients exhibited convergent flows toward stable attractor regions, while unstable patients showed divergent or transitional flows through outcome-enriched zones.

##### C.12.4 Individual patient trajectory visualization

For an example patient (Figure 4 C) we visualized the individual trajectory by:

1. **Temporal smoothing**: Applying 1D Gaussian filtering along the time axis to raw PCA coordinates to reduce high-frequency noise while preserving trajectory structure
2. **Superposition**: Overlaying smoothed trajectory lines and raw observation points on outcome density maps
3. **Clinical correlation**: Synchronizing trajectory plots with time-series plots of relevant physiological variables and annotated outcome events

Smoothing was applied post-hoc for visualization only; all density and flow field analyses used unsmoothed embeddings.

#### C.13 Fairness analysis

**Metrics** For each dataset–task pair, we stratify predictions by demographic group and compute group-level performance metrics for every group with at least 30 members. We report standard utility metrics (AuROC, AuPRC) alongside two fairness disparity metrics[88;89]:

- **ECE disparity**. Expected calibration error measures how well predicted probabilities match observed event frequencies. ECE disparity is the maximum pairwise difference across demographic groups:

$$\Delta\text{ECE} = \max_g \text{ECE}_g - \min_g \text{ECE}_g.$$

  Higher values indicate that the model is better calibrated for some groups than others. We compute ECE scores for local reference models, as well as variants of ICareFM. We exclude clinical scores as many considered in this work provide only a small discrete set of possible scores and have not been designed for the variety in cohorts observed here. Local reference models are generally well calibrated for the overall local cohort and provide a strong reference point.

- **Equalized odds disparity**. Equalized odds[90;91] requires that a classifier's true positive rate (TPR) and false positive rate (FPR) are equal across groups at a given operating point. We define equalized odds disparity as:

$$\Delta\text{EOD}(t) = \max\Big(\max_g \text{TPR}_g(t) - \min_g \text{TPR}_g(t),\ \ \max_g \text{FPR}_g(t) - \min_g \text{FPR}_g(t)\Big),$$

  where $t$ is the decision threshold. We evaluate this at the 1% FPR operating point (low false alarm rate) used throughout this work (see Supplementary C.11). At this operating point, the FPR disparity term is small by construction, hence the metric is primarily driven by TPR differences across groups.

**Approach** Our evaluation spans 4 models, 9 datasets, 7 tasks, and 3 demographic properties, yielding over 100 model–dataset–task–variable combinations. Drawing conclusions from per-group results at this scale is infeasible, so inspired by the fairness benchmarking literature[88;92], we compare models by examining the distribution of disparity metrics across all dataset–task pairs (Supplementary C.13 and Figure S26 A, B). We show examples of group-level performance for eICU and circulatory failure in Figure S26 C. Complete per-group performance breakdowns for all models, datasets, tasks, and demographic properties are provided in Tables S1–S2 (available as separate files). We note that aggregated disparity metrics inevitably compress information relative to these full breakdowns, and that such compression can obscure important patterns[93]. This highlights the importance of future work on in-depth fairness analysis for specific models and cohorts.

**Statistical comparison** To formally compare LGBM (locally trained reference) against ICareFM (external adaptation), we use the Bayesian signed-rank test[94], which estimates three posterior probabilities for each comparison: $P(\text{LGBM better})$, $P(\text{practically equivalent})$, and $P(\text{ICareFM better})$. Two models are considered practically equivalent when their disparity metric difference falls within a region of practical equivalence (ROPE) of $[-\delta, +\delta]$. This test offers advantages over classical null hypothesis significance testing by enabling direct estimation of the probability that two models are practically equivalent, rather than testing only against a point null of zero difference[94;95]. We interpret posterior probabilities using the following decision rules, adapted from the thresholds used in Benavoli et al.[94] and Maron et al.[95]. All rules use a single posterior probability threshold of 0.9: *equivalence* when $P(\text{equiv}) > 0.9$; *non-inferiority* of model X when $P(\text{equiv}) + P(\text{X}) > 0.9$ but $P(\text{equiv}) \leq 0.9$; *model X better* when $P(\text{X}) > 0.9$; and *inconclusive* otherwise.

We set $\delta = 0.05$ as the primary margin for all metrics. To our knowledge, no published standard defines thresholds for clinically meaningful calibration or equalized odds disparity[96;97]. The absence of such standards has also been noted in fairness benchmarking work[88;98]. We chose $\delta = 0.05$ guided by two considerations. First, disparities of this order are consistent with the range of ECE and equalized odds gaps reported across foundation models and datasets in recent fairness benchmarks[88]. Second, this value is comparable to the magnitude of metric fluctuations we would expect when retraining models with different random seeds or bootstrap resamples of patients, and below which differences are attributable to stochastic variation rather than systematic model differences. This choice remains a modelling decision rather than a clinically-verified standard, so we report sensitivity analysis at $\delta = 0.02$ (strict) and $\delta = 0.08$ (lenient). We encourage future work to establish validated, task-specific thresholds for fairness disparities relevant for practical clinical deployment.

| Metric Disparity | Variable | $\delta = 0.02$ | | | | $\delta = 0.05$ | | | | $\delta = 0.08$ | | | |
|---|---|---|---|---|---|---|---|---|---|---|---|---|---|
| | | P(LGBM) | P(Equiv) | P(ICareFM) | Conclusion | P(LGBM) | P(Equiv) | P(ICareFM) | Conclusion | P(LGBM) | P(Equiv) | P(ICareFM) | Conclusion |
| ECE Disparity | Sex | 0.000 | 1.000 | 0.000 | Equivalence | 0.000 | 1.000 | 0.000 | Equivalence | 0.000 | 1.000 | 0.000 | Equivalence |
| | Age | 0.000 | 1.000 | 0.000 | Equivalence | 0.000 | 1.000 | 0.000 | Equivalence | 0.000 | 1.000 | 0.000 | Equivalence |
| | Ethnic | 0.002 | 0.998 | 0.000 | Equivalence | 0.000 | 1.000 | 0.000 | Equivalence | 0.000 | 1.000 | 0.000 | Equivalence |
| EOD @ 1% FPR | Sex | 0.000 | 1.000 | 0.000 | Equivalence | 0.000 | 1.000 | 0.000 | Equivalence | 0.000 | 1.000 | 0.000 | Equivalence |
| | Age | 0.869 | 0.006 | 0.126 | Inconclusive | 0.083 | 0.917 | 0.000 | Equivalence | 0.006 | 0.994 | 0.000 | Equivalence |
| | Ethnic | 0.506 | 0.438 | 0.056 | Inconclusive | 0.033 | 0.966 | 0.000 | Equivalence | 0.001 | 0.999 | 0.000 | Equivalence |
| EOD @ 10% FPR | Sex | 0.002 | 0.996 | 0.002 | Equivalence | 0.000 | 1.000 | 0.000 | Equivalence | 0.000 | 1.000 | 0.000 | Equivalence |
| | Age | 0.509 | 0.003 | 0.487 | Inconclusive | 0.069 | 0.907 | 0.025 | Equivalence | 0.002 | 0.998 | 0.000 | Equivalence |
| | Ethnic | 0.003 | 0.032 | 0.964 | ICareFM better | 0.000 | 0.889 | 0.111 | ICareFM non-inferior | 0.000 | 0.994 | 0.006 | Equivalence |

**Table S4:** Statistical comparison of fairness metrics for local LGBM and domain-generalized ICareFM using Bayesian ROPE two-sided tests. $P$ indicates the posterior probability of a model being better or the two models being equivalent up to the difference of $\delta$. Conclusions follow prespecified decision rules: *equivalence* when $P(\text{Equiv}) > 0.9$; *non-inferiority* of model X when $P(\text{Equiv}) + P(\text{X}) > 0.9$ but $P(\text{Equiv}) \leq 0.9$; *X better* when $P(\text{X}) > 0.9$; and *inconclusive* otherwise[94;95]. At the primary margin of $\delta = 0.05$, the two models are equivalent across all metric-variable combinations except EOD at 10 % FPR on ethnicity, where ICareFM is non-inferior.

**Results** Posterior probabilities are shown in Table S4. At the primary threshold $\delta = 0.05$, the two models are equivalent across all metric-variable combinations. Equivalence is clear for all ECE disparity comparisons, for EOD on sex at both operating points, and for EOD at 1 % FPR on ethnicity. For EOD on age at both operating points, equivalence holds with $P(\text{equiv})$ of 0.92 and 0.91, respectively. For EOD at 10 % FPR on ethnicity, $P(\text{equiv}) = 0.89$ falls just below the equivalence threshold. However, the one-sided probability $P(\text{ICareFM not worse}) = 1.00$ provides strong evidence of ICareFM non-inferiority. At the lenient margin ($\delta = 0.08$), equivalence is strong across the board.

For ECE disparity, equivalence holds even at the strictest threshold tested of $\delta = 0.02$, reflecting the very small absolute calibration differences visible in Figure S26 A. For EOD, the picture is more nuanced. At $\delta = 0.02$ for the 1 % FPR, there is moderate evidence that LGBM achieves lower TPR disparity for age, and results are inconclusive for ethnicity. Conversely, at the 10 % FPR operating point and the same strict margin, ICareFM achieves lower EOD for ethnicity. These contrasting findings at strict margins illustrate that the direction of small disparity differences depends on the EOD operating threshold, which is consistent with the general observation that fairness properties are threshold-dependent. Importantly, at the primary margin all of these differences are absorbed within the ROPE. We note that classification thresholds are defined globally in this work. Therefore adjusting them per subgroup could improve fairness properties. We encourage future work to investigate this choice as well as the utility-fairness tradeoff.

These results indicate that at the primary $\delta$, ICareFM achieves fairness properties comparable to a locally trained reference model despite being trained exclusively on external data. The sensitivity analysis further reveals that at finer margins, fairness properties vary across demographic axes and operating thresholds. This highlights that the specific metric, classification threshold, and choice of $\delta$ are important considerations for practical fairness and should be studied in future works in order to derive standards for models for deployment.

#### C.14 External Validation: Charité – Universitätsmedizin Berlin

We perform additional validation of the generalization bounds and performance estimates established on the core data sources harmonized for the study. Charité data was only integrated and harmonized after model design and training had been completed. No hyperparameter or architectural design choices could or have been influenced by Charité data.

**External validation of the LPE bounds** We train the same set of reference models (Supplementary C.1) on the Charité data as for the other included datasets and create scaling curves for reference models, adapted models, and run inference for externally trained models. Note however, that due to the large size of the cohort, we can establish a data-confirmed scaling curve up to 250,000 local patient stays.

We then perform direct inference of ICareFM in zero-shot mode and in externally task adapted mode and observe and report the intersections with the locally trained models. Similarly we tune using local Charité data in the local and staged adaptation modes to assess LPE (Supplementary C.4) intersections with the local model's scaling behavior. Results are shown in Figure 3 D/E).

**Assessing the impact of pretraining cohort heterogeneity** We use the Charité cohort's large size to answer the question whether a foundation model trained on a single more uniform cohort can compete with the same

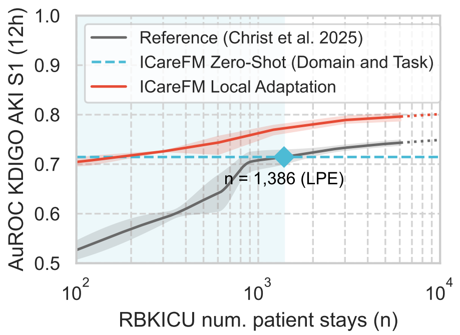


**Figure 6: KDIGO AKI Stage 1 Comparison** of ICareFM with a local task-specific model developed by Christ et al. at RBK. We compare the local task-specific survival model on a clinically relevant 12 hour prediction horizon to ICareFM in zero-shot and local adaptation mode.

model trained on a heterogeneous cohort from different smaller hospitals.

We train ICareFM using the full training set of the Charité cohort and then train a variant of ICareFM using an equal amount of training and validation data sampled at random from the heterogeneous collection of harmonized ICU datasets (holding out again a target dataset from training completely). We then compare transfer performance of the Charité based ICareFM with the heterogeneous ICareFM model in zero-shot and external adaptation mode (Section 2.2, Supplementary B.9). Results are shown in Figure 3 C).

**Validating scaling curve behavior** We train a variant of ICareFM including the training split of the Charité cohort in the pretraining set and then assess zero-shot and external adaptation transfer performance to nine external ICU databases held-out from training. The observed performances contribute as additional validation points to the scaling behaviors established in Figure 3 A and B as well as in Figure S12 C. Note that the additional points have not been used to derive the reported scaling law, but rather to validate and contextualize it.

**Temporal generalization and ICU ward type sensitivity** We assess the temporal generalization behavior of ICareFM on the Charité cohort over time. Given the pretraining cohort Tables S1 and S2 ICareFM's *knowledge cutoff* is in 2022 (EHRSHOT was not used for pretraining). We bin patient stays by year and assess yearly performance of ICareFM in zero-shot and external adaptation mode (hence not using any local Charité data for adaptation). Results are shown in Figure 3 F). Similarly we stratify the cohort by ICU ward type to observe ICareFM's performance sensitivity to specialized ICU units. Due to the long evaluation period several years back into the past not all stays could be mapped reliably to a the specialization of a specific ward at a current time in the past (since these also changed over time). Only patient stays with a reliable assignment were considered for this stratification. Results are shown in Figure 3 G) (external adaptation) and in Figure S27 (dual zero-shot).

#### C.15 External Validation: Robert Bosch Krankenhaus

We perform additional validation of the generalization bounds and performance estimates established on the core data sources harmonized for the study. Data from the Robert Bosch Krankenhaus (RBKICU) data was only integrated and harmonized after model design and training had been completed. No hyperparameter or architectural design choices could or have been influenced by RBKICU data.

**External validation of the LPE bounds** We train the same set of reference models (Supplementary C.1) on the RBKICU data as for the other included datasets and create curves for reference models, adapted models, and run inference for externally trained models. Results are shown in Figure 3 D/E).

##### C.15.1 Comparison with locally developed specialized kidney model

We perform a dedicated comparison to a locally developed specialized model by Christ et al.[99] at RBK to detect KDIGO Stage 1 AKI (Acute Kidney Injury) on a heart surgical cohort. The cohort is a subset of the full RBKICU dataset harmonized and benchmarked for the validation experiments of ICareFM.

We perform inference on the model by Christ et al. at varying sample sizes and compare to ICareFM in zero-shot mode and in local adaptation mode also being adapted using growing sample sizes.

Note that while the labels have been matched exactly the two models have been shown different data representations. ICareFM was shown the general purpose selection of harmonized clinical variables determined as relevant to build a general patient state representation, while the local model uses an optimized kidney failure oriented dataset. While some information overlaps, ICareFM's input contains a richer set of treatments and more variables not directly related to kidney function, while the local model had access to specialized data such as pre-ICU creatinine levels or surgical urgency status. Inference was provided by the authors of the local model.

Results on a fixed 12 hour prediction horizon are shown in Figure 6. ICareFM zero-shot performs similarly to the local model using about 1,300 training patients, which is in line with the general generalization bound estimates provided in the result section Section 3. ICareFM notably outperforms the reference model by Christ et al. when using local data for adaptation.

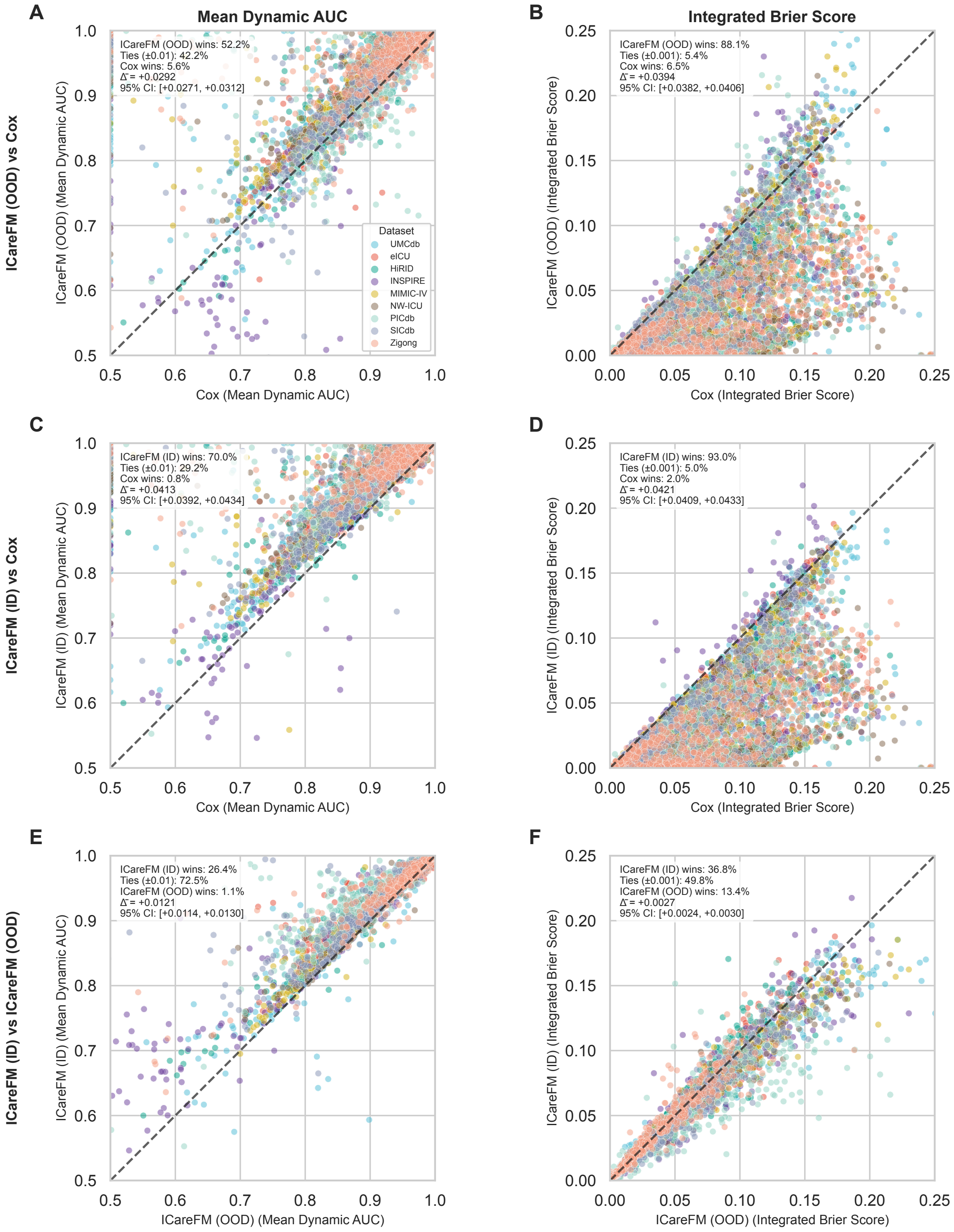


**Figure S15: Pretrained ICareFM Survival Evaluation** Paired scatter plots comparing ICareFM model performance against Cox baseline and between ICareFM variants. **(A–B)** ICareFM zero-shot vs Cox; **(C–D)** ICareFM adapted vs Cox; **(E–F)** ICareFM adapted vs zero-shot. The *adapted* in-distribution (ID) model continued the *pretraining* using in-distribution data after finishing the large pretraining on external data. Left column: Mean Dynamic AUC; right column: Integrated Brier Score. Each point represents one comparison across dataset-variable-threshold-scenario combinations (n=6,487). Points above (AUC) or below (IBS) the diagonal indicate superior performance of the y-axis model. Colors denote datasets (legend in A). Summary statistics show win percentages, ties, and mean difference (Δ). ICareFM models outperform Cox, with adapted models showing additional gains over zero-shot.

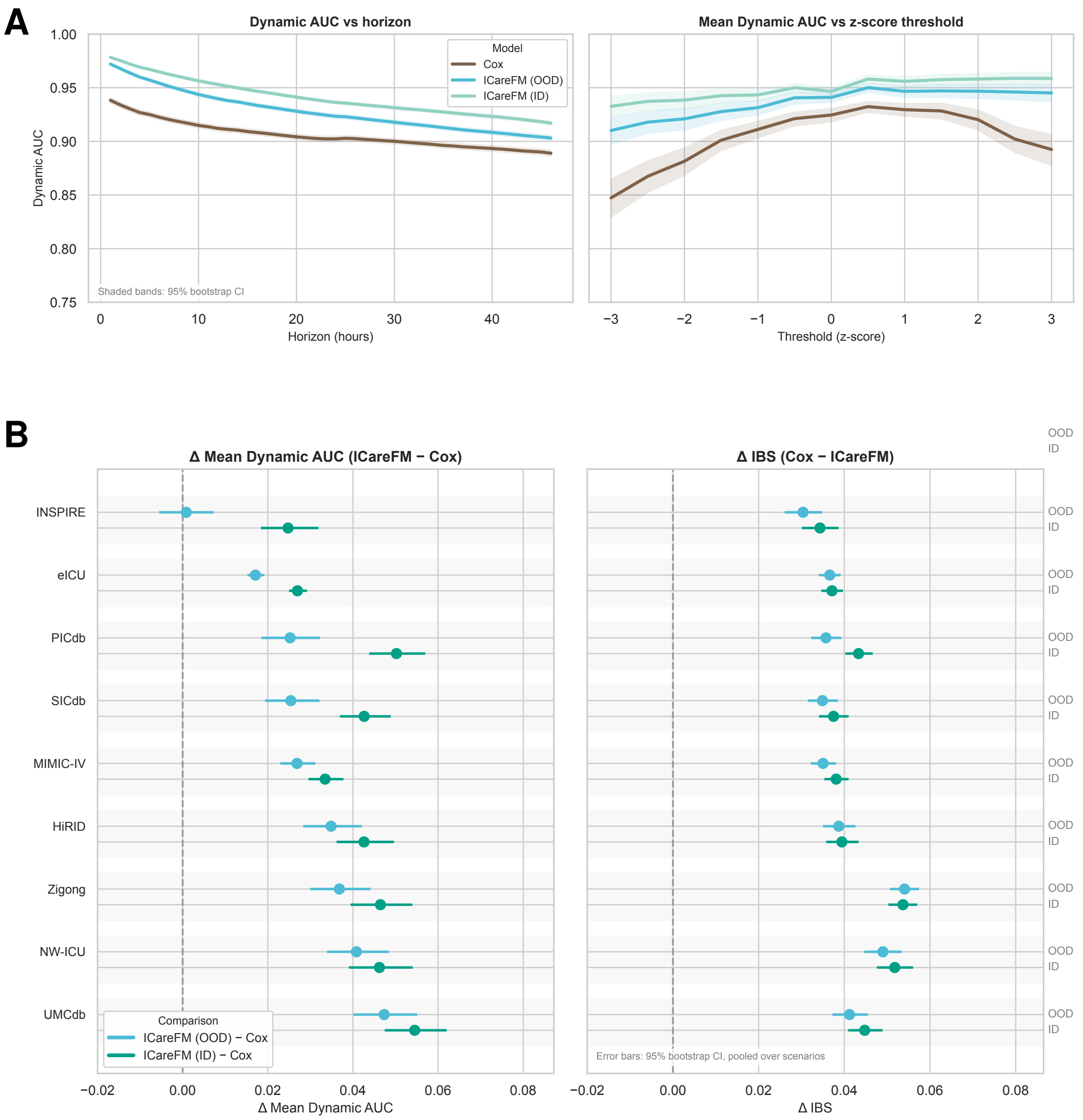


**Figure S16: Pretrained ICareFM Survival Evaluation A)** ICareFM performance across prediction horizons and event thresholds. Dynamic AUC of ICareFM models and the Cox baseline is summarized across all variables, datasets, thresholds, and scenarios. **Left:** Dynamic AUC as a function of prediction horizon, showing that both ICareFM variants (zero-shot and adapted) consistently outperform Cox across clinically relevant horizons. **Right:** Mean Dynamic AUC as a function of the z-score event threshold, indicating that ICareFM maintains its advantage over Cox across a wide range of event severities. Shaded bands denote 95% bootstrap CI across variable–dataset–scenario combinations, and solid lines show the mean. **B)** ICareFM performance improvements by dataset. Forest plots comparing ICareFM models (zero-shot and adapted) against Cox baseline across nine ICU datasets. **Left:** Δ Mean Dynamic AUC (ICareFM - Cox); **Right:** Δ Integrated Brier Score (Cox - ICareFM). Each dataset shows two comparisons: zero-shot (light blue) and adapted (dark green) versus Cox. Error bars show 95% CI pooled across variable-threshold-scenario combinations. The "Overall" row (diamonds) summarizes across all datasets. ICareFM consistently outperforms Cox, with adapted models showing additional gains.

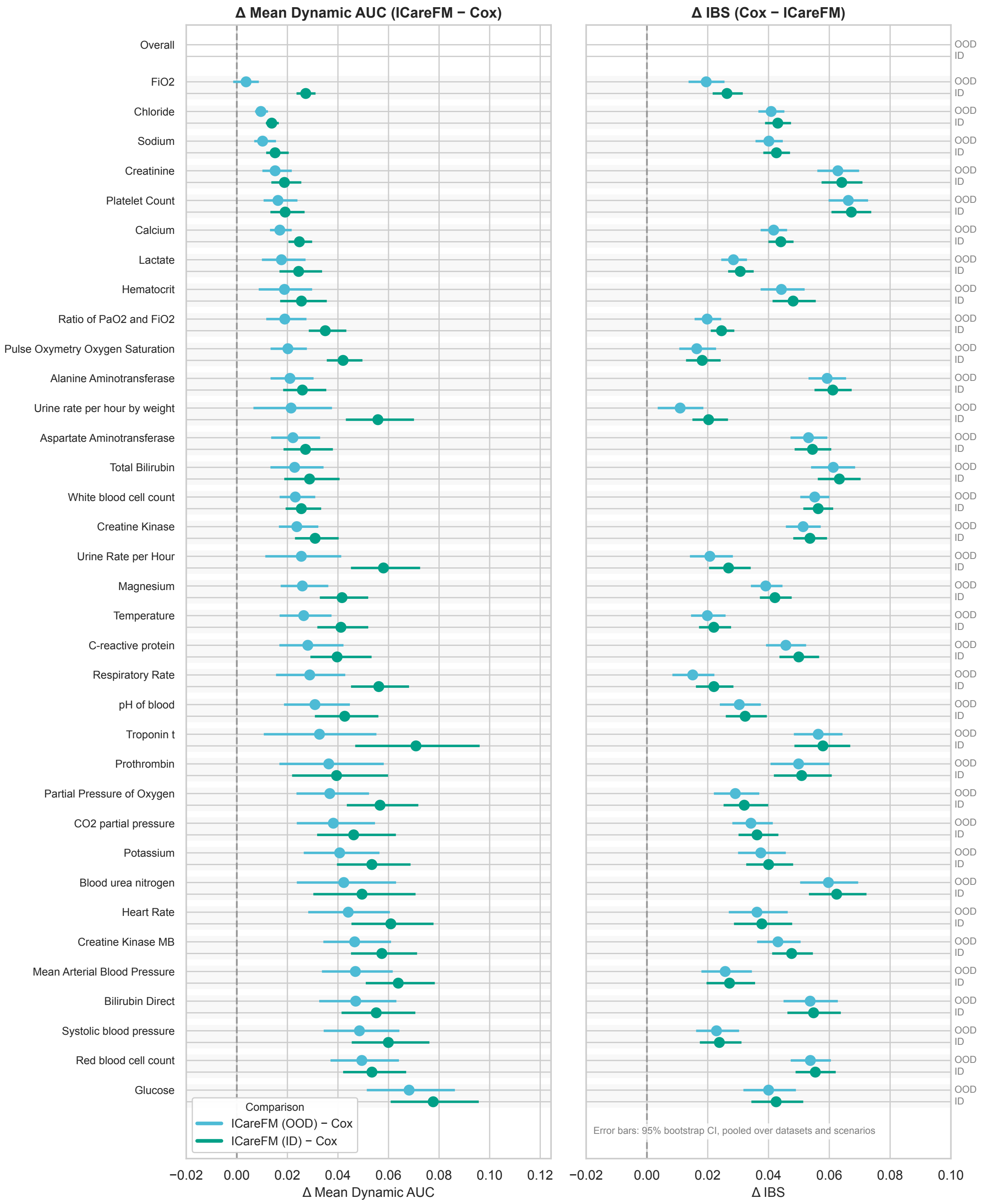


**Figure S17: ICareFM performance improvements by clinical variable, sorted by zero-shot benefit.** Forest plots comparing ICareFM models (zero-shot and adapted) against Cox baseline across 35 clinical variables. **Left:** Δ Mean Dynamic AUC (ICareFM - Cox); **Right:** Δ Integrated Brier Score (Cox - ICareFM). Each variable shows two comparisons: zero-shot (light blue) and adapted (dark green) versus Cox. Variables are ordered by zero-shot performance improvement (lowest to highest). Error bars show 95% CI pooled across dataset-threshold-scenario combinations. The "Overall" row (diamonds) summarizes across all variables. ICareFM consistently outperforms Cox across variables, with adapted models showing additional gains.

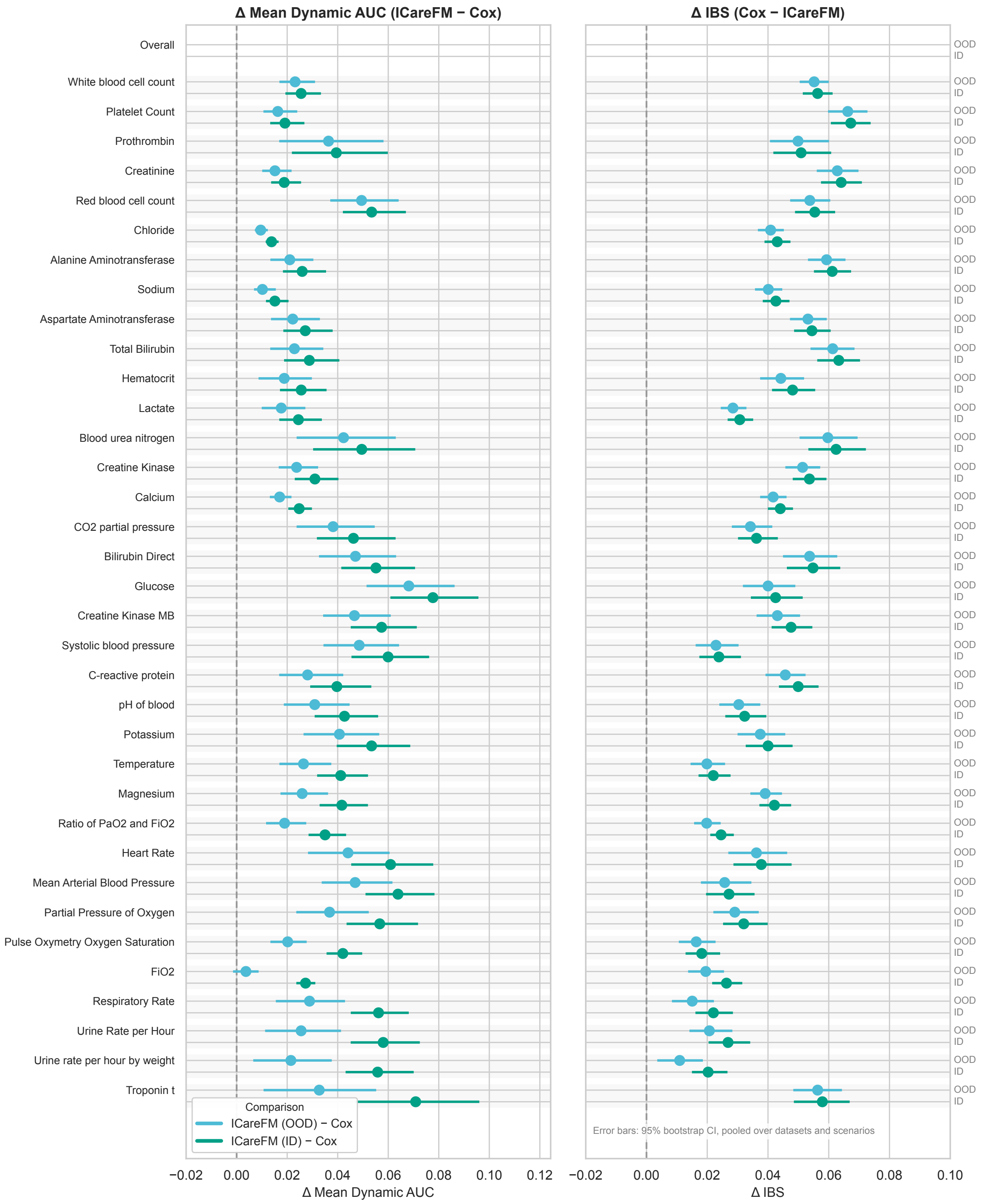


**Figure S18: ICareFM performance improvements by clinical variable, sorted by adaptation benefit.** Forest plots comparing ICareFM models (zero-shot and adapted) against Cox baseline across 35 clinical variables. **Left:** Δ Mean Dynamic AUC (ICareFM - Cox); **Right:** Δ Integrated Brier Score (Cox - ICareFM). Each variable shows two comparisons: zero-shot (light blue) and adapted (dark green) versus Cox. Variables are ordered by the additional benefit from adaptation ($\Delta_{\text{ID}} - \Delta_{\text{OOD}}$), from smallest to largest gain. Error bars show 95% CI pooled across dataset-threshold-scenario combinations. The "Overall" row (diamonds) summarizes across all variables. ICareFM consistently outperforms Cox across variables, with adapted models showing variable-specific adaptation benefits.

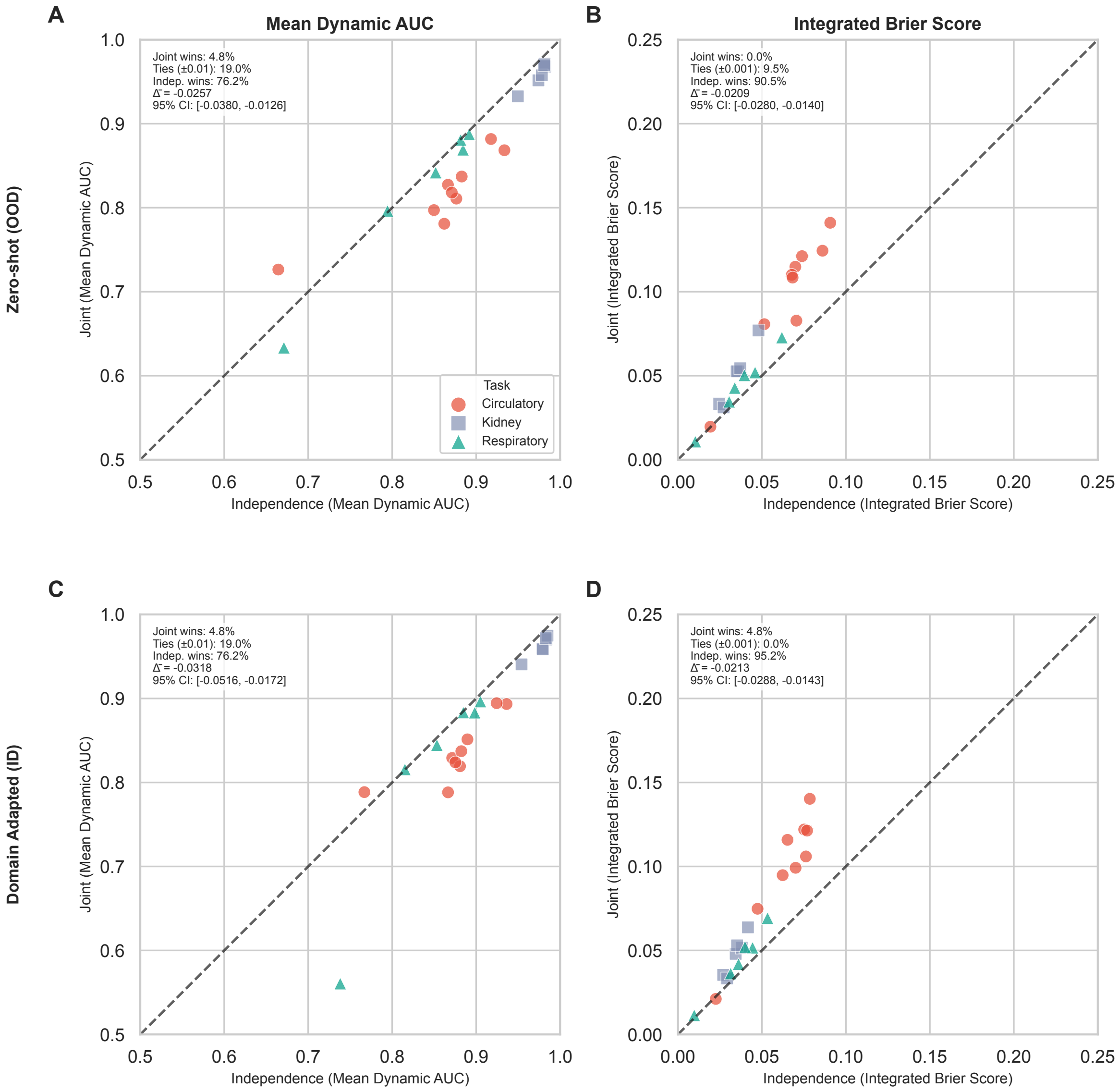


**Figure S19: Comparing bivariate versus univariate product of marginals inference** We compare the bivariate survival heads (Supplementary B.3.1) (*Joint*) performance against the product of marginals inference approach using the standard univariate heads (*Independence*). For circulatory failure we consider mean arterial pressure below 65 mmHg and blood serum lactate above 2 mmol/l, for respiratory we consider SpO2 below 90% and FiO2 above 60%, for kidney we consider creatinine > 2.0 mg/dl and Urine Rate < 0.5 ml/h. We measure mean time-dependent AuROC and integrated Brier Score. We note that the univariate product of marginals approach performs better in almost all tested settings.

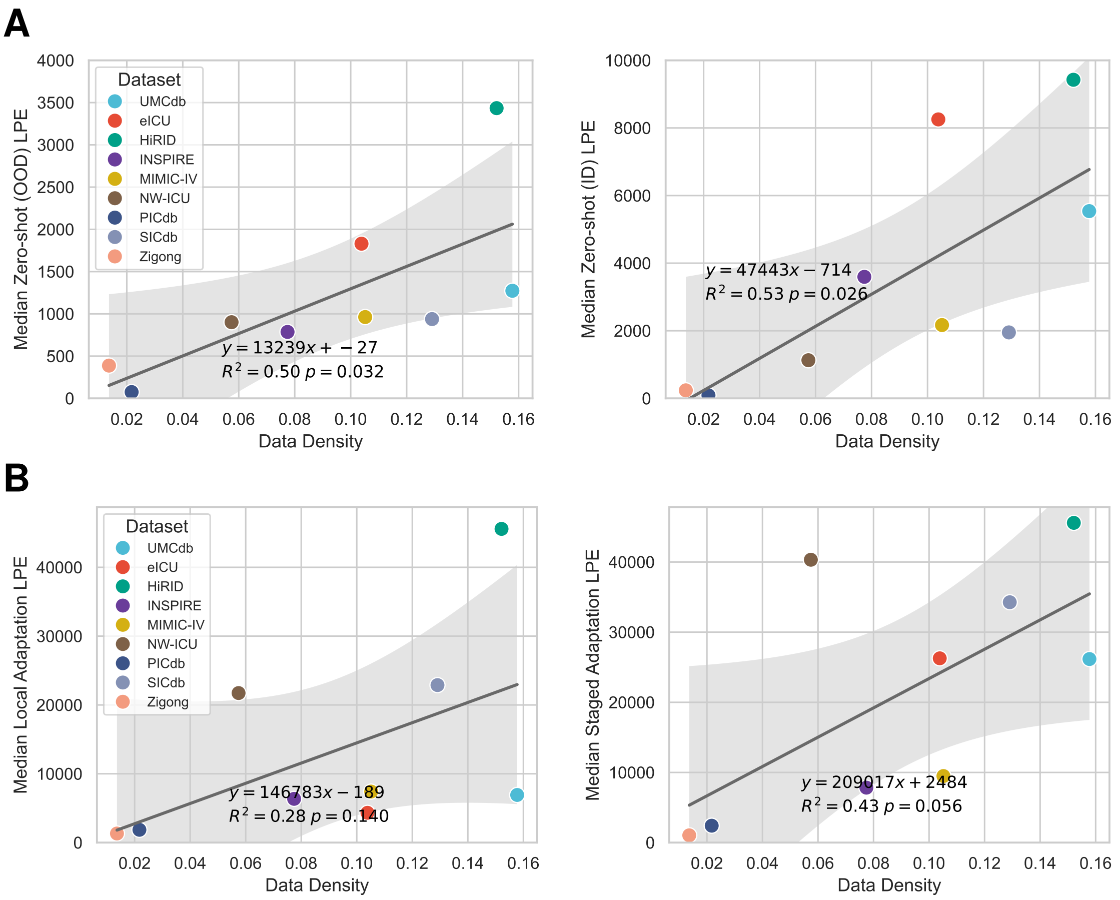


**Figure S20: Impact of data density on foundation model zero-shot performance.** Each point represents the median zero-shot patient equivalence across all benchmarked clinical tasks for a single dataset. **(A Left)** Out-of-distribution (OOD) evaluation: Median dual zero-shot LPE when the test dataset was excluded from pretraining, plotted against data density (proportion of non-missing values). Linear regression reveals a moderate positive correlation ($R^2 = 0.5$), with denser datasets yielding superior zero-shot generalization. **(A Right)** In-distribution (ID) evaluation (task zero-shot): Median zero-shot LPE when training data from the target hospital was used to continue pretraining. The slightly stronger correlation ($R^2 = 0.53$) indicates that data density has a more pronounced effect on in-distribution performance than on out-of-distribution generalization. Gray shaded regions denote 95% confidence intervals for the regression lines. Patient equivalence quantifies the number of supervised training patients required by a task-specific baseline to match the foundation model's zero-shot performance. **A Left** Local adaptation LPE against data density. **A Right** Staged adaptation LPE against data density.

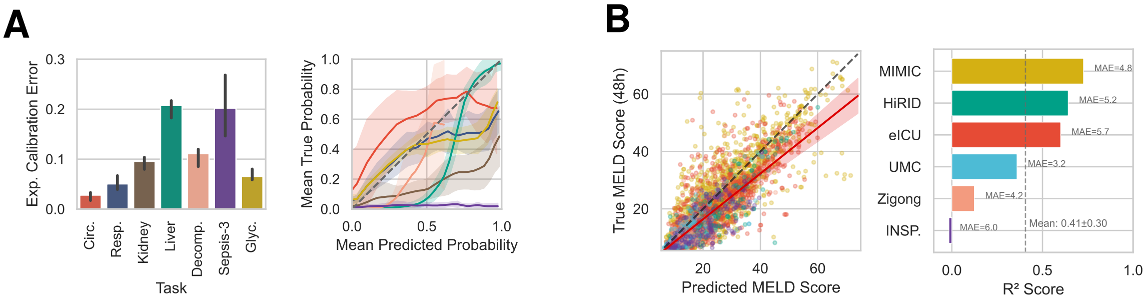


**Figure S21: Out-of-distribution calibration for early event predictions A) Zero-shot calibration analysis across clinical prediction tasks.** (Left) Expected Calibration Error (ECE) per task, aggregated across datasets (median with 50% prediction interval). (Right) Reliability diagrams showing mean predicted vs. true probability for each task; shaded regions indicate $\pm 1$ standard deviation across datasets. Dashed line indicates perfect calibration. Tasks: circulatory failure (Circ.), respiratory failure (Resp.), kidney failure (Kidney), liver dysfunction (Liver), decompensation (Decomp.), sepsis (Sepsis-3), and hyperglycemia (Glyc.). **B) MELD score regression analysis.** (Left) Predicted vs. true MELD scores at 48h across six datasets. Red line shows mean regression fit ($\pm 1\sigma$) across datasets; dashed line indicates perfect prediction. (Right) Per-dataset $R^2$ scores with mean absolute error (MAE) annotations. Mean $R^2 = 0.41 \pm 0.30$, MAE $= 4.9$ MELD points.

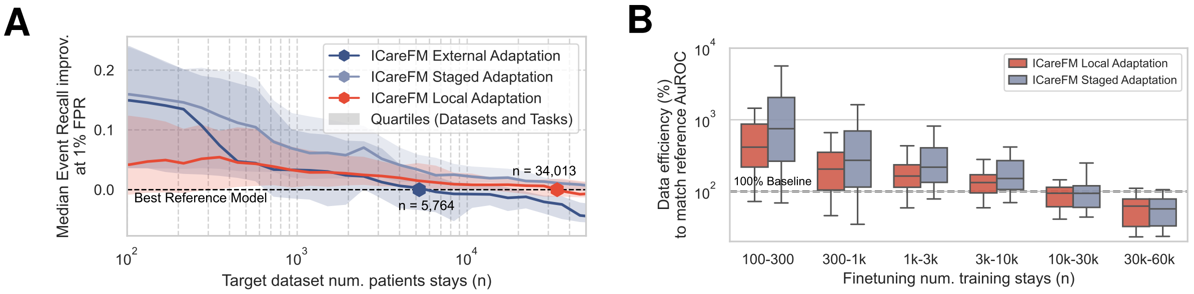


**Figure S22: Adaptation Capabilities A)** Event recall scaling behavior relative to reference models in high precision configuration for different adaptation settings. Note that reported intersections are conditioned on being observed within at most two times the available data (extrapolation bound), hence the reported intersection is only observed for a subset of the available data. **B)** Data efficiency trend with increasing local data size within the regions where performances have been observed for both ICareFM and the reference model without extrapolation. ICareFM exceeds the maximum performance achievable by the reference model in 56% and 66% for local and staged adaptation respectively (if additionally considering an equivalence margin of 0.5% AuROC this improves to 77% and 84%, respectively).

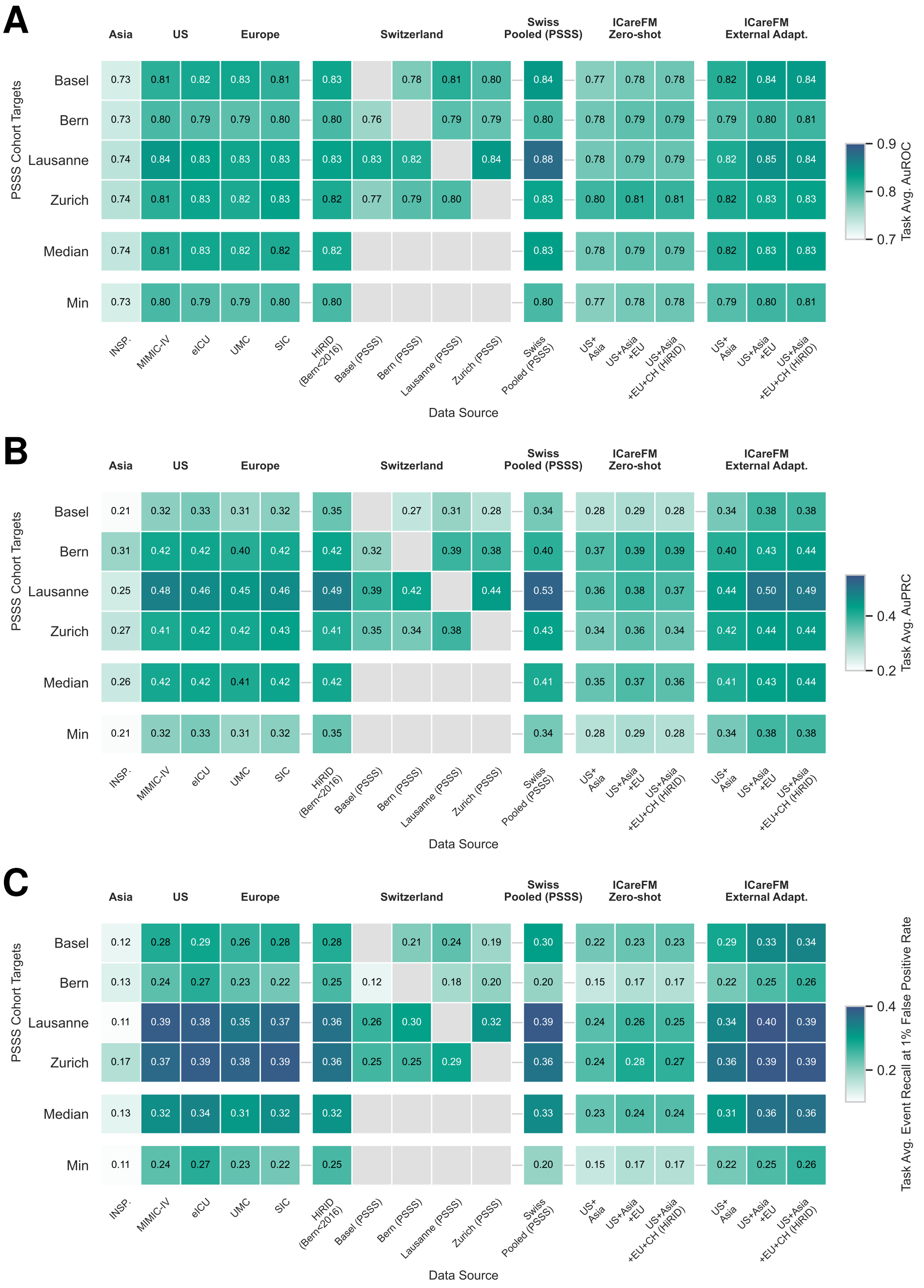


**Figure S23: Metric PSSS Transfer Study Heatmaps** Comprehensive out-of-distribution transfer performance to Swiss PSSS cohorts measured by task-averaged AuROC in **A**), AuPRC in **B**), and Event Recall at high precision 1% false positive rate (FPR). Each cell shows mean discrimination performance across all clinically relevant prediction tasks for a given target-source hospital pair. Data sources are organized by geographic region (Asia, US, Europe, Switzerland) followed by aggregated approaches (Swiss Pooled combining multiple PSSS centers but excluding the target, ICareFM Zero-shot with varying geographic training compositions, ICareFM External Adaptation as an externally task-tuned model on varying geographic training compositions). Gray cells indicate unavailable combinations (same source-target pairs). Median and minimum statistics across target cohorts demonstrate aggregate transfer robustness. ICareFM's zero-shot and external adaptation approaches achieve median AuPRC of 0.36/0.37 and 0.44 respectively, exceeding Swiss pooled baselines (0.41) and outperforming single-source hospital transfers in aggregate. The progressive improvement from US+Asia to US+Asia+Europe to US+Asia+Europe+Switzerland configurations illustrates the benefit of including geographically close data in the pretraining corpus of a foundation model. Worst case results indicate that the foundation model is helpful to stabilize transfer performance making it the most viable candidate if in doubt about which source model to choose. Trends are most pronounced for Event Recall at high precision, but also show similarly for AuPRC, and less visible but present for AuROC.

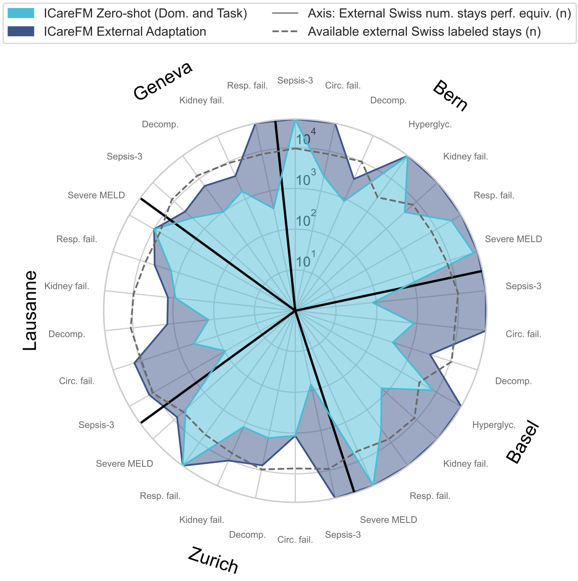


**Figure S24: ICareFM sample efficiency against same-country external data.** External Swiss patient equivalence for ICareFM zero-shot and external adaptation transfer to Swiss PSSS cohorts. Each spoke represents a task-cohort pair. Radial axis shows number of external Swiss patients (from other PSSS hospitals) required for supervised models to match ICareFM performance. Gray dashed line indicates available external data. Median zero-shot equivalence: 1,585 patients; median external adaptation: 32,031 patients, demonstrating foundation models' advantage over same-country pooled approaches.

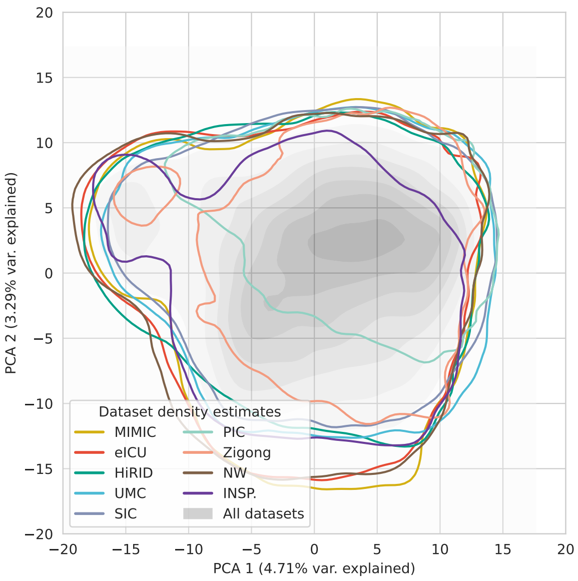


**Figure S25: Dataset-specific density distributions in ICareFM's latent space.** Kernel density estimation contours for nine ICU datasets projected onto the first two principal components of ICareFM's learned embeddings. Each colored contour represents the spatial distribution of patient states (timepoints) from a single dataset. The gray shaded region shows the aggregate density across all datasets combined. Despite being trained on harmonized data from diverse healthcare systems spanning three continents, the foundation model's learned representations exhibit similar spatial distributions across the first two principal components. Only datasets with the strongest distribution shifts, such as PICdb (pediatric care), show a partial spatial separation from the main cluster. These qualitative results suggest that the foundation model learned an underlying structure that focuses on clinically relevant patient characteristics shared across severely ill patients globally. This further supports the presented generalization results in Figure 2.

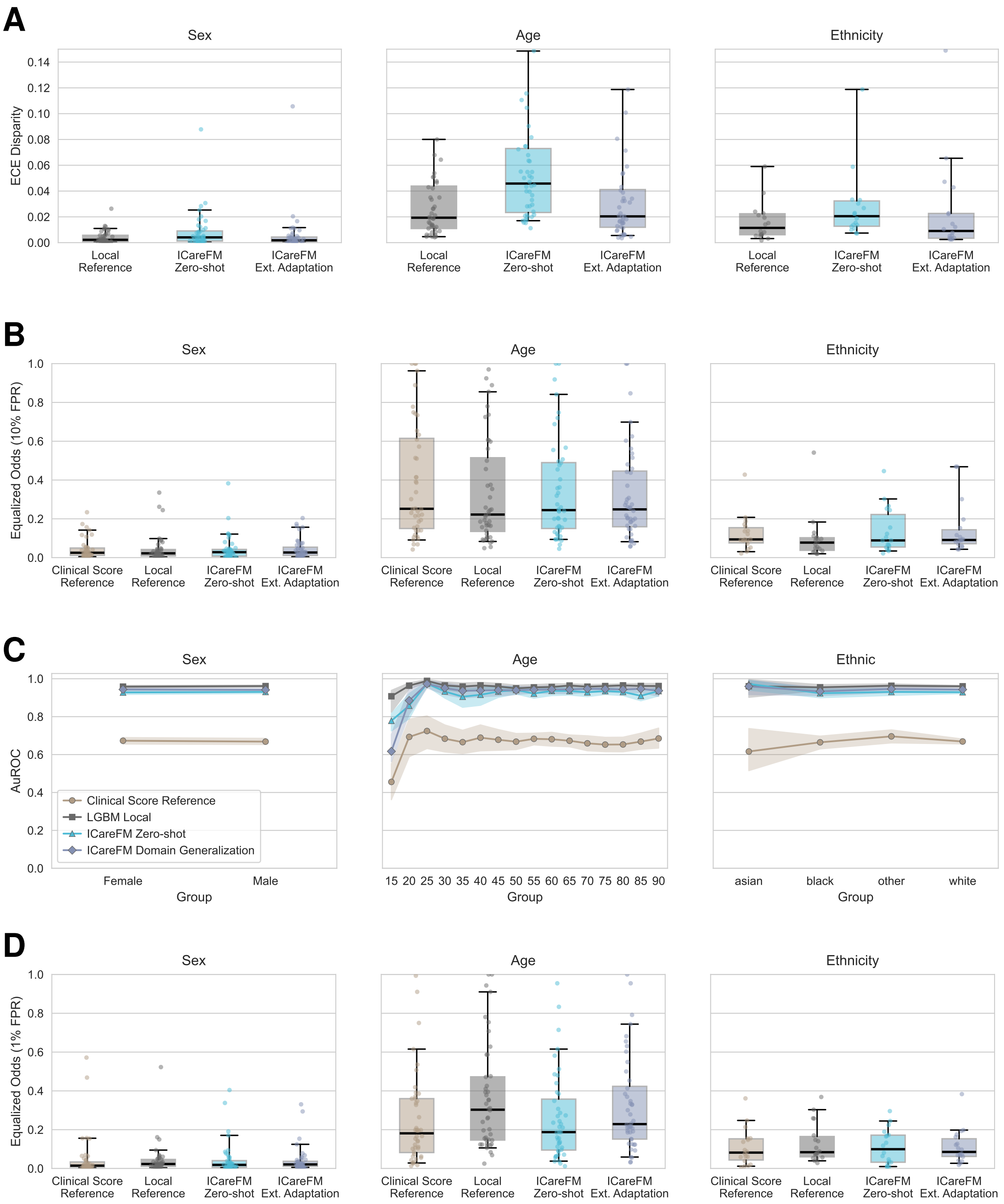


**Figure S26: Fairness analysis across demographic groups A)** Expected calibration error (ECE) disparity across dataset–task pairs, stratified by demographic property. Each point represents one dataset–task pair; lower values indicate more equitable performance across demographic groups. **B)** Equalized odds disparity (EOD) at 10% FPR across dataset–task pairs, stratified by demographic property. Each point represents one dataset–task pair; lower values indicate more equitable performance across demographic groups. **C)** Group-level AuROC for circulatory failure prediction on eICU, showing granular performance differences across demographic categories. Confidence intervals obtained from bootstrapping across patients ($N = 1000$). **D)** same as B) but at 1% FPR.

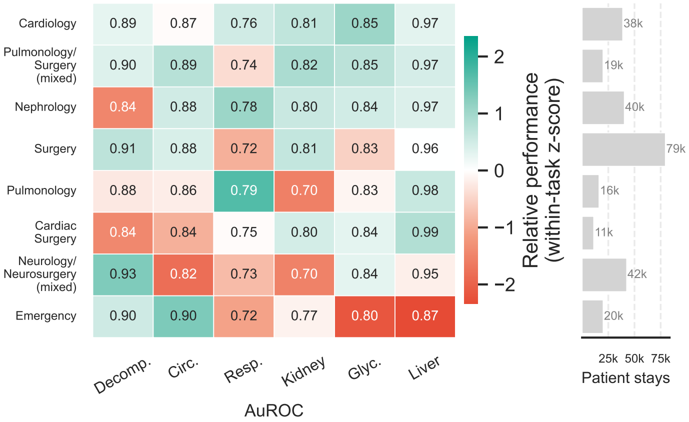


**Figure S27: Ward specialization sensitivity analysis.** Dual zero-shot AuROC stratified by ward specialization in the Charité cohort, colored by within-task relative performance.
